# Polygenic prediction of coronary heart disease among 130,000 Mexican adults

**DOI:** 10.1101/2024.12.20.24319332

**Authors:** Tianshu Liu, Jaime Berumen, Jason Torres, Jesus Alegre-Díaz, Paulina Baca, Carlos González-Carballo, Raul Ramirez-Reyes, Fernando Rivas, Diego Aguilar-Ramirez, Fiona Bragg, Will Herrington, Michael Hill, Eirini Trichia, Alejandra Vergara, Rachel Wade, Rory Collins, Pablo Kuri-Morales, Jonathan Emberson, Roberto Tapia-Conyer, Louisa Gnatiuc Friedrichs

**Affiliations:** Clinical Trial Service Unit and Epidemiological Studies Unit, Nuffield Department of Population Health (NDPH), University of Oxford, UK; Experimental Research Unit, Faculty of Medicine, National Autonomous University of Mexico (UNAM), Mexico City, Mexico; Health Data Research UK Oxford, University of Oxford, Oxford, UK; Instituto Tecnológico y de Estudios Superiores de Monterrey, Monterrey, Mexico; Faculty of Medicine, UNAM, Mexico City, Mexico

## Abstract

**Importance:** Coronary heart disease (CHD) is a leading cause of premature mortality globally. Most polygenic risk scores (PRSs) for CHD have been derived in populations of European ancestry. Their utility for CHD risk prediction in other populations is uncertain.

**Objective:** To evaluate the performance of eight established CHD PRSs in an admixed cohort of Mexican adults.

**Design, Setting, Participants:** 133,207 genotyped participants aged 35–79 years from the Mexico City Prospective Study (MCPS), a cohort recruited between 1998–2004, with follow-up for mortality until September 30, 2022.

**Exposures:** Eight PRSs for CHD, comprising between 44 and 6,472,620 single nucleotide polymorphism (SNP) variants, were selected and recreated for MCPS participants.

**Main outcomes and measure:** Premature CHD comprised prior doctor-diagnosed CHD at recruitment or CHD-related death before age 80. Logistic regression adjusted for age, sex, and the first seven genetic principal components (PCs) assessed PRS associations with CHD. Additional analyses evaluated performance by key participant characteristics, and after adjustment for vascular risk factors. Risk discrimination was assessed using C-statistics.

**Results:** Of the participants, 67% were women, the mean (±SD) age was 51±12 years, and Indigenous American ancestry averaged 67%. Premature CHD occurred in 5,163 participants (3.9%), including 1,901 prevalent and 3,479 fatal cases. All eight PRSs were positively and log-linearly associated with CHD, with odds ratios (ORs) per 1 SD increase ranging from 1.05 (95% CI, 1.03–1.08) to 1.29 (95% CI, 1.25–1.33). Associations were consistent across strata of age, ancestry, and relatedness. For six PRSs, however, associations were stronger in men than women (e.g., for the PRS with the strongest overall association: OR 1.37 [1.32–1.43] in men vs. 1.23 [1.18–1.28] in women). Adjustment for vascular risk factors did not substantially alter associations. Models including age, sex, genetic PCs and a PRS achieved an AUC of 0.72.

**Conclusion and Relevance:** In this Mexican population, existing PRSs derived from predominantly European ancestry populations predicted premature CHD independently of established vascular risk factors, particularly in men. Polygenic risk scores better capturing genetic variation in Latin American men and women may further enhance CHD risk prediction among Mexican and other Hispanic populations.

**Key points:** *Question:* To what extent do previously-published coronary heart disease (CHD) polygenic risk scores (PRS) predict CHD risk in an admixed Mexican population?

*Findings:* Among 133,207 Mexican adults aged 36-79 years, eight external PRSs were positively and log-linearly associated with CHD. Six of the eight showed significantly stronger associations with CHD in men compared to women. Multi-ancestry PRSs outperformed Eurocentric-ancestry PRSs.

*Meaning:* PRSs that better capture genetic variation in Latin-American men and women may further enhance CHD risk prediction among Mexican and other Hispanic populations.

## Introduction

Coronary heart disease (CHD) is a major cause of death and disability worldwide. It was estimated that in 2021, CHD was responsible for 9 million deaths (13% of deaths globally)^1^. Despite declines in age-specific CHD mortality rates in many developed countries over the past few decades, CHD remains the leading cause of death among adults globally and therefore its prevention remains a global public health priority.

CHD is estimated to have a heritability up to 50%^2, 3^ and several studies have shown that accounting for genetic predisposition can enhance CHD risk-stratification beyond conventional vascular risk factors^4–6^. Polygenic CHD risk scores (PRS) that combine multiple single nucleotide polymorphism (SNP) effects into a single genetic score have gained particular interest and have been shown to identify individuals with CHD risk equivalent to (or even higher than) those with rare monogenic mutations with large effects (e.g., familial hypercholesterolemia)^7^. Increasingly advanced methods have been developed to improve the predictive power of PRSs among European populations^8–10^, but the transferability of these PRSs to populations of other ancestries (with different linkage disequilibrium patterns and allele frequencies) is unclear.^11^ Since participants of non-European descent comprise only about 5% of those in existing GWAS studies,^12^ PRS evaluation and development in other populations has been limited.

In Mexico, the CHD mortality rate at ages 35-69 years has increased over the past 50 years^1^ (in men from about 60 per 100,000 in 1970 to 210 per 100,000 in 2020, and in women from about 40 per 100,000 in 1970 to 90 per 100,000 in 2020),^13^ in large part because of substantial population increases in major CHD risk factors over this period (in particular obesity and diabetes). The impact of varying prevalences (and effects) of major CHD risk factors on the performance of a PRS in a population is uncertain, especially for scores derived in one population but applied to another. Using data from the Mexico City Prospective Study (MCPS), the aim of this paper is to evaluate the predictive ability of eight existing CHD polygenic risk scores in an admixed population of Mexican adults with high levels of obesity and diabetes.

## Methods

### Study design and population

Details of the MCPS design, methods and population have been described previously^14^. Briefly, between 1998 and 2004, households in two districts of Mexico City (Coyoacán and Iztapalapa) were visited and household members aged 35 years or older were invited to participate. Of 112,333 households with eligible inhabitants, one or more individuals from 106,059 (95%) households consented to participate. Ethics approval was obtained from the Mexican Ministry of Health, the Mexican National Council for Science and Technology, and the University of Oxford. All participants provided written informed consent.

### Data collection

During household visits, trained nurses administered electronic questionnaires collecting information on sociodemographic and lifestyle factors, medication and disease history. Height, weight, hip circumference, waist circumference, and sitting blood pressure were measured using calibrated instruments and standard protocols. A non-fasting venous blood sample was collected into an EDTA vacutainer and separated into two plasma and one buffy coat aliquots for long-term storage at −150°C. Variants were genotyped on the Illumina GSAv2 beadchip and mapped to genome build GRCh38 (hg38)^15^. Variant quality control of genotype and individual-level missingness, departures from Hardy-Weinberg equilibrium, Mendel errors, genotype imputation (to TOPMed version 2), and per-individual proportions of Indigenous, European, African, and East Asian ancestry were estimated as previously described^15^.

### CHD polygenic risk scores

From a systematic search of PubMed, Embase and Medline, we selected eight published CHD-specific PRSs based either on their relevance to the ethnicities represented in the MCPS cohort, their extensive evaluation in the existing literature or the methodology they employed (see **WebTable 1** for details) to assess their transferability for predicting premature CHD risk within the MCPS cohort. Five of the selected PRSs (Khera et al^7^, Inouye et al^16^, Tamlander et al^17^, Oni-Orisan et al^18^ and Tada et al^19^) were derived from European ancestry populations using a variety of methods. The remaining three PRSs selected (Koyama et al^20^, Patel et al^21^ and Tcheandjieu et al^22^) involved multiple ancestries during training (i.e., GWAS source or PRS tuning). The eight CHD-specific PRSs were recreated for each MCPS participant using individual genotyped data and the pipeline developed by the PGS Catalog team^23, 24^. The SNP variant matching rates for all the selected PRSs were >85%, based on exact matching with no use of proxy SNPs. The PGS Catalog program pgsc_calc (v1.3.2) was performed using Nextflow-23.04.0^25^, PLINK-2.0^26^, and Anaconda Distribution 3.

### Follow-up for mortality

Participants are followed up for cause-specific mortality through probabilistic linkage (based on name, including phonetic coding of names, age, and sex) to the Mexican System for Epidemiologic Death Statistics (Subsistema Epidemiológico y Estadístico de Defunciones or SEED) electronic death registry in Mexico City, administered by the Ministry of Health. Field validation of more than 7000 matched deaths confirmed the reliability of the matching algorithm in over 95% of cases. Death registration in Mexico City is reliable and complete, with causes of almost all deaths certified medically^27^. Diseases recorded on death certificates are coded using the International Statistical Classification of Diseases and Related Health Problems, Tenth Revision (ICD-10), with subsequent review by study clinicians (who are unaware of baseline information) to recode, when necessary, the underlying cause of death^28^.

### Statistical analyses

The main analyses define “premature CHD” as either a self-reported history of heart attack or angina at the baseline assessment (for individuals aged <80 years at recruitment) or death before age 80 with CHD (ICD-10 codes I20-I25) listed anywhere on the death certificate. For individuals aged <80 years at recruitment, logistic regression was then used to estimate the association between each PRS and the odds of premature CHD risk, first by categorising each PRS into fifths (with the lowest fifth treated as the reference category and group-specific confidence intervals around each odds ratio [OR], including the reference group with OR of 1.0)^29^ and second by treating each PRS as a continuous variable (per 1SD increase). These regression models were adjusted initially just for age at recruitment, sex and the first seven genetic principal components (‘partial adjustment’) and then subsequently also for educational attainment (university or college, high school, elementary/other, none), waist-to-hip ratio (WHR), systolic and diastolic blood pressure (SBP and DBP), smoking status (never, former, current) and diabetes at recruitment (previously-diagnosed or HbA1c ≥6.5%, vs not) (‘full adjustment’). For covariates included in the fully adjusted model, the few participants with missing data had values imputed using the median for numeric variables and the most frequently selected category for categorical variables. The area under the receiver-operating-characteristic curve (i.e., the ‘AUC’) was used to assess model discrimination.

Analyses subsequently assessed for potential effect modification by age at risk, sex, proportion of Indigenous American ancestry, as well as other factors included in the fully adjusted model, with tests of heterogeneity or trend performed to assess whether the ORs in these subgroups varied significantly around the overall odds ratio. Sensitivity analyses included redefining the primary CHD outcome excluding deaths where CHD was not the underlying (i.e., the primary) cause, restricting to non-fatal self-reported myocardial infarction, angina or both at baseline, and restricting to the CHD death component (as listed anywhere on the death certificate and as listed as the underlying cause). Analyses of the primary CHD outcome were also repeated limited to participants who were unrelated to the 3^rd^ degree (based on identity-by-descent [IBD] estimates^15^). Finally, additional analyses extended the age range studied (participants and CHD events) up to age 90 years. Analyses were done using R (version 4.3.2).

## Results

### Included participants

Of the 159,755 participants recruited, 22,364 (14%) were excluded. These comprised 18,924 (11.8%) without genetic data available or genetic data meeting QC thresholds, a further 2,221 (1.4%) with uncertain mortality linkage or missing covariate data, and a further 1,219 (0.7%) aged ≥90 years at recruitment. Of the remaining 137,391 participants, 133,207 were aged 35-79 years at recruitment and 4,184 were aged 80-89 years.

### Baseline characteristics

Of the 133,207 participants aged 35-79 years at recruitment, the mean age was 51 years (SD 12 years), 33% were men, and 56% were unrelated to the 3^rd^ family degree. Average ancestry proportions were 67% Indigenous American, 28% European, 3% African and 1% East Asian (**Error! Reference source not found.**). The highest level of educational attainment was university or college for 16%, high school for 25% and elementary school for 47%. Women had a lower proportion of university or college attainment and higher proportion of high school attainment than men. Overall, 32% were current smokers and 20% former smokers, with a much higher proportion of men than women being current or former smokers. Mean SBP and DBP were 127 mmHg (SD 17 mmHg) and 83 mmHg (SD 10 mmHg) respectively. Self-reported history of CHD (heart attack or angina) was reported by 1,901 (1.4%) participants and stroke was reported by 1,414 (1.1%). 24,796 participants (19%) had self-reported diabetes or an HbA1c concentration indicative of undiagnosed diabetes (≥6.5%), while 8% reported having at least one other chronic disease. The baseline characteristics of all 137,391 participants aged 35-89 years at recruitment are shown in **Webtable 2**.

### Fatal and non-fatal CHD cases

During a median (IQR) follow-up in survivors of 20.3 years (19.4 to 21.5 years), there were 24,596 deaths at ages 35-79 years including 3,479 where CHD was listed on the death certificate (2,927 as the underlying cause and 552 elsewhere on the certificate). Of these 3,479 participants, 217 reported having CHD at recruitment and 3,262 did not. Consequently, in total, there were 5,163 participants who *either* had self-reported CHD prior to age 80 *or* died before age 80 with CHD listed on their death certificate (i.e., 3479 with CHD death before age 80 plus 1901 with pre-existing CHD at recruitment minus 217 with both). Extending the population to the 137,391 participants aged 35-89 at recruitment, 7,155 had either self-reported CHD prior to age 90 *or* died before age 90 with CHD listed on their death certificate.

### Relationship of each PRS to the odds of CHD

The eight evaluated CHD PRSs included between 44 and 6,472,620 SNPs; **Web figure 1** displays the pairwise correlations between the eight PRSs. The three European PRSs that included thousands of SNPs^7, 16, 17^ were all strongly correlated with each other. Two of the multi-ancestry PRSs^20, 21^ were also strongly correlated with each other and with the European PRSs. Baseline characteristics of those aged 35-79 years at recruitment by fifth of each of the eight PRSs are shown in **Webtables 3-10**.

**Figure 1:**
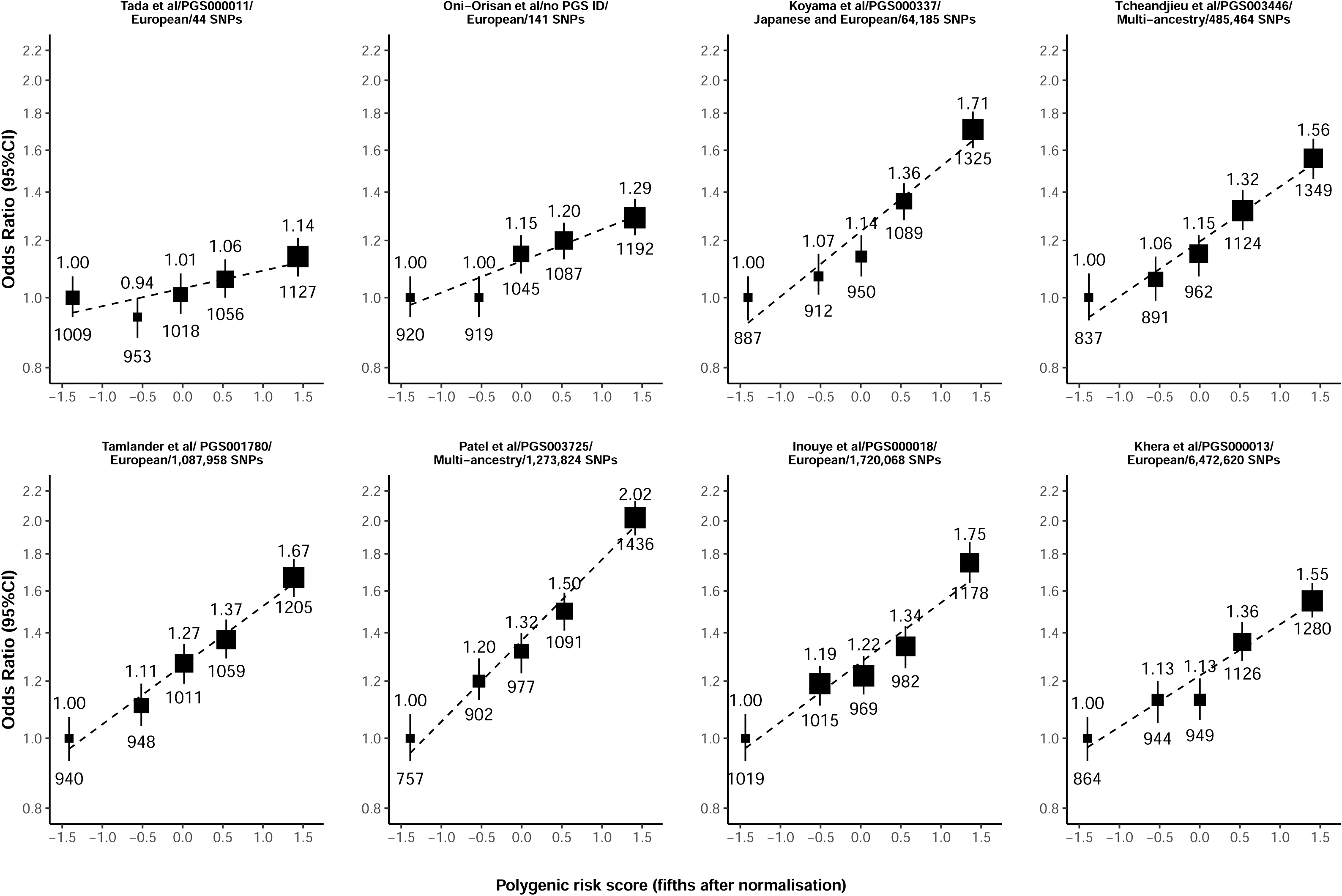
Odds of premature CHD by fifth of each PRS Analyses are adjusted for age, sex and the first 7 principal components. Each group is plotted against the mean of the normalized PRS. The vertical lines through each point represent 95% confidence intervals and are shown for each category (including the reference category with RR=1.0). The area of each square is inversely proportional to the square of the standard error of the log odds ratio (i.e., it is proportional to the amount of statistical information). ORs are shown above each point and the number of premature CHD cases below each point. PGS IDs refer to the ID number of PRSs on the PGS catalogue.

For all eight PRSs, the associations between the PRS and the odds of CHD were positive and approximately log-linear (**Figure 1**). Those with the highest fifth of genetic predisposition to CHD had 1.14 to 2.02 times the odds of CHD compared with those in the lowest (reference) fifth (**Figure 1**). The PRSs with fewer SNPs included tended to show weaker associations compared with the PRS which included thousands of SNPs. When adjusted for age, sex and the first seven genetic principal components, the PRS by Patel et al^21^ showed the strongest overall association with CHD, with each 1SD higher level of genetic predisposition associated with a 29% increase in the odds of CHD (OR=1.29, 95% CI 1.25-1.33) (**Figure 2**). By contrast, the weakest association was seen for the PRS by Tada et al^19^, with each 1SD higher level of genetic predisposition associated with only a 5% increase in the odds of CHD (OR=1.05, 1.03-1.08). For all eight PRSs, the strengths of association were attenuated marginally after further adjustment for conventional cardiovascular risk factors (**Figure 2**). The multi-ancestry sourced PRSs displayed relatively stronger associations with CHD risk compared with the European-sourced PRSs (ORs ranged from 1.19 to 1.29 versus 1.05 to 1.22, respectively).

**Figure 2:**
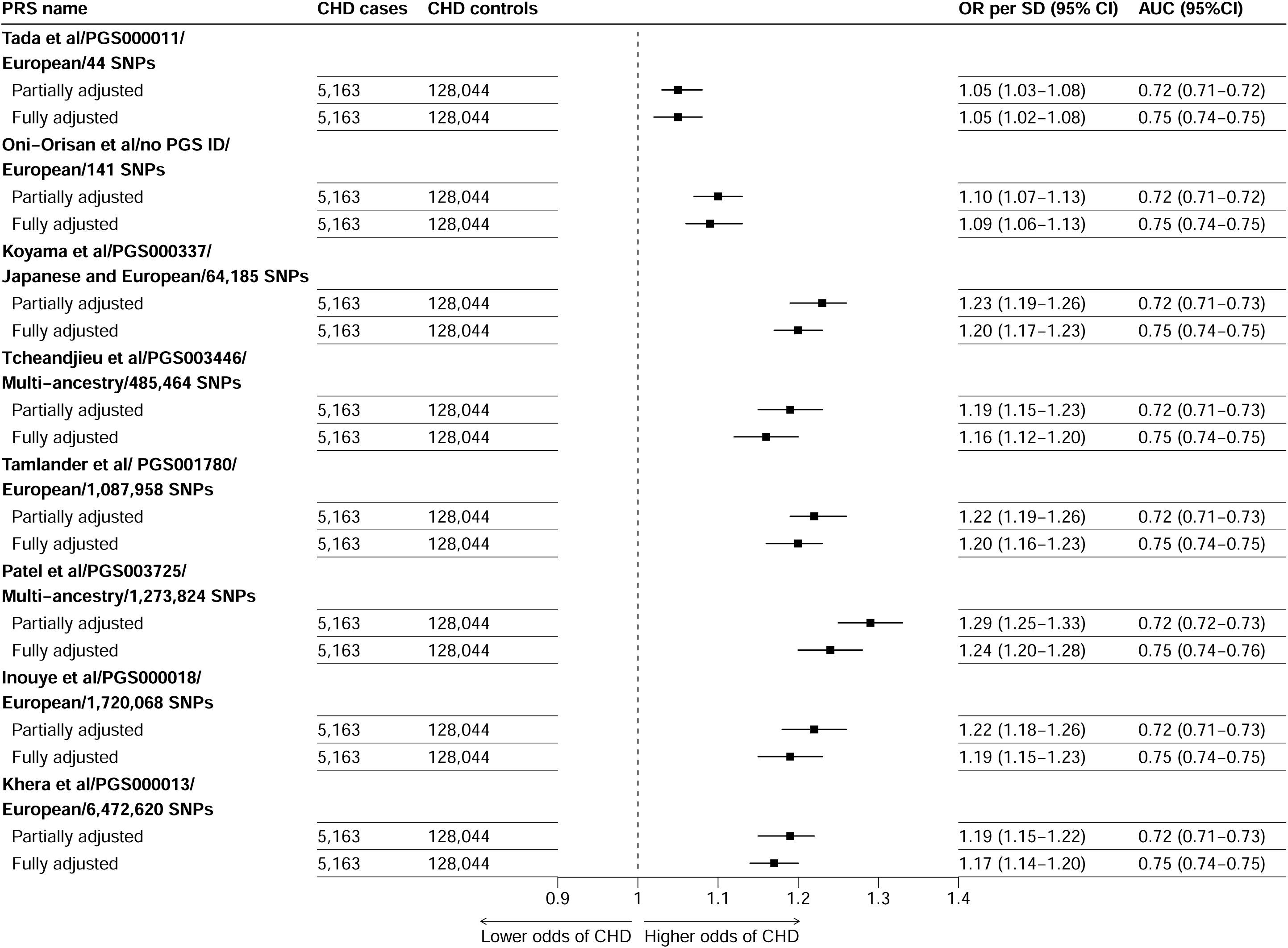
Odds of premature CHD per 1SD increase in each PRS In the partially adjusted model adjustment is for age, sex and the first 7 genetic principal components. The fully adjusted model is further adjusted for baseline waist-to-hip ratio, systolic and diastolic blood pressure, smoking status, level of education, and diabetes. Analyses are restricted to eligible participants with complete data on all covariates. PGS IDs refer to the ID number of PRSs on the PGS catalogue.

Based on age, sex, the first seven genetic principal components and the PRS, the model AUC was 0.72 for each PRS. For each PRS, further inclusion of other cardiovascular risk factors increased the AUC in each case to 0.75 (Error! Reference source not found.).

### Differences in genetic predisposition to CHD risk by sex

Two of the evaluated PRSs with only around a hundred genome-wide significant SNPs included (Tada et al^19^ [44 SNPs] and Oni-Orisan et al^18^ [141 SNPs]) showed similar strengths of associations in men and women (**Figure 3**). However, for the other six PRSs evaluated, each of which included thousands or hundreds of thousands of SNPs (Koyama et al^20^, Tcheandjieu et al^22^, Tamlander et al^17^, Patel et al^21^, Inouye et al^16^ and Khera et al^7^), genetic predisposition to subsequent CHD risk was significantly stronger for men compared to women. The (multi-ancestry sourced) Tcheandjieu et al^17^ PRS exhibited the greatest heterogeneity by sex, with an OR per 1SD higher genetic predisposition of 1.30 (95% CI 1.24-1.37) in men and 1.10 (95% CI 1.05-1.15) in women. In both men and women, the Patel et al^21^ PRS displayed the strongest association with CHD risk, with an OR of 1.37 (1.32-1.43) in men and 1.23 (1.18-1.28) in women. Despite this, the AUC was slightly higher for women than men for all PRS. Further adjustments for conventional vascular risk factors did not explain the sex differences observed.

**Figure 3:**
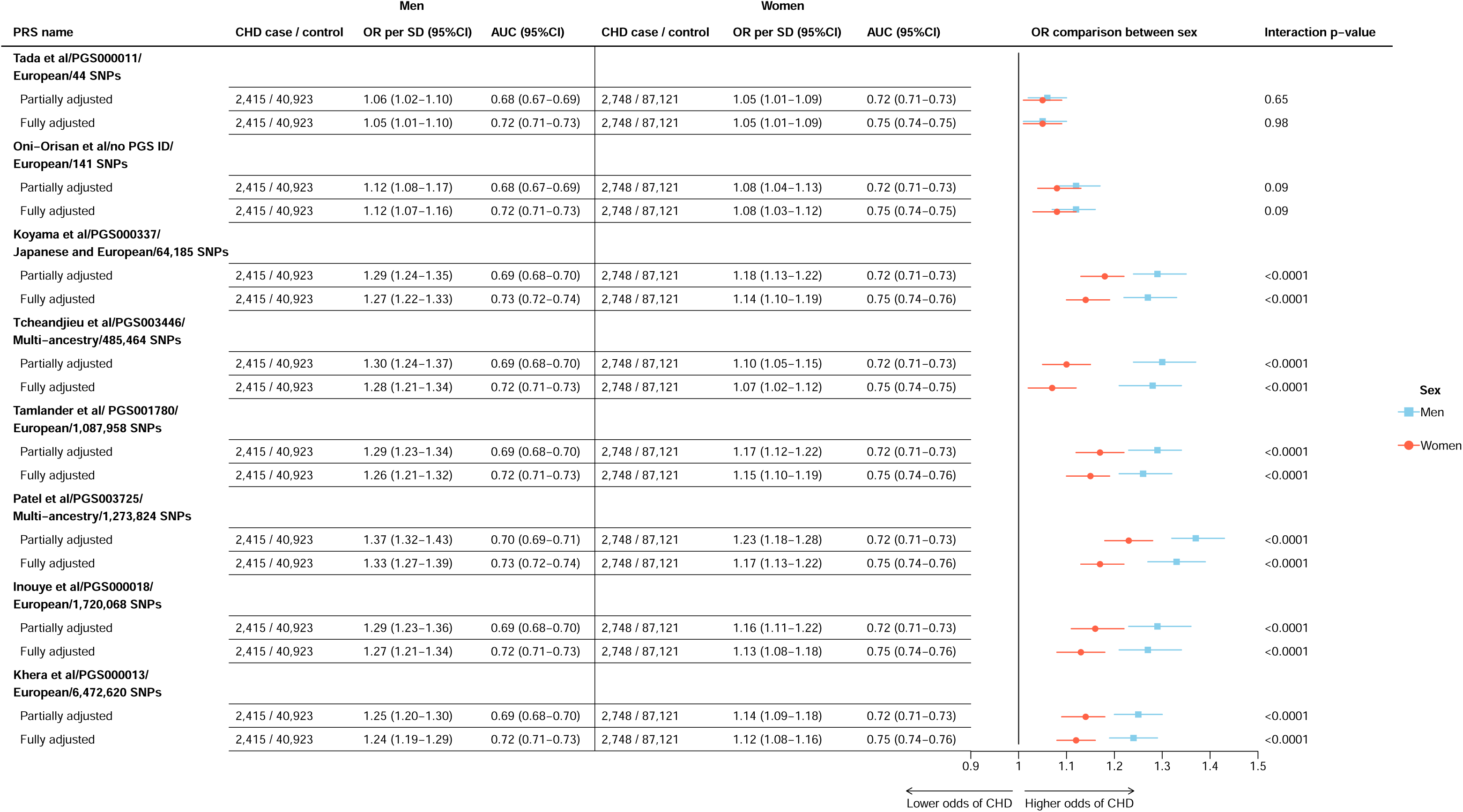
Odds of premature CHD per 1SD increase in each PRS, by sex Analyses as for Figure 2, now presented separately for men and women.

### Sensitivity analyses

Subgroup analyses of each PRS by age, education, waist-hip ratio, blood pressure, smoking status, history of diagnosed or undiagnosed diabetes, and indigenous ancestry proportion are shown in **Web figures 2-9**. Overall, results for each PRS were broadly consistent by levels of these factors. However, for the strongest overall performing PRSs, associations appeared slightly stronger at younger ages, among those with a higher waist-to-hip ratio, among those with a higher level of education, and among those with lower blood pressure.

Alternative definitions of CHD gave broadly similar results, with the weakest association generally seen for self-reported angina (albeit with wide confidence intervals) (**Web figures 10-17**). Restriction to those unrelated to the 3^rd^ degree or extension to those aged 35-89 years at recruitment (and CHD deaths before age 90) reinforced the main findings (**Web figures 18-20**).

## Discussion

In this large Mexican prospective study, we evaluated the relevance of genetic predisposition to CHD risk, using eight external PRSs derived from large European or multi-ancestry source populations. Overall, we found reasonable transferability and potential for predicting CHD risk in a Mexican population, with all of the evaluated PRSs showing clear positive log-linear associations with the odds of CHD at ages 35-79 years.

For six of the eight PRSs (the PRSs with tens of thousands if not millions of SNPs) the association observed in men was significantly stronger than that for women, perhaps due to the ability of PRSs with larger numbers of SNPs to better capture sex-specific genetic predisposition to CHD. A few other studies in European populations^16, 21, 30^ also reported differences in CHD risk predicted by PRSs between men than in women (with stronger associations in men than women). A previous study based on UKB^30^ investigated this difference by creating three mediating trait-based subscores (for blood pressure, blood lipids and body mass index) and found that only the blood pressure-mediated score exhibited a significant difference between men and women in its relation to CHD. Poorer genetic prediction of CHD in women may also reflect inherent genetic sex-biases that exist clinical definitions of CHD outcomes (e.g., progression, presentation, age at onset) that are dominated by studies of men, resulting in poorer performance in women.^31^

With the exception of sex, the associations of the PRSs with CHD were broadly similar according to other participant characteristics, and were comparable when using different definitions of CHD. Associations were largely independent of established vascular risk factors, which is consistent with previous reports from European populations^32–34^. Indeed, several recent developments that employed state-of-the-art methods for PRS construction^8–10, 35^, have shown that when integrated with existing clinical risk-score tools for CHD, genetic information can enhance screening in addition to existing clinical tools, and inform precision medicine efforts for better stratification of CHD risk among otherwise ‘healthy’ individuals^5, 32,36^.

Polygenic risk scores derived from European populations have demonstrated strong performance within European cohorts (as shown in **Webtable 1**) achieving AUC ranging from 0.75 to 0.81. However, their performance was somewhat weaker when applied to the current Mexican population. Consistent with our findings, several other studies reported weaker yet positive associations in genetic predisposition to CHD risk when European GWAS-sourced PRSs were subsequently applied to Hispanic populations^22, 37–39^, as well as other ethnically-diverse groups, including populations of both Middle-Eastern and African ancestries^22, 38–40^. The performance of a PRS depends on several factors, including sample size of the input GWAS and the accurate estimation of genetic effects for causal variants. These genetic effects can be influenced by population-specific environmental factors, which vary significantly across ancestries. Ideally, PRS for Hispanic individuals would use effect sizes derived from large, population-specific cohorts to capture both shared and unique genetic risk factors. Unfortunately, Hispanics remain significantly underrepresented in GWAS, leaving a critical gap in the development of PRS. Leveraging diverse population sources for GWAS SNP inputs may enhance transferability across diverse populations worldwide, as the true causal variants for CHD risk are likely shared across populations^34^. Indeed, in our analyses, the multi-ancestry GWAS-sourced PRSs from Patel et al^21^ and Koyama et al^20^, which were derived from some of the largest GWAS conducted to date, outperformed most of the European-dominant sourced-PRSs. However, the multi-ancestry PRSs also have limitations as they do not fully account for population-specific environmental influences and may overlook risk variants that are unique to underrepresented populations. The findings underscore the need for large-scale GWAS in Hispanic populations to ensure their fair representation in genetic research and to enhance the accuracy and applicability of PRSs in diverse populations.

The major strength of the present analysis is that it includes a large number of people from a previously-understudied population of Hispanic adults. The high-quality genetic data allowed CHD-specific PRSs to be reconstructed with careful consideration of genetic misclassification, genetic ancestry admixture and relatedness. A limitation of the study was the lack of data on non-fatal CHD incidence, although the large sample size and prolonged follow-up for mortality resulted in a reasonable number of CHD cases for analysis. The participants also arose from just two districts of Mexico City (Coyoacán and Iztapalapa) and therefore do not reflect the full genetic diversity of all Mexican adults. However, the findings from the current study clearly illustrate the potential for CHD polygenic risk scores to predict CHD risk in Mexican adults.

In summary, in this large Mexican population, existing polygenic risk scores were able to predict CHD risk independently of established vascular risk factors. However, current PRSs do not fully reflect the genetic architecture of CHD in Mexico. Ancestry-specific instruments that more closely represent genetic variation in Mexico may further enhance polygenic prediction of CHD risk in Mexican adults.

## Data Availability

Data from the Mexico City Prospective Study are available to bona fide researchers. The study's Data and Sample Sharing policy can be downloaded (in English or Spanish: https://www.ctsu.ox.ac.uk/research/mcps). Available study data can be examined in detail through the study's Data Showcase (https://datashare.ndph.ox.ac.uk/mexico/). MCPS ancestry-specific allele frequencies are available in a public browser (https://rgc-mcps.regeneron.com/).

## Acknowledgements

We thank the study participants and staff. This work was supported by the Mexican Health Ministry; the National Council of Science and Technology for Mexico; Wellcome [058299/Z/99]; Cancer Research UK; the British Heart Foundation [RE/13/1/30181]; and the UK Medical Research Council [MC_UU_00017/2, MR/Z504543/1]. Genotyping was funded through an academic partnership between the National Autonomous University of Mexico, the University of Oxford, Regeneron and AstraZeneca. The computational aspects of this research were supported by the Wellcome Trust Core Award Grant Number 203141/Z/16/Z and the NIHR Oxford BRC. The funding sources had no role in the design, conduct or analysis of the study or the decision to submit the manuscript for publication.

## Contributors

JA-D, RT-C, and PK-M established the cohort. RC provided support during its inception and RC and JRE have provided support to enhance it over the years. TL designed and conducted the analyses and wrote the first version of the manuscript, under the supervision of JT, JRE and LGF. MH provided laboratory support. All authors contributed to data acquisition, analysis, or interpretation of the data, to the critical revision of the manuscript for important intellectual content, and have seen and approved the final version. TL, JT and JRE had full access to the data, and take responsibility for the integrity and accuracy of the analysis.

## Rights retention statement

For the purposes of open access, the authors have applied a Creative Commons Attribution (CC BY) licence to any Author Accepted Manuscript version arising.

---

**Table 1:** Baseline characteristics of 133,207 participants aged 35-79 years Numbers are n (%) or mean (SD). SBP=systolic blood pressure, DBP=diastolic blood pressure, BMI=body mass index, HDL-C=high density lipoprotein cholesterol, LDL-C=low density lipoprotein cholesterol, HbA1c=glycosylated haemoglobin A1c *Self-reported previous diagnoses unless otherwise stated. †Self-reported previously-diagnosed diabetes or glycosylated haemoglobin ≥6.5%. ‡Other diseases include self-reported emphysema, chronic kidney disease, peptic ulcer, liver cirrhosis, and peripheral arterial disease.

|  | Men (n=43,338) | Women (n=89,869) | All (n=133,207) |
| --- | --- | --- | --- |
| **Age, years** | 52.0 (12.1) | 50.8 (11.7) | 51.2 (11.8) |
| **Resident of Coyoacán** | 17,958 (41%) | 33,057 (37%) | 51,015 (38%) |
| **Ancestry admixture percentage** |  |  |  |
| Indigenous American | 66.5 (18.0) | 67.0 (17.8) | 66.8 (17.9) |
| African | 3.4 (2.8) | 3.5 (2.8) | 3.4 (2.8) |
| East Asian | 1.4 (2.0) | 1.4 (1.7) | 1.4 (1.8) |
| European | 28.7 (16.3) | 28.1 (16.1) | 28.3 (16.2) |
| **Highest attained educational level** |  |  |  |
| University/college | 10,340 (24%) | 10,369 (12%) | 20,709 (16%) |
| High school | 11,468 (26%) | 21,964 (24%) | 33,432 (25%) |
| Elementary | 17,902 (41%) | 45,005 (50%) | 62,907 (47%) |
| Other | 3,616 (8%) | 12,475 (14%) | 16,091 (12%) |
| Missing | 12 (<0.1%) | 56 (<0.1%) | 68 (<0.1%) |
| **Smoking status** |  |  |  |
| Never | 8,719 (20%) | 55,872 (62%) | 64,591 (48%) |
| Former | 13,060 (30%) | 13,116 (15%) | 26,176 (20%) |
| Current | 21,521 (50%) | 20,809 (23%) | 42,330 (32%) |
| Missing | 38 (<0.1%) | 72 (<0.1%) | 110 (<0.1%) |
| **Alcohol intake** |  |  |  |
| Never | 2,660 (6%) | 23,443 (26%) | 26,103 (20%) |
| Former | 7,598 (18%) | 10,653 (12%) | 18,251 (14%) |
| Current | 33,062 (76%) | 55,741 (62%) | 88,803 (67%) |
| Missing | 18 (<0.1%) | 32 (<0.1%) | 50 (<0.1%) |
| **Physical measures** |  |  |  |
| SBP, mmHg | 128.5 (15.8) | 126.6 (16.9) | 127.2 (16.6) |
| DBP, mmHg | 84.4 (9.9) | 82.5 (10.2) | 83.1 (10.2) |
| BMI, kg/m<sup>2</sup> | 28.0 (4.3) | 29.6 (5.3) | 29.1 (5.1) |
| Waist-to-hip Ratio | 0.95 (0.07) | 0.88 (0.07) | 0.90 (0.08) |
| **Laboratory measurements** |  |  |  |
| HDL-C, mmol/L | 0.93 (0.19) | 1.03 (0.22) | 1.00 (0.21) |
| LDL-C, mmol/L | 2.39 (0.79) | 2.49 (0.79) | 2.46 (0.79) |
| Triglycerides, mmol/L | 1.65 (0.68) | 1.54 (0.64) | 1.57 (0.66) |
| HbA1c, % | 6.10 (1.72) | 6.10 (1.71) | 6.10 (1.71) |
| eGFR, ml/min/1.73m<sup>2</sup><sup>§</sup> | 101.0 (15.9) | 102.0 (16.1) | 101.7 (16.0) |
| **Prior disease** |  |  |  |
| Coronary heart disease | 848 (2%) | 1,053 (1%) | 1,901 (1%) |
| Stroke | 485 (1%) | 929 (1%) | 1,414 (1%) |
| Cancer | 266 (1%) | 1,314 (1%) | 1,580 (1%) |
| Diabetes<sup>†</sup> | 8,250 (19%) | 16,546 (18%) | 24,796 (19%) |
| Other<sup>‡</sup> | 2,276 (5%) | 8,837 (10%) | 11,113 (8%) |

<sup>§</sup>Calculated using the 2021 CKD-EPI equation, based on NMR-measured creatinine levels.

