## Appendix for "Polygenic prediction of coronary heart disease among 130,000 Mexican adults"

### Online appendix


| | Page |
|---|---|
| **Webtables** | |
| 1. CHD polygenic risk scores selected for evaluation | 2 |
| 2. Baseline characteristics of 137,391 participants aged 35-89 years | 3 |
| Baseline characteristics of 133,207 participants aged 35-79 years by fifth of PRS by: | |
| 3. Tada *et al.* | 4 |
| 4. Oni-Orisan *et al.* | 5 |
| 5. Koyama *et al.* | 6 |
| 6. Tcheandjieu *et al.* | 7 |
| 7. Tamlander *et al.* | 8 |
| 8. Patel *et al.* | 9 |
| 9. Inouye *et al.* | 10 |
| 10. Khera *et al.* | 11 |
| **Webfigures** | |
| 1. Pairwise correlations between the eight selected PRS | 12 |
| Odds of premature CHD per 1SD increase in each PRS: | |
| 2. At different ages at risk | 13 |
| 3. By highest level of education | 14 |
| 4. By waist-to-hip ratio | 15 |
| 5. By systolic blood pressure | 16 |
| 6. By diastolic blood pressure | 17 |
| 7. By smoking status | 18 |
| 8. By baseline diabetes status | 19 |
| 9. By level of Indigenous American ancestry | 20 |
| Odds ratios per 1SD increase in each PRS among participants aged 35-79 years at recruitment, for the outcome of: | |
| 10. Baseline self-reported angina | 21 |
| 11. Baseline self-reported heart attack | 22 |
| 12. Baseline self-reported angina *or* heart attack | 23 |
| 13. Premature CHD, defined as baseline self-reported angina or heart attack, *or* death before age 80 with CHD listed as the *primary* cause of death | 24 |
| 14. Death before age 80 with CHD listed *anywhere* on the death certificate | 25 |
| 15. Death before age 80 with CHD listed as the *primary* cause on the death certificate | 26 |
| Sensitivity analyses | |
| 16. Varying the CHD outcome, with partial adjustments | 27 |
| 17. Varying the CHD outcome, with full adjustments | 28 |
| 18. Odds of premature CHD among participants unrelated to the 3rd degree | 29 |
| 19. Odds of CHD before age 90 years per 1SD increase in each PRS, among participants aged 35-89 years at recruitment | 30 |
| 20. Odds of CHD before age 90 years by fifth of each PRS, among participants aged 35-89 years at recruitment | 31 |

### Webtable 1: CHD polygenic risk scores selected for evaluation

| Authors / PRS ID in PGS catalogue/Ancestry | PRS characteristics | CHD odds or hazard ratio (95% CI)* | AUC (95%CI) |
|---|---|---|---|
| Tada et al[w1] /PGS000011/European | A PRS constructed using 50 SNPs and European GWAS. **GWAS:** European dominant (~95%) with a small proportion of Asian samples **Evaluation sample:** 100% European | HR per SD: 1.23 (1.18-1.28) | 0.75 (no CI reported) |
| Oni-orisan et al[w2] / No PGS ID/European | A PRS constructed using European GWAS and 164 genome-wide significant CHD risk SNPs from UKB and CC4D GWAS. It was evaluated among datasets that contains Hispanic participants (n=1538). **GWAS:** European dominant **Evaluation sample:** Multi-ancestry | HR per SD: 1.87 (1.19-2.95) | NR |
| Koyama et al[w3] /PGS000337/Japanese and European | A trans-ancestry PRS that used Japanese and European genetic information during construction. **GWAS:** European and Japanese dominant **Training sample:** Japanese dominant **Evaluation sample:** Japanese dominant | OR per SD: 1.84 (1.74-1.94) | 0.67 (0.66–0.69) |
| Tcheandjieu et al[w4] /PGS003446/Multi-ancestry | A trans-ancestry PRS that used Hispanic, European and Japanese genetic information during construction using pruning and thresholding [w5]. **GWAS:** Multi-ancestry (Hispanics, African, Japanese, European) **Training sample:** European, African and Hispanics **Evaluation sample:** Hispanic | OR per SD: 1.43 (1.27-1.61) | NR |
| Tamlander et al[w6] / PGS001780/European | A PRS developed using a novel method, PRS-CS-auto. **GWAS:** European dominant (~80%) with a small proportion of Asian samples **Evaluation sample:** European | OR per SD (in UK Biobank): 1.72 (1.70-1.75) | 0.79 (0.79–0.80) |
| Patel et al[w7] /PGS003725/Multi-ancestry | A trans-ancestry and multi-trait PRS derived using Hispanic, European, African and Asian genetic information. **GWAS:** Multi-ancestry (Hispanics, African, Japanese, European) **Training sample:** European **Evaluation sample:** Hispanic | OR per SD: 1.61 (1.53-1.70) | NR |
| Inouye et al[w8] /PGS000018/European | A meta-score based on the weighted average of 3 PRSs. **GWAS:** European dominant (~90%) with a small proportion of Asian samples **Training sample:** European dominant **Evaluation sample**: European dominant | HR per SD: 1.71 (1.68-1.73) | 0.79 (no CI reported) |
| Khera et al[w9] /PGS000013/European | A PRS developed using LDPred2[w10] with a large European dominant GWAS input. **GWAS:** European dominant (~80%) with a small proportion of Asian samples **Training sample:** European dominant **Evaluation sample:** European dominant | OR per SD (in training sample): 1.72 (1.67-1.78) | 0.81 (0.81-0.81) |

* In the ‘evaluation sample’ unless otherwise stated. AUC=Area under the receiver operating characteristic curve. CHD=Coronary heart disease. CI=Confidence interval. OR= Odds ratio. HR=Hazard ratio. NR=Not reported

### Webtable 2: Baseline characteristics of 137,391 participants aged 35-89 years

| | Men (n=44,853) | Women (n=92,538) | All (n=137,391) |
|---|---|---|---|
| **Age, years** | 53.1 (13.1) | 51.8 (12.8) | 52.2 (12.9) |
| **Resident of Coyoacán** | 18,425 (41%) | 33,947 (37%) | 52,372 (38%) |
| **Ancestry admixture percentage** | | | |
| Indigenous American | 66.4 (18.1) | 67.0 (17.9) | 66.8 (17.9) |
| African | 3.4 (2.8) | 3.5 (2.8) | 3.4 (2.8) |
| East Asian | 1.4 (2.0) | 1.4 (1.7) | 1.4 (1.8) |
| European | 28.7 (16.4) | 28.2 (16.2) | 28.4 (16.2) |
| **Highest attained educational level** | | | |
| University/college | 10,443 (23%) | 10,421 (11%) | 20,864 (15%) |
| High school | 11,596 (26%) | 22,116 (24%) | 33,712 (25%) |
| Elementary | 18,694 (42%) | 46,226 (50%) | 64,920 (47%) |
| Other | 4,104 (9%) | 13,716 (15%) | 17,820 (13%) |
| Missing | 16 (<0.1%) | 59 (<0.1%) | 75 (<0.1%) |
| **Smoking status** | | | |
| Never | 9,084 (20%) | 57,951 (63%) | 67,035 (49%) |
| Former | 13,876 (31%) | 13,553 (15%) | 27,429 (20%) |
| Current | 21,852 (49%) | 20,958 (23%) | 42,810 (31%) |
| Missing | 41 (<0.1%) | 76 (<0.1%) | 117 (<0.1%) |
| **Alcohol intake** | | | |
| Never | 2,859 (6%) | 24,582 (27%) | 27,441 (20%) |
| Former | 8,179 (18%) | 11,160 (12%) | 19,339 (14%) |
| Current | 33,793 (75%) | 56,763 (61%) | 90,556 (66%) |
| Missing | 22 (<0.1%) | 33 (<0.1%) | 55 (<0.1%) |
| **Physical measures** | | | |
| SBP, mmHg | 128.8 (15.9) | 127.0 (17.2) | 127.6 (16.8) |
| DBP, mmHg | 84.4 (10.0) | 82.5 (10.3) | 83.1 (10.2) |
| BMI, kg/m² | 27.9 (4.3) | 29.6 (5.3) | 29.0 (5.1) |
| Waist-to-hip Ratio | 0.95 (0.07) | 0.88 (0.07) | 0.90 (0.08) |
| **Laboratory measurements** | | | |
| HDL-C, mmol/L | 0.93 (0.19) | 1.04 (0.22) | 1.00 (0.21) |
| LDL-C, mmol/L | 2.38 (0.79) | 2.49 (0.79) | 2.45 (0.79) |
| Triglycerides, mmol/L | 1.64 (0.68) | 1.53 (0.64) | 1.57 (0.66) |
| HbA1c, % | 6.10 (1.70) | 6.10 (1.70) | 6.10 (1.70) |
| eGFR, ml/min/1.73m²[§] | 100.3 (16.4) | 101.3 (16.6) | 100.9 (16.5) |
| **Prior disease[*]** | | | |
| Coronary heart disease | 937 (2%) | 1,152 (1%) | 2,089 (2%) |
| Stroke | 547 (1%) | 1,011 (1%) | 1,558 (1%) |
| Cancer | 311 (1%) | 1,377 (1%) | 1,688 (1%) |
| Diabetes[†] | 8,599 (19%) | 17,242 (19%) | 25,841 (19%) |
| Other[‡] | 2,449 (5%) | 9,128 (10%) | 11,577 (8%) |

Numbers are *n* (%) or mean (SD). SBP=systolic blood pressure, DBP=diastolic blood pressure, BMI=body mass index, HDL-C=high density lipoprotein cholesterol, LDL-C=low density lipoprotein cholesterol, HbA1c=glycosylated haemoglobin A1c

[*]Self-reported previous diagnoses unless otherwise stated.

[†]Self-reported previously-diagnosed diabetes or glycosylated haemoglobin ≥6.5%.

[‡]Other diseases include self-reported emphysema, chronic kidney disease, peptic ulcer, liver cirrhosis, and peripheral arterial disease.

[§]Calculated using the 2021 CKD-EPI equation, based on NMR-measured creatinine levels.

### Webtable 3: Baseline characteristics of 133,207 participants aged 35-79 by fifth of PRS by Tada *et al.*

| **Tada et al/PGS000011/ European/44 SNPs** | **Fifth of the PRS distribution** | | | | | **All (n=133,207)** |
|---|---|---|---|---|---|---|
| | **1 (n=26,642)** | **2 (n=26,641)** | **3 (n=26,641)** | **4 (n=26,641)** | **5 (n=26,642)** | |
| **Age, years** | 51.3 (11.9) | 51.3 (11.9) | 51.2 (11.9) | 51.2 (11.8) | 51.2 (11.7) | 51.2 (11.8) |
| **Resident of Coyoacán** | 10,079 (38%) | 10,295 (39%) | 10,077 (38%) | 10,257 (39%) | 10,307 (39%) | 51,015 (38%) |
| **Ancestry admixture percentage** | | | | | | |
| Indigenous American | 67.1 (17.5) | 66.8 (17.9) | 66.7 (18.0) | 67.0 (18.0) | 66.6 (17.8) | 66.8 (17.9) |
| African | 3.5 (3.0) | 3.5 (2.9) | 3.4 (2.8) | 3.4 (2.7) | 3.4 (2.7) | 3.4 (2.8) |
| East Asian | 1.4 (1.8) | 1.4 (1.6) | 1.4 (2.0) | 1.4 (1.8) | 1.4 (1.9) | 1.4 (1.8) |
| European | 27.9 (15.8) | 28.4 (16.3) | 28.4 (16.3) | 28.2 (16.3) | 28.6 (16.1) | 28.3 (16.2) |
| **Highest attained educational level** | | | | | | |
| University/college | 4,091 (15%) | 4,164 (16%) | 4,143 (16%) | 4,183 (16%) | 4,128 (16%) | 20,709 (16%) |
| High school | 6,695 (25%) | 6,776 (25%) | 6,560 (25%) | 6,613 (25%) | 6,788 (25%) | 33,432 (25%) |
| Elementary | 12,557 (47%) | 12,444 (47%) | 12,679 (48%) | 12,668 (48%) | 12,559 (47%) | 62,907 (47%) |
| Other | 3,282 (12%) | 3,244 (12%) | 3,244 (12%) | 3,167 (12%) | 3,154 (12%) | 16,091 (12%) |
| Missing | 17 (0%) | 13 (0%) | 15 (0%) | 10 (0%) | 13 (0%) | 68 (0%) |
| **Smoking status** | | | | | | |
| Never | 12,874 (48%) | 12,888 (48%) | 12,929 (49%) | 13,051 (49%) | 12,849 (48%) | 64,591 (48%) |
| Former | 5,199 (20%) | 5,156 (19%) | 5,225 (20%) | 5,213 (20%) | 5,383 (20%) | 26,176 (20%) |
| Current | 8,545 (32%) | 8,576 (32%) | 8,462 (32%) | 8,362 (31%) | 8,385 (32%) | 42,330 (32%) |
| Missing | 24 (0%) | 21 (0%) | 25 (0%) | 15 (0%) | 25 (0%) | 110 (0%) |
| **Alcohol intake** | | | | | | |
| Never | 5,082 (19%) | 5,384 (20%) | 5,245 (20%) | 5,268 (20%) | 5,124 (19%) | 26,103 (20%) |
| Former | 3,701 (14%) | 3,571 (13%) | 3,683 (14%) | 3,605 (14%) | 3,691 (14%) | 18,251 (14%) |
| Current | 17,847 (67%) | 17,680 (66%) | 17,699 (66%) | 17,759 (67%) | 17,818 (67%) | 88,803 (67%) |
| Missing | 12 (0%) | 6 (0%) | 14 (0%) | 9 (0%) | 9 (0%) | 50 (0%) |
| **Physical measures** | | | | | | |
| SBP, mmHg | 127.0 (16.5) | 127.1 (16.6) | 127.3 (16.6) | 127.2 (16.6) | 127.6 (16.7) | 127.2 (16.6) |
| DBP, mmHg | 83.0 (10.2) | 83.0 (10.1) | 83.1 (10.1) | 83.2 (10.2) | 83.2 (10.3) | 83.1 (10.2) |
| BMI, kg/m² | 29.2 (5.1) | 29.1 (5.1) | 29.1 (5.1) | 29.1 (5.0) | 29.1 (5.0) | 29.1 (5.1) |
| Waist-to-hip Ratio | 0.90 (0.08) | 0.90 (0.08) | 0.90 (0.08) | 0.90 (0.08) | 0.90 (0.08) | 0.90 (0.08) |
| **Laboratory measurements** | | | | | | |
| HDL-C, mmol/L | 1.00 (0.21) | 1.00 (0.21) | 1.00 (0.21) | 1.00 (0.21) | 1.00 (0.21) | 1.00 (0.21) |
| LDL-C, mmol/L | 2.45 (0.79) | 2.47 (0.79) | 2.46 (0.79) | 2.46 (0.79) | 2.45 (0.79) | 2.46 (0.79) |
| Triglycerides, mmol/L | 1.56 (0.65) | 1.57 (0.65) | 1.57 (0.66) | 1.58 (0.66) | 1.58 (0.66) | 1.57 (0.66) |
| HbA1c, % | 6.09 (1.70) | 6.09 (1.70) | 6.09 (1.71) | 6.09 (1.70) | 6.12 (1.75) | 6.10 (1.71) |
| eGFR, ml/min/1.73m²[§] | 101.7 (16.1) | 101.6 (16.2) | 101.7 (15.9) | 101.6 (15.9) | 101.6 (16.1) | 101.7 (16.0) |
| **Prior disease**[*] | | | | | | |
| Coronary heart disease | 375 (1%) | 380 (1%) | 346 (1%) | 386 (1%) | 414 (2%) | 1,901 (1%) |
| Stroke | 249 (1%) | 292 (1%) | 295 (1%) | 287 (1%) | 291 (1%) | 1,414 (1%) |
| Cancer | 335 (1%) | 318 (1%) | 317 (1%) | 315 (1%) | 295 (1%) | 1,580 (1%) |
| Diabetes[†] | 4,955 (19%) | 4,901 (18%) | 4,900 (18%) | 4,938 (19%) | 5,102 (19%) | 24,796 (19%) |
| Other[‡] | 2,197 (8%) | 2,204 (8%) | 2,166 (8%) | 2,282 (9%) | 2,264 (8%) | 11,113 (8%) |

Numbers are n (%) or mean (SD). SBP=systolic blood pressure, DBP=diastolic blood pressure, BMI=body mass index, HDL-C=high density lipoprotein cholesterol, LDL-C=low density lipoprotein cholesterol, HbA1c=glycosylated haemoglobin A1c

[*]Self-reported previous diagnoses unless otherwise stated.

[†]Self-reported previously-diagnosed diabetes or glycosylated haemoglobin ≥6.5%.

[‡]Other diseases include self-reported emphysema, chronic kidney disease, peptic ulcer, liver cirrhosis, and peripheral arterial disease.

[§]Calculated using the 2021 CKD-EPI equation, based on NMR-measured creatinine levels.

### Webtable 4: Baseline characteristics of 133,207 participants aged 35-79 by fifth of PRS by Oni-Orisan *et al.*

| Oni-Orisan et al/no PGS ID/ European/141 SNPs | Fifth of the PRS distribution | | | | | All (n=133,207) |
|---|---|---|---|---|---|---|
| | 1 (n=26,642) | 2 (n=26,641) | 3 (n=26,641) | 4 (n=26,641) | 5 (n=26,642) | |
| **Age, years** | 51.3 (11.9) | 51.3 (11.8) | 51.2 (11.8) | 51.1 (11.8) | 51.3 (11.8) | 51.2 (11.8) |
| **Resident of Coyoacán** | 9,883 (37%) | 10,000 (38%) | 10,221 (38%) | 10,280 (39%) | 10,631 (40%) | 51,015 (38%) |
| **Ancestry admixture percentage** | | | | | | |
| Indigenous American | 69.0 (17.5) | 68.4 (17.8) | 67.5 (17.8) | 66.2 (17.9) | 63.1 (17.7) | 66.8 (17.9) |
| African | 3.2 (2.6) | 3.3 (2.7) | 3.4 (2.8) | 3.5 (2.9) | 3.8 (3.0) | 3.4 (2.8) |
| East Asian | 1.4 (1.5) | 1.4 (1.5) | 1.4 (1.4) | 1.4 (2.1) | 1.5 (2.3) | 1.4 (1.8) |
| European | 26.4 (15.8) | 26.9 (16.1) | 27.7 (16.0) | 28.8 (16.2) | 31.6 (16.1) | 28.3 (16.2) |
| **Highest attained educational level** | | | | | | |
| University/college | 3,932 (15%) | 4,007 (15%) | 4,029 (15%) | 4,224 (16%) | 4,517 (17%) | 20,709 (16%) |
| High school | 6,629 (25%) | 6,577 (25%) | 6,696 (25%) | 6,659 (25%) | 6,871 (26%) | 33,432 (25%) |
| Elementary | 12,677 (48%) | 12,718 (48%) | 12,582 (47%) | 12,651 (48%) | 12,279 (46%) | 62,907 (47%) |
| Other | 3,391 (13%) | 3,323 (12%) | 3,320 (12%) | 3,094 (12%) | 2,963 (11%) | 16,091 (12%) |
| Missing | 13 (0%) | 16 (0%) | 14 (0%) | 13 (0%) | 12 (0%) | 68 (0%) |
| **Smoking status** | | | | | | |
| Never | 13,096 (49%) | 13,115 (49%) | 13,046 (49%) | 12,857 (48%) | 12,477 (47%) | 64,591 (48%) |
| Former | 5,300 (20%) | 5,223 (20%) | 5,202 (20%) | 5,164 (19%) | 5,287 (20%) | 26,176 (20%) |
| Current | 8,232 (31%) | 8,285 (31%) | 8,368 (31%) | 8,593 (32%) | 8,852 (33%) | 42,330 (32%) |
| Missing | 14 (0%) | 18 (0%) | 25 (0%) | 27 (0%) | 26 (0%) | 110 (0%) |
| **Alcohol intake** | | | | | | |
| Never | 5,202 (20%) | 5,271 (20%) | 5,261 (20%) | 5,325 (20%) | 5,044 (19%) | 26,103 (20%) |
| Former | 3,681 (14%) | 3,747 (14%) | 3,625 (14%) | 3,563 (13%) | 3,635 (14%) | 18,251 (14%) |
| Current | 17,745 (67%) | 17,614 (66%) | 17,744 (67%) | 17,748 (67%) | 17,952 (67%) | 88,803 (67%) |
| Missing | 14 (0%) | 9 (0%) | 11 (0%) | 5 (0%) | 11 (0%) | 50 (0%) |
| **Physical measures** | | | | | | |
| SBP, mmHg | 126.6 (16.6) | 127.0 (16.6) | 127.4 (16.6) | 127.3 (16.5) | 127.8 (16.6) | 127.2 (16.6) |
| DBP, mmHg | 82.8 (10.2) | 83.0 (10.1) | 83.2 (10.3) | 83.2 (10.2) | 83.4 (10.2) | 83.1 (10.2) |
| BMI, kg/m$^2$ | 29.2 (5.1) | 29.2 (5.1) | 29.1 (5.1) | 29.1 (5.1) | 29.0 (5.0) | 29.1 (5.1) |
| Waist-to-hip Ratio | 0.90 (0.08) | 0.90 (0.08) | 0.90 (0.08) | 0.90 (0.08) | 0.90 (0.08) | 0.90 (0.08) |
| **Laboratory measurements** | | | | | | |
| HDL-C, mmol/L | 1.00 (0.21) | 1.00 (0.21) | 1.00 (0.21) | 1.00 (0.21) | 1.00 (0.21) | 1.00 (0.21) |
| LDL-C, mmol/L | 2.43 (0.78) | 2.45 (0.79) | 2.45 (0.79) | 2.47 (0.79) | 2.49 (0.80) | 2.46 (0.79) |
| Triglycerides, mmol/L | 1.56 (0.65) | 1.57 (0.66) | 1.57 (0.66) | 1.58 (0.66) | 1.59 (0.67) | 1.57 (0.66) |
| HbA1c, % | 6.09 (1.69) | 6.12 (1.73) | 6.10 (1.72) | 6.09 (1.72) | 6.08 (1.70) | 6.10 (1.71) |
| eGFR, ml/min/1.73m$^2$[§] | 101.9 (16.1) | 101.8 (16.0) | 101.6 (16.3) | 101.6 (16.0) | 101.4 (15.8) | 101.7 (16.0) |
| **Prior disease**[*] | | | | | | |
| Coronary heart disease | 345 (1%) | 306 (1%) | 385 (1%) | 396 (1%) | 469 (2%) | 1,901 (1%) |
| Stroke | 259 (1%) | 245 (1%) | 322 (1%) | 308 (1%) | 280 (1%) | 1,414 (1%) |
| Cancer | 303 (1%) | 326 (1%) | 312 (1%) | 320 (1%) | 319 (1%) | 1,580 (1%) |
| Diabetes[†] | 4,903 (18%) | 5,078 (19%) | 5,027 (19%) | 4,895 (18%) | 4,893 (18%) | 24,796 (19%) |
| Other[‡] | 2,202 (8%) | 2,151 (8%) | 2,309 (9%) | 2,245 (8%) | 2,206 (8%) | 11,113 (8%) |

Numbers are n (%) or mean (SD). SBP=systolic blood pressure, DBP=diastolic blood pressure, BMI=body mass index, HDL-C=high density lipoprotein cholesterol, LDL-C=low density lipoprotein cholesterol, HbA1c=glycosylated haemoglobin A1c

[*]Self-reported previous diagnoses unless otherwise stated.

[†]Self-reported previously-diagnosed diabetes or glycosylated haemoglobin ≥6.5%.

[‡]Other diseases include self-reported emphysema, chronic kidney disease, peptic ulcer, liver cirrhosis, and peripheral arterial disease.

[§]Calculated using the 2021 CKD-EPI equation, based on NMR-measured creatinine levels.

### Webtable 5: Baseline characteristics of 133,207 participants aged 35-79 by fifth of PRS by Koyama *et al.*

| Koyama et al/PGS000337/ Japanese and European/64,185 SNPs | Fifth of the PRS distribution | | | | | All (n=133,207) |
|---|---|---|---|---|---|---|
| | 1 (n=26,642) | 2 (n=26,641) | 3 (n=26,641) | 4 (n=26,641) | 5 (n=26,642) | |
| **Age, years** | 51.7 (12.0) | 51.4 (11.9) | 51.2 (11.8) | 51.0 (11.8) | 50.9 (11.6) | 51.2 (11.8) |
| **Resident of Coyoacán** | 10,764 (40%) | 10,244 (38%) | 10,137 (38%) | 10,027 (38%) | 9,843 (37%) | 51,015 (38%) |
| **Ancestry admixture percentage** | | | | | | |
| Indigenous American | 62.6 (18.2) | 65.9 (17.8) | 67.4 (17.7) | 68.6 (17.5) | 69.7 (17.3) | 66.8 (17.9) |
| African | 3.7 (2.7) | 3.5 (2.8) | 3.4 (2.7) | 3.3 (2.8) | 3.3 (3.0) | 3.4 (2.8) |
| East Asian | 1.4 (1.7) | 1.4 (1.6) | 1.4 (1.9) | 1.4 (1.7) | 1.4 (2.1) | 1.4 (1.8) |
| European | 32.3 (16.7) | 29.2 (16.2) | 27.7 (16.0) | 26.6 (15.7) | 25.6 (15.4) | 28.3 (16.2) |
| **Highest attained educational level** | | | | | | |
| University/college | 4,846 (18%) | 4,238 (16%) | 4,102 (15%) | 3,910 (15%) | 3,613 (14%) | 20,709 (16%) |
| High school | 6,753 (25%) | 6,774 (25%) | 6,733 (25%) | 6,641 (25%) | 6,531 (25%) | 33,432 (25%) |
| Elementary | 12,110 (45%) | 12,472 (47%) | 12,567 (47%) | 12,775 (48%) | 12,983 (49%) | 62,907 (47%) |
| Other | 2,921 (11%) | 3,146 (12%) | 3,225 (12%) | 3,296 (12%) | 3,503 (13%) | 16,091 (12%) |
| Missing | 12 (0%) | 11 (0%) | 14 (0%) | 19 (0%) | 12 (0%) | 68 (0%) |
| **Smoking status** | | | | | | |
| Never | 12,947 (49%) | 13,096 (49%) | 12,831 (48%) | 12,915 (49%) | 12,802 (48%) | 64,591 (48%) |
| Former | 5,258 (20%) | 5,191 (20%) | 5,356 (20%) | 5,162 (19%) | 5,209 (20%) | 26,176 (20%) |
| Current | 8,419 (32%) | 8,332 (31%) | 8,429 (32%) | 8,539 (32%) | 8,611 (32%) | 42,330 (32%) |
| Missing | 18 (0%) | 22 (0%) | 25 (0%) | 25 (0%) | 20 (0%) | 110 (0%) |
| **Alcohol intake** | | | | | | |
| Never | 5,023 (19%) | 5,300 (20%) | 5,166 (19%) | 5,293 (20%) | 5,321 (20%) | 26,103 (20%) |
| Former | 3,495 (13%) | 3,583 (13%) | 3,590 (13%) | 3,732 (14%) | 3,851 (14%) | 18,251 (14%) |
| Current | 18,115 (68%) | 17,746 (67%) | 17,876 (67%) | 17,608 (66%) | 17,458 (66%) | 88,803 (67%) |
| Missing | 9 (0%) | 12 (0%) | 9 (0%) | 8 (0%) | 12 (0%) | 50 (0%) |
| **Physical measures** | | | | | | |
| SBP, mmHg | 126.2 (16.2) | 126.9 (16.6) | 127.3 (16.5) | 127.6 (16.8) | 128.2 (16.9) | 127.2 (16.6) |
| DBP, mmHg | 82.5 (10.0) | 82.9 (10.2) | 83.1 (10.2) | 83.3 (10.2) | 83.6 (10.3) | 83.1 (10.2) |
| BMI, kg/m² | 29.0 (5.1) | 29.1 (5.1) | 29.1 (5.0) | 29.2 (5.1) | 29.2 (5.1) | 29.1 (5.1) |
| Waist-to-hip Ratio | 0.90 (0.08) | 0.90 (0.08) | 0.90 (0.08) | 0.90 (0.08) | 0.90 (0.08) | 0.90 (0.08) |
| **Laboratory measurements** | | | | | | |
| HDL-C, mmol/L | 1.01 (0.22) | 1.01 (0.21) | 1.00 (0.21) | 1.00 (0.21) | 0.99 (0.21) | 1.00 (0.21) |
| LDL-C, mmol/L | 2.45 (0.78) | 2.45 (0.78) | 2.46 (0.79) | 2.46 (0.80) | 2.46 (0.80) | 2.46 (0.79) |
| Triglycerides, mmol/L | 1.52 (0.63) | 1.56 (0.65) | 1.58 (0.66) | 1.59 (0.66) | 1.62 (0.68) | 1.57 (0.66) |
| HbA1c, % | 5.98 (1.58) | 6.06 (1.66) | 6.10 (1.72) | 6.13 (1.75) | 6.22 (1.83) | 6.10 (1.71) |
| eGFR, ml/min/1.73m²[§] | 101.2 (15.9) | 101.5 (16.0) | 101.7 (16.1) | 101.9 (16.1) | 102.0 (16.0) | 101.7 (16.0) |
| **Prior disease**[*] | | | | | | |
| Coronary heart disease | 325 (1%) | 330 (1%) | 357 (1%) | 396 (1%) | 493 (2%) | 1,901 (1%) |
| Stroke | 252 (1%) | 279 (1%) | 284 (1%) | 263 (1%) | 336 (1%) | 1,414 (1%) |
| Cancer | 336 (1%) | 357 (1%) | 285 (1%) | 318 (1%) | 284 (1%) | 1,580 (1%) |
| Diabetes[†] | 4,208 (16%) | 4,731 (18%) | 4,980 (19%) | 5,164 (19%) | 5,713 (21%) | 24,796 (19%) |
| Other[‡] | 2,392 (9%) | 2,242 (8%) | 2,237 (8%) | 2,151 (8%) | 2,091 (8%) | 11,113 (8%) |

Numbers are n (%) or mean (SD). SBP=systolic blood pressure, DBP=diastolic blood pressure, BMI=body mass index, HDL-C=high density lipoprotein cholesterol, LDL-C=low density lipoprotein cholesterol, HbA1c=glycosylated haemoglobin A1c

[*]Self-reported previous diagnoses unless otherwise stated.

[†]Self-reported previously-diagnosed diabetes or glycosylated haemoglobin ≥6.5%.

[‡]Other diseases include self-reported emphysema, chronic kidney disease, peptic ulcer, liver cirrhosis, and peripheral arterial disease.

[§]Calculated using the 2021 CKD-EPI equation, based on NMR-measured creatinine levels.

### Webtable 6: Baseline characteristics of 133,207 participants aged 35-79 by fifth of PRS by Tcheandjieu *et al.*

| **Tcheandjieu et al/PGS003446/ Multi-ancestry/485,464 SNPs** | **Fifth of the PRS distribution** | | | | | **All (n=133,207)** |
|---|---|---|---|---|---|---|
| | **1 (n=26,642)** | **2 (n=26,641)** | **3 (n=26,641)** | **4 (n=26,641)** | **5 (n=26,642)** | |
| **Age, years** | 51.0 (11.8) | 51.0 (11.8) | 51.0 (11.8) | 51.3 (11.8) | 51.9 (12.0) | 51.2 (11.8) |
| **Resident of Coyoacán** | 8,908 (33%) | 9,590 (36%) | 10,094 (38%) | 10,650 (40%) | 11,773 (44%) | 51,015 (38%) |
| **Ancestry admixture percentage** | | | | | | |
| Indigenous American | 80.2 (14.4) | 73.0 (15.4) | 67.0 (15.5) | 61.3 (15.2) | 52.7 (15.4) | 66.8 (17.9) |
| African | 2.4 (2.9) | 3.0 (2.8) | 3.5 (2.6) | 3.9 (2.6) | 4.4 (2.6) | 3.4 (2.8) |
| East Asian | 1.3 (2.4) | 1.4 (1.9) | 1.5 (2.0) | 1.5 (1.3) | 1.5 (1.3) | 1.4 (1.8) |
| European | 16.1 (12.2) | 22.6 (13.5) | 28.0 (13.8) | 33.4 (13.9) | 41.3 (14.8) | 28.3 (16.2) |
| **Highest attained educational level** | | | | | | |
| University/college | 3,238 (12%) | 3,697 (14%) | 4,117 (15%) | 4,400 (17%) | 5,257 (20%) | 20,709 (16%) |
| High school | 6,051 (23%) | 6,469 (24%) | 6,823 (26%) | 7,013 (26%) | 7,076 (27%) | 33,432 (25%) |
| Elementary | 13,386 (50%) | 13,054 (49%) | 12,564 (47%) | 12,299 (46%) | 11,604 (44%) | 62,907 (47%) |
| Other | 3,951 (15%) | 3,411 (13%) | 3,121 (12%) | 2,915 (11%) | 2,693 (10%) | 16,091 (12%) |
| Missing | 16 (0%) | 10 (0%) | 16 (0%) | 14 (0%) | 12 (0%) | 68 (0%) |
| **Smoking status** | | | | | | |
| Never | 14,516 (55%) | 13,488 (51%) | 12,956 (49%) | 12,215 (46%) | 11,416 (43%) | 64,591 (48%) |
| Former | 5,093 (19%) | 5,151 (19%) | 5,305 (20%) | 5,260 (20%) | 5,367 (20%) | 26,176 (20%) |
| Current | 7,011 (26%) | 7,984 (30%) | 8,359 (31%) | 9,145 (34%) | 9,831 (37%) | 42,330 (32%) |
| Missing | 22 (0%) | 18 (0%) | 21 (0%) | 21 (0%) | 28 (0%) | 110 (0%) |
| **Alcohol intake** | | | | | | |
| Never | 5,649 (21%) | 5,233 (20%) | 5,163 (19%) | 5,127 (19%) | 4,931 (19%) | 26,103 (20%) |
| Former | 3,658 (14%) | 3,712 (14%) | 3,668 (14%) | 3,548 (13%) | 3,665 (14%) | 18,251 (14%) |
| Current | 17,330 (65%) | 17,685 (66%) | 17,799 (67%) | 17,955 (67%) | 18,034 (68%) | 88,803 (67%) |
| Missing | 5 (0%) | 11 (0%) | 11 (0%) | 11 (0%) | 12 (0%) | 50 (0%) |
| **Physical measures** | | | | | | |
| SBP, mmHg | 126.2 (16.2) | 126.8 (16.4) | 127.3 (16.6) | 127.6 (16.7) | 128.3 (17.0) | 127.2 (16.6) |
| DBP, mmHg | 82.5 (9.9) | 82.9 (10.1) | 83.2 (10.2) | 83.3 (10.3) | 83.6 (10.4) | 83.1 (10.2) |
| BMI, kg/m² | 29.0 (4.9) | 29.1 (5.0) | 29.2 (5.0) | 29.2 (5.1) | 29.1 (5.3) | 29.1 (5.1) |
| Waist-to-hip Ratio | 0.90 (0.07) | 0.90 (0.08) | 0.90 (0.08) | 0.90 (0.08) | 0.90 (0.08) | 0.90 (0.08) |
| **Laboratory measurements** | | | | | | |
| HDL-C, mmol/L | 1.00 (0.21) | 1.00 (0.21) | 1.00 (0.21) | 1.00 (0.21) | 1.00 (0.22) | 1.00 (0.21) |
| LDL-C, mmol/L | 2.39 (0.77) | 2.43 (0.78) | 2.45 (0.78) | 2.48 (0.78) | 2.53 (0.82) | 2.46 (0.79) |
| Triglycerides, mmol/L | 1.57 (0.64) | 1.57 (0.66) | 1.58 (0.66) | 1.57 (0.65) | 1.58 (0.67) | 1.57 (0.66) |
| HbA1c, % | 6.16 (1.77) | 6.12 (1.72) | 6.10 (1.71) | 6.08 (1.71) | 6.04 (1.66) | 6.10 (1.71) |
| eGFR, ml/min/1.73m²[§] | 102.9 (15.6) | 102.4 (15.9) | 101.8 (16.0) | 101.2 (16.1) | 99.9 (16.4) | 101.7 (16.0) |
| **Prior disease[*]** | | | | | | |
| Coronary heart disease | 249 (1%) | 275 (1%) | 355 (1%) | 450 (2%) | 572 (2%) | 1,901 (1%) |
| Stroke | 267 (1%) | 289 (1%) | 269 (1%) | 276 (1%) | 313 (1%) | 1,414 (1%) |
| Cancer | 278 (1%) | 295 (1%) | 282 (1%) | 335 (1%) | 390 (1%) | 1,580 (1%) |
| Diabetes[†] | 5,066 (19%) | 5,053 (19%) | 5,033 (19%) | 4,816 (18%) | 4,828 (18%) | 24,796 (19%) |
| Other[‡] | 1,880 ( 7%) | 2,106 ( 8%) | 2,180 ( 8%) | 2,346 ( 9%) | 2,601 (10%) | 11,113 (8%) |

Numbers are n (%) or mean (SD). SBP=systolic blood pressure, DBP=diastolic blood pressure, BMI=body mass index, HDL-C=high density lipoprotein cholesterol, LDL-C=low density lipoprotein cholesterol, HbA1c=glycosylated haemoglobin A1c

[*]Self-reported previous diagnoses unless otherwise stated.

[†]Self-reported previously-diagnosed diabetes or glycosylated haemoglobin ≥6.5%.

[‡]Other diseases include self-reported emphysema, chronic kidney disease, peptic ulcer, liver cirrhosis, and peripheral arterial disease.

[§]Calculated using the 2021 CKD-EPI equation, based on NMR-measured creatinine levels.

### Webtable 7: Baseline characteristics of 133,207 participants aged 35-79 by fifth of PRS by Tamlander *et al.*

| Tamlander et al/ PGS001780/ European/1,087,958 SNPs | Fifth of the PRS distribution | | | | | All (n=133,207) |
|---|---|---|---|---|---|---|
| | 1 (n=26,642) | 2 (n=26,641) | 3 (n=26,641) | 4 (n=26,641) | 5 (n=26,642) | |
| **Age, years** | 52.0 (12.0) | 51.4 (11.9) | 51.0 (11.8) | 51.0 (11.8) | 50.7 (11.7) | 51.2 (11.8) |
| **Resident of Coyoacán** | 11,363 (43%) | 10,657 (40%) | 10,099 (38%) | 9,634 (36%) | 9,262 (35%) | 51,015 (38%) |
| **Ancestry admixture percentage** | | | | | | |
| Indigenous American | 56.7 (16.9) | 63.7 (17.0) | 67.6 (17.0) | 71.1 (16.8) | 75.1 (15.9) | 66.8 (17.9) |
| African | 4.3 (3.1) | 3.8 (2.8) | 3.4 (2.7) | 3.1 (2.6) | 2.6 (2.4) | 3.4 (2.8) |
| East Asian | 1.5 (1.5) | 1.4 (1.7) | 1.4 (1.8) | 1.4 (2.0) | 1.4 (2.1) | 1.4 (1.8) |
| European | 37.5 (15.8) | 31.1 (15.5) | 27.5 (15.3) | 24.5 (14.9) | 20.9 (14.1) | 28.3 (16.2) |
| **Highest attained educational level** | | | | | | |
| University/college | 5,318 (20%) | 4,523 (17%) | 4,002 (15%) | 3,605 (14%) | 3,261 (12%) | 20,709 (16%) |
| High school | 7,075 (27%) | 6,861 (26%) | 6,787 (25%) | 6,478 (24%) | 6,231 (23%) | 33,432 (25%) |
| Elementary | 11,578 (43%) | 12,270 (46%) | 12,655 (48%) | 13,087 (49%) | 13,317 (50%) | 62,907 (47%) |
| Other | 2,654 (10%) | 2,975 (11%) | 3,190 (12%) | 3,459 (13%) | 3,813 (14%) | 16,091 (12%) |
| Missing | 17 (0%) | 12 (0%) | 7 (0%) | 12 (0%) | 20 (0%) | 68 (0%) |
| **Smoking status** | | | | | | |
| Never | 12,389 (47%) | 12,766 (48%) | 12,950 (49%) | 13,056 (49%) | 13,430 (50%) | 64,591 (48%) |
| Former | 5,382 (20%) | 5,189 (19%) | 5,207 (20%) | 5,305 (20%) | 5,093 (19%) | 26,176 (20%) |
| Current | 8,846 (33%) | 8,661 (33%) | 8,464 (32%) | 8,259 (31%) | 8,100 (30%) | 42,330 (32%) |
| Missing | 25 (0%) | 25 (0%) | 20 (0%) | 21 (0%) | 19 (0%) | 110 (0%) |
| **Alcohol intake** | | | | | | |
| Never | 4,887 (18%) | 5,091 (19%) | 5,212 (20%) | 5,389 (20%) | 5,524 (21%) | 26,103 (20%) |
| Former | 3,494 (13%) | 3,527 (13%) | 3,621 (14%) | 3,769 (14%) | 3,840 (14%) | 18,251 (14%) |
| Current | 18,249 (69%) | 18,012 (68%) | 17,800 (67%) | 17,477 (66%) | 17,265 (65%) | 88,803 (67%) |
| Missing | 12 (0%) | 11 (0%) | 8 (0%) | 6 (0%) | 13 (0%) | 50 (0%) |
| **Physical measures** | | | | | | |
| SBP, mmHg | 126.7 (16.4) | 127.1 (16.4) | 127.0 (16.6) | 127.6 (16.7) | 127.9 (16.8) | 127.2 (16.6) |
| DBP, mmHg | 82.9 (10.1) | 83.0 (10.2) | 83.0 (10.2) | 83.3 (10.2) | 83.4 (10.2) | 83.1 (10.2) |
| BMI, kg/m² | 29.0 (5.2) | 29.1 (5.1) | 29.1 (5.1) | 29.2 (5.0) | 29.2 (5.0) | 29.1 (5.1) |
| Waist-to-hip Ratio | 0.90 (0.08) | 0.90 (0.08) | 0.90 (0.08) | 0.90 (0.08) | 0.91 (0.07) | 0.90 (0.08) |
| **Laboratory measurements** | | | | | | |
| HDL-C, mmol/L | 1.01 (0.22) | 1.00 (0.21) | 1.00 (0.21) | 1.00 (0.21) | 0.99 (0.21) | 1.00 (0.21) |
| LDL-C, mmol/L | 2.47 (0.78) | 2.47 (0.78) | 2.46 (0.79) | 2.45 (0.80) | 2.45 (0.80) | 2.46 (0.79) |
| Triglycerides, mmol/L | 1.53 (0.64) | 1.56 (0.65) | 1.57 (0.66) | 1.59 (0.66) | 1.63 (0.68) | 1.57 (0.66) |
| HbA1c, % | 5.95 (1.55) | 6.03 (1.63) | 6.10 (1.72) | 6.15 (1.76) | 6.26 (1.87) | 6.10 (1.71) |
| eGFR, ml/min/1.73m²[§] | 100.5 (16.0) | 101.4 (16.0) | 101.8 (16.0) | 102.1 (16.1) | 102.4 (16.0) | 101.7 (16.0) |
| **Prior disease**[*] | | | | | | |
| Coronary heart disease | 379 (1%) | 367 (1%) | 357 (1%) | 384 (1%) | 414 (2%) | 1,901 (1%) |
| Stroke | 255 (1%) | 279 (1%) | 300 (1%) | 278 (1%) | 302 (1%) | 1,414 (1%) |
| Cancer | 367 (1%) | 329 (1%) | 301 (1%) | 308 (1%) | 275 (1%) | 1,580 (1%) |
| Diabetes[†] | 4,096 (15%) | 4,624 (17%) | 4,952 (19%) | 5,287 (20%) | 5,837 (22%) | 24,796 (19%) |
| Other[‡] | 2,504 (9%) | 2,352 (9%) | 2,233 (8%) | 2,135 (8%) | 1,889 (7%) | 11,113 (8%) |

Numbers are *n* (%) or mean (SD). SBP=systolic blood pressure, DBP=diastolic blood pressure, BMI=body mass index, HDL-C=high density lipoprotein cholesterol, LDL-C=low density lipoprotein cholesterol, HbA1c=glycosylated haemoglobin A1c

[*]Self-reported previous diagnoses unless otherwise stated.

[†]Self-reported previously-diagnosed diabetes or glycosylated haemoglobin ≥6.5%.

[‡]Other diseases include self-reported emphysema, chronic kidney disease, peptic ulcer, liver cirrhosis, and peripheral arterial disease.

[§]Calculated using the 2021 CKD-EPI equation, based on NMR-measured creatinine levels.

### Webtable 8: Baseline characteristics of 133,207 participants aged 35-79 by fifth of PRS by Patel *et al.*

| Patel et al/PGS003725/ Multi-ancestry/1,273,824 SNPs | Fifth of the PRS distribution | | | | | All (n=133,207) |
|---|---|---|---|---|---|---|
| | 1 (n=26,642) | 2 (n=26,641) | 3 (n=26,641) | 4 (n=26,641) | 5 (n=26,642) | |
| **Age, years** | 51.5 (12.0) | 51.4 (11.9) | 51.3 (11.9) | 51.1 (11.7) | 50.9 (11.6) | 51.2 (11.8) |
| **Resident of Coyoacán** | 9,980 (37%) | 10,079 (38%) | 10,204 (38%) | 10,162 (38%) | 10,590 (40%) | 51,015 (38%) |
| **Ancestry admixture percentage** | | | | | | |
| Indigenous American | 70.4 (17.7) | 68.8 (17.7) | 67.5 (17.7) | 65.6 (17.7) | 62.0 (17.4) | 66.8 (17.9) |
| African | 2.9 (2.3) | 3.2 (2.5) | 3.4 (2.7) | 3.6 (2.8) | 4.1 (3.4) | 3.4 (2.8) |
| East Asian | 1.3 (1.5) | 1.4 (1.7) | 1.4 (1.9) | 1.4 (1.7) | 1.5 (2.3) | 1.4 (1.8) |
| European | 25.4 (16.0) | 26.7 (16.1) | 27.7 (16.0) | 29.4 (16.0) | 32.3 (15.9) | 28.3 (16.2) |
| **Highest attained educational level** | | | | | | |
| University/college | 4,224 (16%) | 4,076 (15%) | 4,110 (15%) | 4,080 (15%) | 4,219 (16%) | 20,709 (16%) |
| High school | 6,441 (24%) | 6,761 (25%) | 6,658 (25%) | 6,691 (25%) | 6,881 (26%) | 33,432 (25%) |
| Elementary | 12,709 (48%) | 12,498 (47%) | 12,576 (47%) | 12,642 (47%) | 12,482 (47%) | 62,907 (47%) |
| Other | 3,256 (12%) | 3,292 (12%) | 3,278 (12%) | 3,218 (12%) | 3,047 (11%) | 16,091 (12%) |
| Missing | 12 (0%) | 14 (0%) | 19 (0%) | 10 (0%) | 13 (0%) | 68 (0%) |
| **Smoking status** | | | | | | |
| Never | 13,720 (52%) | 13,227 (50%) | 12,869 (48%) | 12,634 (47%) | 12,141 (46%) | 64,591 (48%) |
| Former | 5,166 (19%) | 5,245 (20%) | 5,302 (20%) | 5,224 (20%) | 5,239 (20%) | 26,176 (20%) |
| Current | 7,732 (29%) | 8,149 (31%) | 8,450 (32%) | 8,758 (33%) | 9,241 (35%) | 42,330 (32%) |
| Missing | 24 (0%) | 20 (0%) | 20 (0%) | 25 (0%) | 21 (0%) | 110 (0%) |
| **Alcohol intake** | | | | | | |
| Never | 5,127 (19%) | 5,196 (20%) | 5,336 (20%) | 5,331 (20%) | 5,113 (19%) | 26,103 (20%) |
| Former | 3,616 (14%) | 3,599 (14%) | 3,608 (14%) | 3,688 (14%) | 3,740 (14%) | 18,251 (14%) |
| Current | 17,890 (67%) | 17,834 (67%) | 17,690 (66%) | 17,611 (66%) | 17,778 (67%) | 88,803 (67%) |
| Missing | 9 (0%) | 12 (0%) | 7 (0%) | 11 (0%) | 11 (0%) | 50 (0%) |
| **Physical measures** | | | | | | |
| SBP, mmHg | 125.8 (16.1) | 126.6 (16.4) | 127.2 (16.4) | 127.9 (16.9) | 128.6 (17.0) | 127.2 (16.6) |
| DBP, mmHg | 82.3 (10.1) | 82.7 (10.1) | 83.2 (10.1) | 83.4 (10.3) | 83.9 (10.4) | 83.1 (10.2) |
| BMI, kg/m$^2$ | 28.9 (5.0) | 29.1 (5.0) | 29.1 (5.1) | 29.2 (5.1) | 29.3 (5.2) | 29.1 (5.1) |
| Waist-to-hip Ratio | 0.90 (0.08) | 0.90 (0.08) | 0.90 (0.08) | 0.90 (0.08) | 0.90 (0.08) | 0.90 (0.08) |
| **Laboratory measurements** | | | | | | |
| HDL-C, mmol/L | 1.01 (0.22) | 1.01 (0.21) | 1.00 (0.21) | 1.00 (0.21) | 0.99 (0.21) | 1.00 (0.21) |
| LDL-C, mmol/L | 2.40 (0.76) | 2.44 (0.78) | 2.46 (0.79) | 2.48 (0.80) | 2.51 (0.82) | 2.46 (0.79) |
| Triglycerides, mmol/L | 1.50 (0.61) | 1.55 (0.64) | 1.58 (0.66) | 1.60 (0.68) | 1.63 (0.70) | 1.57 (0.66) |
| HbA1c, % | 5.98 (1.58) | 6.05 (1.64) | 6.09 (1.69) | 6.16 (1.78) | 6.22 (1.84) | 6.10 (1.71) |
| eGFR, ml/min/1.73m$^2$[§] | 102.0 (15.7) | 101.9 (15.8) | 101.5 (16.2) | 101.6 (16.1) | 101.3 (16.4) | 101.7 (16.0) |
| **Prior disease**[*] | | | | | | |
| Coronary heart disease | 285 (1%) | 324 (1%) | 332 (1%) | 389 (1%) | 571 (2%) | 1,901 (1%) |
| Stroke | 233 (1%) | 259 (1%) | 310 (1%) | 280 (1%) | 332 (1%) | 1,414 (1%) |
| Cancer | 312 (1%) | 321 (1%) | 316 (1%) | 319 (1%) | 312 (1%) | 1,580 (1%) |
| Diabetes[†] | 4,195 (16%) | 4,583 (17%) | 4,922 (18%) | 5,353 (20%) | 5,743 (22%) | 24,796 (19%) |
| Other[‡] | 2,253 (8%) | 2,208 (8%) | 2,193 (8%) | 2,269 (9%) | 2,190 (8%) | 11,113 (8%) |

Numbers are *n* (%) or mean (SD). SBP=systolic blood pressure, DBP=diastolic blood pressure, BMI=body mass index, HDL-C=high density lipoprotein cholesterol, LDL-C=low density lipoprotein cholesterol, HbA1c=glycosylated haemoglobin A1c

[*]Self-reported previous diagnoses unless otherwise stated.

[†]Self-reported previously-diagnosed diabetes or glycosylated haemoglobin ≥6.5%.

[‡]Other diseases include self-reported emphysema, chronic kidney disease, peptic ulcer, liver cirrhosis, and peripheral arterial disease.

[§]Calculated using the 2021 CKD-EPI equation, based on NMR-measured creatinine levels.

### Webtable 9: Baseline characteristics of 133,207 participants aged 35-79 by fifth of PRS by Inouye *et al.*

| **Inouye et al/PGS000018/ European/1,720,068 SNPs** | **Fifth of the PRS distribution** | | | | | **All (n=133,207)** |
|---|---|---|---|---|---|---|
| | **1 (n=26,642)** | **2 (n=26,641)** | **3 (n=26,641)** | **4 (n=26,641)** | **5 (n=26,642)** | |
| **Age, years** | 52.4 (12.1) | 51.4 (11.8) | 51.0 (11.8) | 50.8 (11.8) | 50.7 (11.7) | 51.2 (11.8) |
| **Resident of Coyoacán** | 11,967 (45%) | 10,593 (40%) | 10,046 (38%) | 9,484 (36%) | 8,925 (33%) | 51,015 (38%) |
| **Ancestry admixture percentage** | | | | | | |
| Indigenous American | 51.8 (15.1) | 61.7 (15.2) | 67.8 (15.6) | 73.3 (15.3) | 79.6 (14.3) | 66.8 (17.9) |
| African | 4.6 (2.8) | 4.0 (2.8) | 3.5 (2.8) | 2.9 (2.6) | 2.3 (2.3) | 3.4 (2.8) |
| East Asian | 1.5 (1.5) | 1.5 (1.6) | 1.5 (1.9) | 1.4 (1.8) | 1.3 (2.2) | 1.4 (1.8) |
| European | 42.1 (14.4) | 32.8 (13.8) | 27.3 (13.9) | 22.4 (13.5) | 16.9 (12.4) | 28.3 (16.2) |
| **Highest attained educational level** | | | | | | |
| University/college | 5,825 (22%) | 4,492 (17%) | 4,008 (15%) | 3,480 (13%) | 2,904 (11%) | 20,709 (16%) |
| High school | 7,149 (27%) | 7,077 (27%) | 6,792 (26%) | 6,366 (24%) | 6,048 (23%) | 33,432 (25%) |
| Elementary | 11,220 (42%) | 12,188 (46%) | 12,757 (48%) | 13,209 (50%) | 13,533 (51%) | 62,907 (47%) |
| Other | 2,440 ( 9%) | 2,872 (11%) | 3,072 (12%) | 3,568 (13%) | 4,139 (16%) | 16,091 (12%) |
| Missing | 8 (0%) | 12 (0%) | 12 (0%) | 18 (0%) | 18 (0%) | 68 (0%) |
| **Smoking status** | | | | | | |
| Never | 11,982 (45%) | 12,541 (47%) | 12,966 (49%) | 13,351 (50%) | 13,751 (52%) | 64,591 (48%) |
| Former | 5,364 (20%) | 5,314 (20%) | 5,198 (20%) | 5,200 (20%) | 5,100 (19%) | 26,176 (20%) |
| Current | 9,268 (35%) | 8,771 (33%) | 8,454 (32%) | 8,067 (30%) | 7,770 (29%) | 42,330 (32%) |
| Missing | 28 (0%) | 15 (0%) | 23 (0%) | 23 (0%) | 21 (0%) | 110 (0%) |
| **Alcohol intake** | | | | | | |
| Never | 4,730 (18%) | 5,224 (20%) | 5,215 (20%) | 5,404 (20%) | 5,530 (21%) | 26,103 (20%) |
| Former | 3,379 (13%) | 3,567 (13%) | 3,661 (14%) | 3,708 (14%) | 3,936 (15%) | 18,251 (14%) |
| Current | 18,520 (70%) | 17,840 (67%) | 17,757 (67%) | 17,519 (66%) | 17,167 (64%) | 88,803 (67%) |
| Missing | 13 (0%) | 10 (0%) | 8 (0%) | 10 (0%) | 9 (0%) | 50 (0%) |
| **Physical measures** | | | | | | |
| SBP, mmHg | 126.9 (16.5) | 127.1 (16.4) | 127.0 (16.6) | 127.5 (16.8) | 127.7 (16.7) | 127.2 (16.6) |
| DBP, mmHg | 83.0 (10.1) | 83.1 (10.2) | 83.1 (10.2) | 83.2 (10.2) | 83.3 (10.2) | 83.1 (10.2) |
| BMI, kg/m² | 28.9 (5.2) | 29.1 (5.1) | 29.1 (5.1) | 29.2 (5.0) | 29.2 (4.9) | 29.1 (5.1) |
| Waist-to-hip Ratio | 0.90 (0.08) | 0.90 (0.08) | 0.90 (0.08) | 0.90 (0.08) | 0.91 (0.07) | 0.90 (0.08) |
| **Laboratory measurements** | | | | | | |
| HDL-C, mmol/L | 1.01 (0.22) | 1.00 (0.21) | 1.00 (0.21) | 1.00 (0.21) | 0.99 (0.21) | 1.00 (0.21) |
| LDL-C, mmol/L | 2.50 (0.78) | 2.48 (0.80) | 2.45 (0.79) | 2.44 (0.78) | 2.42 (0.79) | 2.46 (0.79) |
| Triglycerides, mmol/L | 1.52 (0.63) | 1.56 (0.66) | 1.57 (0.66) | 1.60 (0.67) | 1.62 (0.67) | 1.57 (0.66) |
| HbA1c, % | 5.90 (1.48) | 6.04 (1.66) | 6.10 (1.72) | 6.16 (1.76) | 6.28 (1.89) | 6.10 (1.71) |
| eGFR, ml/min/1.73m²[§] | 99.8 (16.1) | 101.2 (16.0) | 102.0 (15.9) | 102.3 (16.1) | 103.0 (15.9) | 101.7 (16.0) |
| **Prior disease**[*] | | | | | | |
| Coronary heart disease | 432 (2%) | 382 (1%) | 364 (1%) | 332 (1%) | 391 (1%) | 1,901 (1%) |
| Stroke | 255 (1%) | 267 (1%) | 298 (1%) | 285 (1%) | 309 (1%) | 1,414 (1%) |
| Cancer | 384 (1%) | 348 (1%) | 318 (1%) | 263 (1%) | 267 (1%) | 1,580 (1%) |
| Diabetes[†] | 3,898 (15%) | 4,589 (17%) | 4,959 (19%) | 5,418 (20%) | 5,932 (22%) | 24,796 (19%) |
| Other[‡] | 2,664 (10%) | 2,319 ( 9%) | 2,231 ( 8%) | 2,029 ( 8%) | 1,870 ( 7%) | 11,113 (8%) |

Numbers are *n* (%) or mean (SD). SBP=systolic blood pressure, DBP=diastolic blood pressure, BMI=body mass index, HDL-C=high density lipoprotein cholesterol, LDL-C=low density lipoprotein cholesterol, HbA1c=glycosylated haemoglobin A1c

[*]Self-reported previous diagnoses unless otherwise stated.

[†]Self-reported previously-diagnosed diabetes or glycosylated haemoglobin ≥6.5%.

[‡]Other diseases include self-reported emphysema, chronic kidney disease, peptic ulcer, liver cirrhosis, and peripheral arterial disease.

[§]Calculated using the 2021 CKD-EPI equation, based on NMR-measured creatinine levels.

### Webtable 10: Baseline characteristics of 133,207 participants aged 35-79 by fifth of PRS by Khera *et al.*

| **Khera et al/PGS000013/ European/6,472,620 SNPs** | **Fifth of the PRS distribution** | | | | | **All (n=133,207)** |
|---|---|---|---|---|---|---|
| | **1 (n=26,642)** | **2 (n=26,641)** | **3 (n=26,641)** | **4 (n=26,641)** | **5 (n=26,642)** | |
| **Age, years** | 51.4 (11.9) | 51.2 (11.8) | 51.3 (11.8) | 51.1 (11.8) | 51.2 (11.8) | 51.2 (11.8) |
| **Resident of Coyoacán** | 10,270 (39%) | 10,129 (38%) | 10,096 (38%) | 10,227 (38%) | 10,293 (39%) | 51,015 (38%) |
| **Ancestry admixture percentage** | | | | | | |
| Indigenous American | 66.1 (17.7) | 67.9 (17.9) | 67.9 (17.8) | 67.2 (17.9) | 65.1 (17.8) | 66.8 (17.9) |
| African | 3.7 (3.4) | 3.3 (2.7) | 3.3 (2.6) | 3.4 (2.6) | 3.5 (2.6) | 3.4 (2.8) |
| East Asian | 1.4 (1.4) | 1.4 (1.6) | 1.4 (1.6) | 1.4 (1.9) | 1.5 (2.4) | 1.4 (1.8) |
| European | 28.7 (16.0) | 27.4 (16.2) | 27.4 (16.1) | 28.0 (16.2) | 29.9 (16.2) | 28.3 (16.2) |
| **Highest attained educational level** | | | | | | |
| University/college | 4,272 (16%) | 4,068 (15%) | 4,091 (15%) | 4,108 (15%) | 4,170 (16%) | 20,709 (16%) |
| High school | 6,707 (25%) | 6,746 (25%) | 6,642 (25%) | 6,683 (25%) | 6,654 (25%) | 33,432 (25%) |
| Elementary | 12,395 (47%) | 12,676 (48%) | 12,604 (47%) | 12,657 (48%) | 12,575 (47%) | 62,907 (47%) |
| Other | 3,251 (12%) | 3,143 (12%) | 3,291 (12%) | 3,176 (12%) | 3,230 (12%) | 16,091 (12%) |
| Missing | 17 (0%) | 8 (0%) | 13 (0%) | 17 (0%) | 13 (0%) | 68 (0%) |
| **Smoking status** | | | | | | |
| Never | 13,064 (49%) | 13,057 (49%) | 13,048 (49%) | 12,703 (48%) | 12,719 (48%) | 64,591 (48%) |
| Former | 5,294 (20%) | 5,132 (19%) | 5,187 (19%) | 5,256 (20%) | 5,307 (20%) | 26,176 (20%) |
| Current | 8,264 (31%) | 8,437 (32%) | 8,382 (31%) | 8,654 (33%) | 8,593 (32%) | 42,330 (32%) |
| Missing | 20 (0%) | 15 (0%) | 24 (0%) | 28 (0%) | 23 (0%) | 110 (0%) |
| **Alcohol intake** | | | | | | |
| Never | 5,175 (19%) | 5,122 (19%) | 5,258 (20%) | 5,300 (20%) | 5,248 (20%) | 26,103 (20%) |
| Former | 3,641 (14%) | 3,617 (14%) | 3,635 (14%) | 3,659 (14%) | 3,699 (14%) | 18,251 (14%) |
| Current | 17,815 (67%) | 17,896 (67%) | 17,737 (67%) | 17,670 (66%) | 17,685 (66%) | 88,803 (67%) |
| Missing | 11 (0%) | 6 (0%) | 11 (0%) | 12 (0%) | 10 (0%) | 50 (0%) |
| **Physical measures** | | | | | | |
| SBP, mmHg | 126.7 (16.6) | 126.9 (16.3) | 127.3 (16.6) | 127.3 (16.7) | 128.0 (16.8) | 127.2 (16.6) |
| DBP, mmHg | 82.9 (10.1) | 83.0 (10.2) | 83.1 (10.2) | 83.2 (10.2) | 83.4 (10.3) | 83.1 (10.2) |
| BMI, kg/m² | 29.2 (5.2) | 29.1 (5.0) | 29.1 (5.0) | 29.1 (5.1) | 29.0 (5.1) | 29.1 (5.1) |
| Waist-to-hip Ratio | 0.90 (0.08) | 0.90 (0.08) | 0.90 (0.08) | 0.90 (0.08) | 0.90 (0.08) | 0.90 (0.08) |
| **Laboratory measurements** | | | | | | |
| HDL-C, mmol/L | 1.00 (0.21) | 1.00 (0.21) | 1.00 (0.21) | 1.00 (0.21) | 1.00 (0.21) | 1.00 (0.21) |
| LDL-C, mmol/L | 2.42 (0.77) | 2.44 (0.79) | 2.45 (0.79) | 2.46 (0.80) | 2.51 (0.80) | 2.46 (0.79) |
| Triglycerides, mmol/L | 1.55 (0.64) | 1.57 (0.65) | 1.58 (0.67) | 1.58 (0.66) | 1.59 (0.67) | 1.57 (0.66) |
| HbA1c, % | 6.05 (1.66) | 6.09 (1.70) | 6.10 (1.71) | 6.11 (1.73) | 6.13 (1.76) | 6.10 (1.71) |
| eGFR, ml/min/1.73m²[§] | 101.5 (15.9) | 101.8 (16.2) | 101.6 (16.4) | 101.9 (15.8) | 101.5 (15.9) | 101.7 (16.0) |
| **Prior disease**[*] | | | | | | |
| Coronary heart disease | 314 (1%) | 334 (1%) | 328 (1%) | 414 (2%) | 511 (2%) | 1,901 (1%) |
| Stroke | 261 (1%) | 269 (1%) | 302 (1%) | 284 (1%) | 298 (1%) | 1,414 (1%) |
| Cancer | 318 (1%) | 326 (1%) | 314 (1%) | 304 (1%) | 318 (1%) | 1,580 (1%) |
| Diabetes[†] | 4,600 (17%) | 4,969 (19%) | 4,966 (19%) | 5,037 (19%) | 5,224 (20%) | 24,796 (19%) |
| Other[‡] | 2,308 (9%) | 2,195 (8%) | 2,200 (8%) | 2,202 (8%) | 2,208 (8%) | 11,113 (8%) |

Numbers are n (%) or mean (SD). SBP=systolic blood pressure, DBP=diastolic blood pressure, BMI=body mass index, HDL-C=high density lipoprotein cholesterol, LDL-C=low density lipoprotein cholesterol, HbA1c=glycosylated haemoglobin A1c

[*]Self-reported previous diagnoses unless otherwise stated.

[†]Self-reported previously-diagnosed diabetes or glycosylated haemoglobin ≥6.5%.

[‡]Other diseases include self-reported emphysema, chronic kidney disease, peptic ulcer, liver cirrhosis, and peripheral arterial disease.

[§]Calculated using the 2021 CKD-EPI equation, based on NMR-measured creatinine levels.

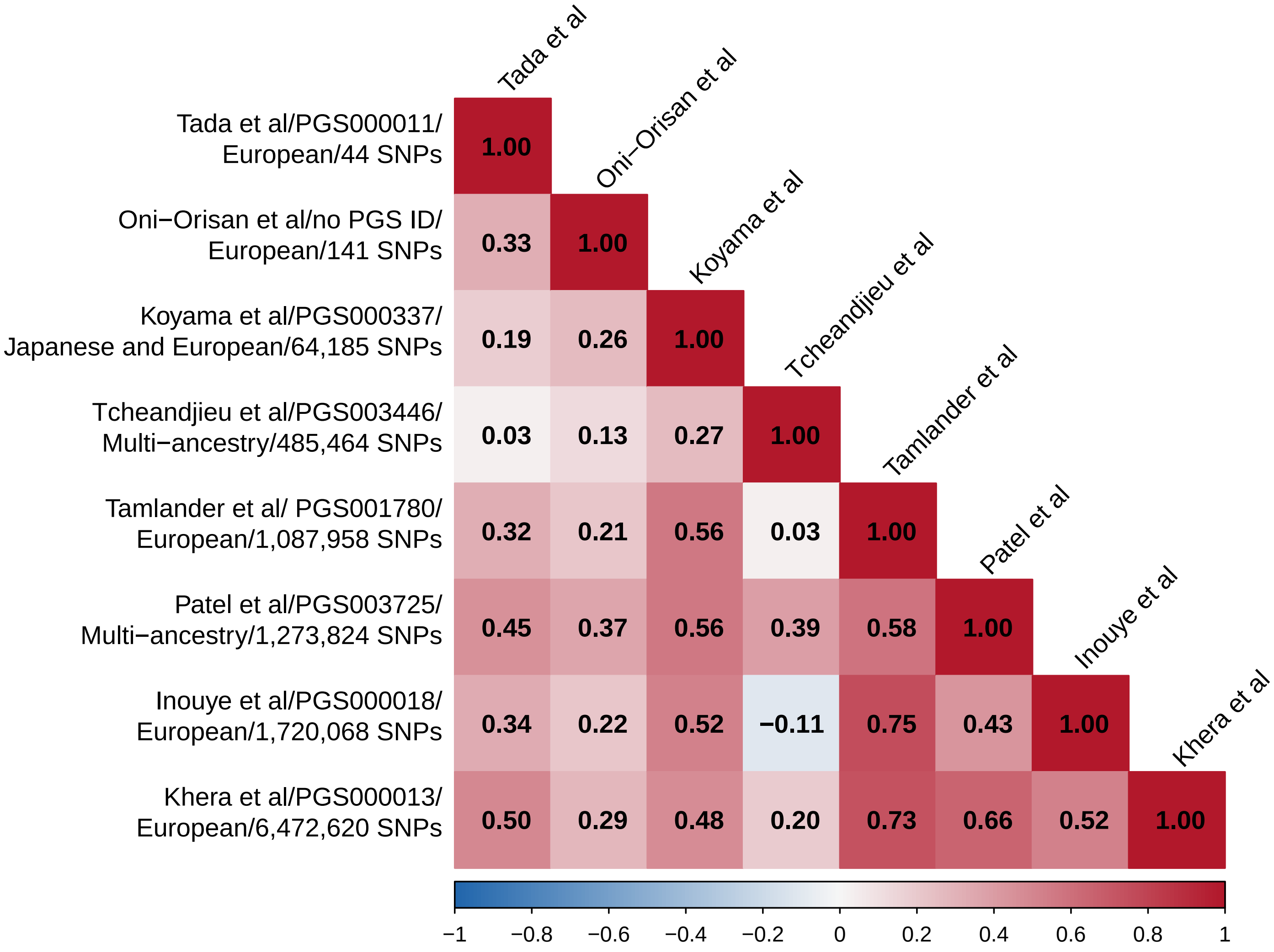


**Webfigure 1: Pairwise correlations between the eight selected PRS**

| PRS / age at risk | CHD cases | CHD controls | OR per SD (95% CI) | AUC (95%CI) |
|---|---|---|---|---|
| **Tada et al/PGS000011/ European/44 SNPs** | | | | |
| 35-54 | 832 | 5,329 | 1.08 (1.00-1.16) | 0.69 (0.67-0.71) |
| 55-64 | 1,238 | 44,611 | 1.01 (0.95-1.07) | 0.83 (0.82-0.85) |
| 65-79 | 3,093 | 51,435 | 1.03 (0.99-1.07) | 0.75 (0.74-0.76) |
| Overall | 5,163 | 128,044 | 1.05 (1.02-1.08) | 0.75 (0.74-0.75) |
| | | | *Trend: X²(df=1)=0.58 (p=0.45)* | |
| | | | *Heterogeneity:Q=1.96 (p=0.38)* | |
| **Oni-Orisan et al/no PGS ID/ European/141 SNPs** | | | | |
| 35-54 | 832 | 5,329 | 1.08 (1.00-1.16) | 0.69 (0.67-0.71) |
| 55-64 | 1,238 | 44,611 | 1.08 (1.02-1.14) | 0.83 (0.82-0.85) |
| 65-79 | 3,093 | 51,435 | 1.08 (1.04-1.12) | 0.75 (0.74-0.76) |
| Overall | 5,163 | 128,044 | 1.09 (1.06-1.13) | 0.75 (0.74-0.75) |
| | | | *Trend: X²(df=1)=0 (p=1)* | |
| | | | *Heterogeneity:Q=0 (p=1)* | |
| **Koyama et al/PGS000337/ Japanese and European/64,185 SNPs** | | | | |
| 35-54 | 832 | 5,329 | 1.18 (1.09-1.27) | 0.70 (0.68-0.71) |
| 55-64 | 1,238 | 44,611 | 1.22 (1.15-1.29) | 0.84 (0.82-0.85) |
| 65-79 | 3,093 | 51,435 | 1.10 (1.06-1.15) | 0.75 (0.74-0.76) |
| Overall | 5,163 | 128,044 | 1.20 (1.17-1.23) | 0.75 (0.74-0.75) |
| | | | *Trend: X²(df=1)=5.76 (p=0.016)* | |
| | | | *Heterogeneity:Q=9.04 (p=0.011)* | |
| **Tcheandjieu et al/PGS003446/ Multi-ancestry/485,464 SNPs** | | | | |
| 35-54 | 832 | 5,329 | 1.09 (0.99-1.19) | 0.69 (0.67-0.71) |
| 55-64 | 1,238 | 44,611 | 1.22 (1.13-1.31) | 0.84 (0.82-0.85) |
| 65-79 | 3,093 | 51,435 | 1.06 (1.01-1.11) | 0.75 (0.74-0.76) |
| Overall | 5,163 | 128,044 | 1.16 (1.12-1.20) | 0.75 (0.74-0.75) |
| | | | *Trend: X²(df=1)=2.62 (p=0.11)* | |
| | | | *Heterogeneity:Q=10.48 (p=0.0053)* | |
| **Tamlander et al/ PGS001780/ European/1,087,958 SNPs** | | | | |
| 35-54 | 832 | 5,329 | 1.13 (1.04-1.23) | 0.69 (0.67-0.71) |
| 55-64 | 1,238 | 44,611 | 1.16 (1.09-1.24) | 0.83 (0.82-0.85) |
| 65-79 | 3,093 | 51,435 | 1.12 (1.08-1.17) | 0.75 (0.74-0.76) |
| Overall | 5,163 | 128,044 | 1.20 (1.16-1.23) | 0.75 (0.74-0.75) |
| | | | *Trend: X²(df=1)=0.24 (p=0.62)* | |
| | | | *Heterogeneity:Q=0.83 (p=0.66)* | |
| **Patel et al/PGS003725/ Multi-ancestry/1,273,824 SNPs** | | | | |
| 35-54 | 832 | 5,329 | 1.11 (1.03-1.20) | 0.69 (0.67-0.71) |
| 55-64 | 1,238 | 44,611 | 1.19 (1.12-1.26) | 0.84 (0.82-0.85) |
| 65-79 | 3,093 | 51,435 | 1.12 (1.08-1.16) | 0.75 (0.74-0.76) |
| Overall | 5,163 | 128,044 | 1.24 (1.20-1.28) | 0.75 (0.74-0.76) |
| | | | *Trend: X²(df=1)=0.18 (p=0.67)* | |
| | | | *Heterogeneity:Q=3.15 (p=0.21)* | |
| **Inouye et al/PGS000018/ European/1,720,068 SNPs** | | | | |
| 35-54 | 832 | 5,329 | 1.20 (1.09-1.32) | 0.69 (0.67-0.71) |
| 55-64 | 1,238 | 44,611 | 1.13 (1.06-1.22) | 0.83 (0.82-0.85) |
| 65-79 | 3,093 | 51,435 | 1.10 (1.05-1.15) | 0.75 (0.74-0.76) |
| Overall | 5,163 | 128,044 | 1.19 (1.15-1.23) | 0.75 (0.74-0.75) |
| | | | *Trend: X²(df=1)=2.7 (p=0.1)* | |
| | | | *Heterogeneity:Q=2.84 (p=0.24)* | |
| **Khera et al/PGS000013/ European/6,472,620 SNPs** | | | | |
| 35-54 | 832 | 5,329 | 1.14 (1.05-1.23) | 0.69 (0.67-0.71) |
| 55-64 | 1,238 | 44,611 | 1.15 (1.09-1.22) | 0.84 (0.82-0.85) |
| 65-79 | 3,093 | 51,435 | 1.12 (1.08-1.16) | 0.75 (0.74-0.76) |
| Overall | 5,163 | 128,044 | 1.17 (1.14-1.20) | 0.75 (0.74-0.75) |
| | | | *Trend: X²(df=1)=0.39 (p=0.53)* | |
| | | | *Heterogeneity:Q=0.61 (p=0.74)* | |

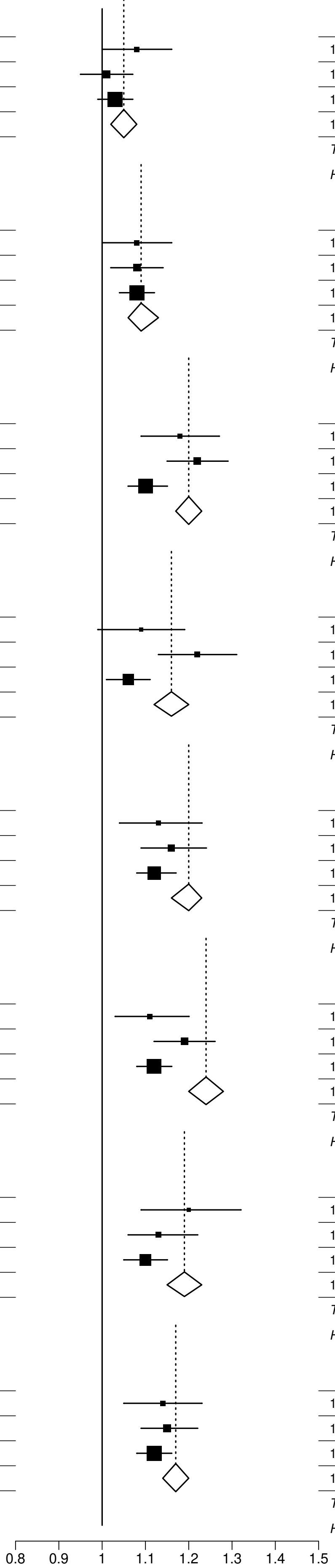


**Webfigure 2: Odds of premature CHD per 1SD increase in each PRS, at different ages at risk**
Analyses as for Figure 2, stratified by age-at-risk 25-54, 55-64 and 65-79.

| PRS / education level | CHD cases | CHD controls | OR per SD (95% CI) | AUC (95%CI) |
|---|---|---|---|---|
| **Tada et al/PGS000011/ European/44 SNPs** | | | | |
| University/college | 564 | 20,145 | 1.11 (1.02-1.21) | 0.79 (0.77-0.81) |
| High school | 958 | 32,474 | 1.02 (0.96-1.09) | 0.78 (0.76-0.79) |
| Elementary | 2,841 | 60,134 | 1.05 (1.01-1.09) | 0.73 (0.72-0.74) |
| Other | 800 | 15,291 | 1.05 (0.98-1.13) | 0.65 (0.63-0.67) |
| Overall | 5,163 | 128,044 | 1.05 (1.02-1.08) | 0.75 (0.74-0.75) |
| | | | *Trend: X²(df=1)=0.29 (p=0.59)* | |
| | | | *Heterogeneity:Q=2.4 (p=0.49)* | |
| **Oni-Orisan et al/no PGS ID/ European/141 SNPs** | | | | |
| University/college | 564 | 20,145 | 1.22 (1.12-1.33) | 0.79 (0.77-0.81) |
| High school | 958 | 32,474 | 1.07 (1.00-1.14) | 0.78 (0.76-0.79) |
| Elementary | 2,841 | 60,134 | 1.06 (1.02-1.10) | 0.73 (0.72-0.74) |
| Other | 800 | 15,291 | 1.15 (1.07-1.24) | 0.65 (0.64-0.67) |
| Overall | 5,163 | 128,044 | 1.09 (1.06-1.13) | 0.75 (0.74-0.75) |
| | | | *Trend: X²(df=1)=1.14 (p=0.29)* | |
| | | | *Heterogeneity:Q=11.12 (p=0.011)* | |
| **Koyama et al/PGS000337/ Japanese and European/64,185 SNPs** | | | | |
| University/college | 564 | 20,145 | 1.29 (1.19-1.41) | 0.80 (0.78-0.82) |
| High school | 958 | 32,474 | 1.22 (1.14-1.30) | 0.78 (0.76-0.79) |
| Elementary | 2,841 | 60,134 | 1.20 (1.15-1.25) | 0.73 (0.72-0.74) |
| Other | 800 | 15,291 | 1.12 (1.04-1.21) | 0.65 (0.63-0.67) |
| Overall | 5,163 | 128,044 | 1.20 (1.17-1.23) | 0.75 (0.74-0.75) |
| | | | *Trend: X²(df=1)=5.76 (p=0.016)* | |
| | | | *Heterogeneity:Q=6.43 (p=0.092)* | |
| **Tcheandjieu et al/PGS003446/ Multi-ancestry/485,464 SNPs** | | | | |
| University/college | 564 | 20,145 | 1.32 (1.19-1.47) | 0.80 (0.78-0.81) |
| High school | 958 | 32,474 | 1.15 (1.06-1.24) | 0.78 (0.76-0.79) |
| Elementary | 2,841 | 60,134 | 1.14 (1.09-1.20) | 0.73 (0.72-0.74) |
| Other | 800 | 15,291 | 1.13 (1.03-1.23) | 0.65 (0.63-0.67) |
| Overall | 5,163 | 128,044 | 1.16 (1.12-1.20) | 0.75 (0.74-0.75) |
| | | | *Trend: X²(df=1)=4.28 (p=0.039)* | |
| | | | *Heterogeneity:Q=6.78 (p=0.079)* | |
| **Tamlander et al/ PGS001780/ European/1,087,958 SNPs** | | | | |
| University/college | 564 | 20,145 | 1.31 (1.19-1.43) | 0.79 (0.78-0.81) |
| High school | 958 | 32,474 | 1.19 (1.11-1.28) | 0.78 (0.76-0.79) |
| Elementary | 2,841 | 60,134 | 1.20 (1.15-1.25) | 0.73 (0.72-0.74) |
| Other | 800 | 15,291 | 1.10 (1.02-1.19) | 0.65 (0.63-0.67) |
| Overall | 5,163 | 128,044 | 1.20 (1.16-1.23) | 0.75 (0.74-0.75) |
| | | | *Trend: X²(df=1)=6.21 (p=0.013)* | |
| | | | *Heterogeneity:Q=8.38 (p=0.039)* | |
| **Patel et al/PGS003725/ Multi-ancestry/1,273,824 SNPs** | | | | |
| University/college | 564 | 20,145 | 1.40 (1.29-1.53) | 0.80 (0.78-0.82) |
| High school | 958 | 32,474 | 1.26 (1.18-1.35) | 0.78 (0.76-0.79) |
| Elementary | 2,841 | 60,134 | 1.22 (1.17-1.27) | 0.73 (0.72-0.74) |
| Other | 800 | 15,291 | 1.17 (1.09-1.26) | 0.65 (0.64-0.67) |
| Overall | 5,163 | 128,044 | 1.24 (1.20-1.28) | 0.75 (0.74-0.76) |
| | | | *Trend: X²(df=1)=9.96 (p=0.0016)* | |
| | | | *Heterogeneity:Q=10.91 (p=0.012)* | |
| **Inouye et al/PGS000018/ European/1,720,068 SNPs** | | | | |
| University/college | 564 | 20,145 | 1.31 (1.19-1.45) | 0.79 (0.78-0.81) |
| High school | 958 | 32,474 | 1.19 (1.10-1.29) | 0.78 (0.76-0.79) |
| Elementary | 2,841 | 60,134 | 1.18 (1.13-1.24) | 0.73 (0.72-0.74) |
| Other | 800 | 15,291 | 1.15 (1.05-1.26) | 0.65 (0.63-0.67) |
| Overall | 5,163 | 128,044 | 1.19 (1.15-1.23) | 0.75 (0.74-0.75) |
| | | | *Trend: X²(df=1)=3.36 (p=0.067)* | |
| | | | *Heterogeneity:Q=4.3 (p=0.23)* | |
| **Khera et al/PGS000013/ European/6,472,620 SNPs** | | | | |
| University/college | 564 | 20,145 | 1.25 (1.15-1.36) | 0.79 (0.77-0.81) |
| High school | 958 | 32,474 | 1.17 (1.10-1.25) | 0.78 (0.76-0.79) |
| Elementary | 2,841 | 60,134 | 1.17 (1.13-1.22) | 0.73 (0.72-0.74) |
| Other | 800 | 15,291 | 1.11 (1.03-1.19) | 0.65 (0.63-0.67) |
| Overall | 5,163 | 128,044 | 1.17 (1.14-1.20) | 0.75 (0.74-0.75) |
| | | | *Trend: X²(df=1)=3.62 (p=0.057)* | |
| | | | *Heterogeneity:Q=4.47 (p=0.21)* | |

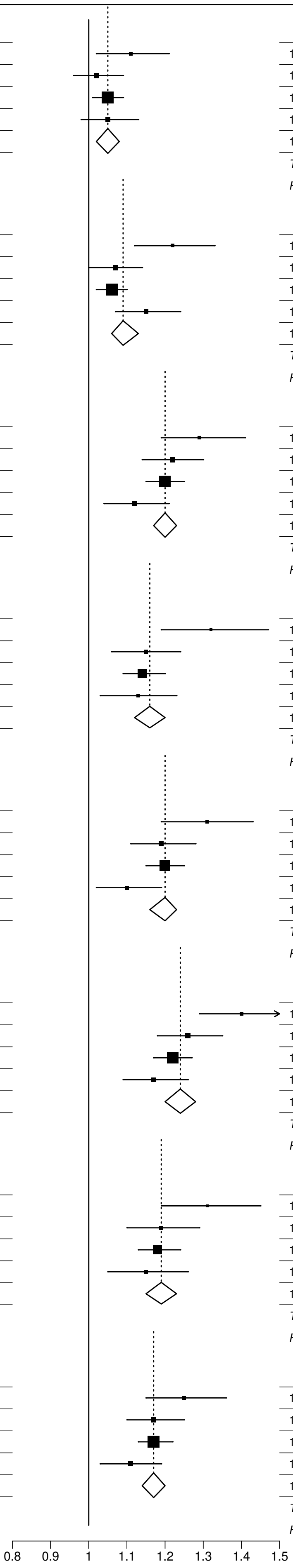


**Webfigure 3: Odds of premature CHD per 1SD increase in each PRS, by highest level of education**
Analyses as for Figure 2, stratified by levels of education.

| PRS / waist-to-hip Ratio | CHD cases | CHD controls | OR per SD (95% CI) | AUC (95%CI) |
|---|---|---|---|---|
| **Tada et al/PGS000011/ European/44 SNPs** | | | | |
| <0.85 | 682 | 30,630 | 1.01 (0.93-1.09) | 0.77 (0.76-0.79) |
| 0.85 to <0.9 | 1,171 | 34,786 | 1.06 (1.00-1.12) | 0.75 (0.74-0.77) |
| ≥0.9 | 3,310 | 62,628 | 1.05 (1.02-1.09) | 0.71 (0.70-0.72) |
| Overall | 5,163 | 128,044 | 1.05 (1.02-1.08) | 0.75 (0.74-0.75) |
| | | | *Trend: X²(df=1)=0.41 (p=0.52)* | |
| | | | *Heterogeneity:Q=1.02 (p=0.6)* | |
| **Oni-Orisan et al/no PGS ID/ European/141 SNPs** | | | | |
| <0.85 | 682 | 30,630 | 1.04 (0.96-1.13) | 0.78 (0.76-0.79) |
| 0.85 to <0.9 | 1,171 | 34,786 | 1.06 (1.00-1.12) | 0.75 (0.74-0.77) |
| ≥0.9 | 3,310 | 62,628 | 1.12 (1.08-1.16) | 0.71 (0.70-0.72) |
| Overall | 5,163 | 128,044 | 1.09 (1.06-1.13) | 0.75 (0.74-0.75) |
| | | | *Trend: X²(df=1)=4.16 (p=0.041)* | |
| | | | *Heterogeneity:Q=4.39 (p=0.11)* | |
| **Koyama et al/PGS000337/ Japanese and European/64,185 SNPs** | | | | |
| <0.85 | 682 | 30,630 | 1.17 (1.08-1.26) | 0.78 (0.76-0.79) |
| 0.85 to <0.9 | 1,171 | 34,786 | 1.09 (1.03-1.16) | 0.75 (0.74-0.77) |
| ≥0.9 | 3,310 | 62,628 | 1.24 (1.20-1.29) | 0.72 (0.71-0.72) |
| Overall | 5,163 | 128,044 | 1.20 (1.17-1.23) | 0.75 (0.74-0.75) |
| | | | *Trend: X²(df=1)=6.2 (p=0.013)* | |
| | | | *Heterogeneity:Q=13.13 (p=0.0014)* | |
| **Tcheandjieu et al/PGS003446/ Multi-ancestry/485,464 SNPs** | | | | |
| <0.85 | 682 | 30,630 | 1.02 (0.93-1.12) | 0.77 (0.76-0.79) |
| 0.85 to <0.9 | 1,171 | 34,786 | 1.08 (1.00-1.16) | 0.75 (0.74-0.77) |
| ≥0.9 | 3,310 | 62,628 | 1.22 (1.17-1.27) | 0.71 (0.70-0.72) |
| Overall | 5,163 | 128,044 | 1.16 (1.12-1.20) | 0.75 (0.74-0.75) |
| | | | *Trend: X²(df=1)=15.78 (p<0.001)* | |
| | | | *Heterogeneity:Q=16.29 (p<0.001)* | |
| **Tamlander et al/ PGS001780/ European/1,087,958 SNPs** | | | | |
| <0.85 | 682 | 30,630 | 1.11 (1.03-1.21) | 0.78 (0.76-0.79) |
| 0.85 to <0.9 | 1,171 | 34,786 | 1.15 (1.07-1.22) | 0.75 (0.74-0.77) |
| ≥0.9 | 3,310 | 62,628 | 1.23 (1.19-1.28) | 0.71 (0.71-0.72) |
| Overall | 5,163 | 128,044 | 1.20 (1.16-1.23) | 0.75 (0.74-0.75) |
| | | | *Trend: X²(df=1)=6.49 (p=0.011)* | |
| | | | *Heterogeneity:Q=6.65 (p=0.036)* | |
| **Patel et al/PGS003725/ Multi-ancestry/1,273,824 SNPs** | | | | |
| <0.85 | 682 | 30,630 | 1.11 (1.03-1.21) | 0.78 (0.76-0.79) |
| 0.85 to <0.9 | 1,171 | 34,786 | 1.19 (1.12-1.26) | 0.75 (0.74-0.77) |
| ≥0.9 | 3,310 | 62,628 | 1.28 (1.24-1.33) | 0.72 (0.71-0.73) |
| Overall | 5,163 | 128,044 | 1.24 (1.20-1.28) | 0.75 (0.74-0.76) |
| | | | *Trend: X²(df=1)=12.19 (p<0.001)* | |
| | | | *Heterogeneity:Q=12.19 (p=0.0023)* | |
| **Inouye et al/PGS000018/ European/1,720,068 SNPs** | | | | |
| <0.85 | 682 | 30,630 | 1.11 (1.01-1.21) | 0.78 (0.76-0.79) |
| 0.85 to <0.9 | 1,171 | 34,786 | 1.11 (1.04-1.20) | 0.75 (0.74-0.77) |
| ≥0.9 | 3,310 | 62,628 | 1.24 (1.19-1.30) | 0.71 (0.71-0.72) |
| Overall | 5,163 | 128,044 | 1.19 (1.15-1.23) | 0.75 (0.74-0.75) |
| | | | *Trend: X²(df=1)=7.82 (p=0.0052)* | |
| | | | *Heterogeneity:Q=9.33 (p=0.0094)* | |
| **Khera et al/PGS000013/ European/6,472,620 SNPs** | | | | |
| <0.85 | 682 | 30,630 | 1.09 (1.01-1.18) | 0.78 (0.76-0.79) |
| 0.85 to <0.9 | 1,171 | 34,786 | 1.11 (1.05-1.18) | 0.75 (0.74-0.77) |
| ≥0.9 | 3,310 | 62,628 | 1.21 (1.17-1.25) | 0.71 (0.71-0.72) |
| Overall | 5,163 | 128,044 | 1.17 (1.14-1.20) | 0.75 (0.74-0.75) |
| | | | *Trend: X²(df=1)=8.92 (p=0.0028)* | |
| | | | *Heterogeneity:Q=9.76 (p=0.0076)* | |

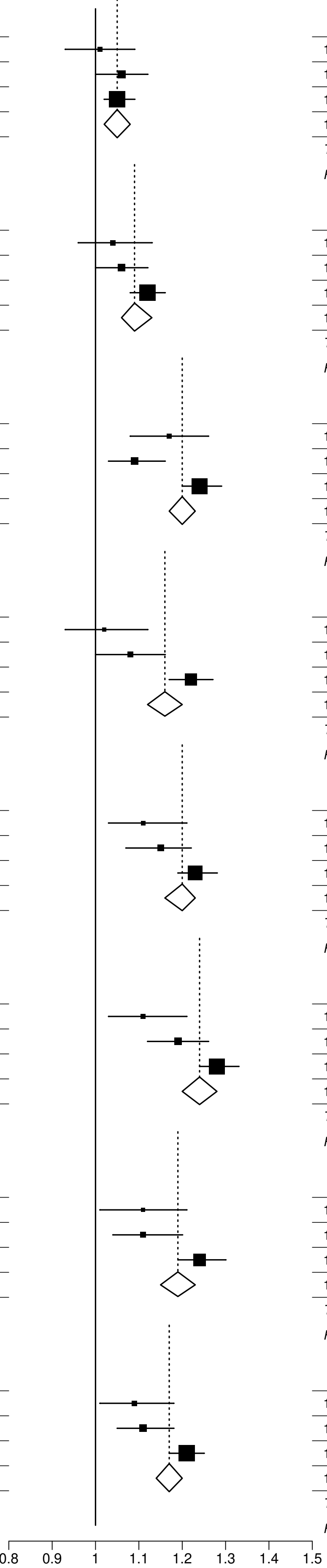


**Webfigure 4: Odds of premature CHD per 1SD increase in each PRS, by waist-to-hip ratio**
Analyses as for Figure 2, stratified by waist-to-hip ration <0.85 0.85 to <0.9 and ≥0.9.

| PRS / systolic blood pressure, mmHg | CHD cases | CHD controls | OR per SD (95% CI) | AUC (95%CI) |
|---|---|---|---|---|
| **Tada et al/PGS000011/ European/44 SNPs** | | | | |
| <120 | 990 | 39,975 | 1.06 (1.00-1.13) | 0.77 (0.76-0.79) |
| 120-139 | 2,411 | 63,641 | 1.08 (1.04-1.13) | 0.73 (0.72-0.74) |
| ≥140 | 1,762 | 24,428 | 1.00 (0.95-1.05) | 0.66 (0.64-0.67) |
| Overall | 5,163 | 128,044 | 1.05 (1.02-1.08) | 0.75 (0.74-0.75) |
| | | | *Trend: X²(df=1)=3.09 (p=0.079)* | |
| | | | *Heterogeneity:Q=5.75 (p=0.056)* | |
| **Oni-Orisan et al/no PGS ID/ European/141 SNPs** | | | | |
| <120 | 990 | 39,975 | 1.10 (1.03-1.17) | 0.77 (0.76-0.79) |
| 120-139 | 2,411 | 63,641 | 1.09 (1.05-1.14) | 0.73 (0.73-0.74) |
| ≥140 | 1,762 | 24,428 | 1.10 (1.04-1.15) | 0.66 (0.64-0.67) |
| Overall | 5,163 | 128,044 | 1.09 (1.06-1.13) | 0.75 (0.74-0.75) |
| | | | *Trend: X²(df=1)=0 (p=0.95)* | |
| | | | *Heterogeneity:Q=0.1 (p=0.95)* | |
| **Koyama et al/PGS000337/ Japanese and European/64,185 SNPs** | | | | |
| <120 | 990 | 39,975 | 1.29 (1.21-1.38) | 0.78 (0.76-0.79) |
| 120-139 | 2,411 | 63,641 | 1.20 (1.15-1.25) | 0.74 (0.73-0.75) |
| ≥140 | 1,762 | 24,428 | 1.15 (1.10-1.21) | 0.66 (0.65-0.67) |
| Overall | 5,163 | 128,044 | 1.20 (1.17-1.23) | 0.75 (0.74-0.75) |
| | | | *Trend: X²(df=1)=7.32 (p=0.0068)* | |
| | | | *Heterogeneity:Q=7.56 (p=0.023)* | |
| **Tcheandjieu et al/PGS003446/ Multi-ancestry/485,464 SNPs** | | | | |
| <120 | 990 | 39,975 | 1.19 (1.10-1.28) | 0.77 (0.76-0.79) |
| 120-139 | 2,411 | 63,641 | 1.18 (1.12-1.24) | 0.74 (0.73-0.75) |
| ≥140 | 1,762 | 24,428 | 1.11 (1.05-1.18) | 0.66 (0.64-0.67) |
| Overall | 5,163 | 128,044 | 1.16 (1.12-1.20) | 0.75 (0.74-0.75) |
| | | | *Trend: X²(df=1)=2.41 (p=0.12)* | |
| | | | *Heterogeneity:Q=2.95 (p=0.23)* | |
| **Tamlander et al/ PGS001780/ European/1,087,958 SNPs** | | | | |
| <120 | 990 | 39,975 | 1.21 (1.12-1.29) | 0.77 (0.76-0.79) |
| 120-139 | 2,411 | 63,641 | 1.23 (1.17-1.28) | 0.74 (0.73-0.75) |
| ≥140 | 1,762 | 24,428 | 1.15 (1.09-1.21) | 0.66 (0.65-0.67) |
| Overall | 5,163 | 128,044 | 1.20 (1.16-1.23) | 0.75 (0.74-0.75) |
| | | | *Trend: X²(df=1)=2 (p=0.16)* | |
| | | | *Heterogeneity:Q=3.73 (p=0.15)* | |
| **Patel et al/PGS003725/ Multi-ancestry/1,273,824 SNPs** | | | | |
| <120 | 990 | 39,975 | 1.29 (1.21-1.38) | 0.78 (0.76-0.79) |
| 120-139 | 2,411 | 63,641 | 1.26 (1.21-1.32) | 0.74 (0.73-0.75) |
| ≥140 | 1,762 | 24,428 | 1.18 (1.12-1.24) | 0.66 (0.65-0.67) |
| Overall | 5,163 | 128,044 | 1.24 (1.20-1.28) | 0.75 (0.74-0.76) |
| | | | *Trend: X²(df=1)=5.2 (p=0.023)* | |
| | | | *Heterogeneity:Q=5.68 (p=0.058)* | |
| **Inouye et al/PGS000018/ European/1,720,068 SNPs** | | | | |
| <120 | 990 | 39,975 | 1.23 (1.14-1.33) | 0.77 (0.76-0.79) |
| 120-139 | 2,411 | 63,641 | 1.23 (1.17-1.29) | 0.74 (0.73-0.75) |
| ≥140 | 1,762 | 24,428 | 1.12 (1.06-1.19) | 0.66 (0.64-0.67) |
| Overall | 5,163 | 128,044 | 1.19 (1.15-1.23) | 0.75 (0.74-0.75) |
| | | | *Trend: X²(df=1)=4.75 (p=0.029)* | |
| | | | *Heterogeneity:Q=6.49 (p=0.039)* | |
| **Khera et al/PGS000013/ European/6,472,620 SNPs** | | | | |
| <120 | 990 | 39,975 | 1.18 (1.11-1.26) | 0.77 (0.76-0.79) |
| 120-139 | 2,411 | 63,641 | 1.20 (1.15-1.25) | 0.74 (0.73-0.75) |
| ≥140 | 1,762 | 24,428 | 1.13 (1.08-1.19) | 0.66 (0.65-0.67) |
| Overall | 5,163 | 128,044 | 1.17 (1.14-1.20) | 0.75 (0.74-0.75) |
| | | | *Trend: X²(df=1)=1.76 (p=0.18)* | |
| | | | *Heterogeneity:Q=3.47 (p=0.18)* | |

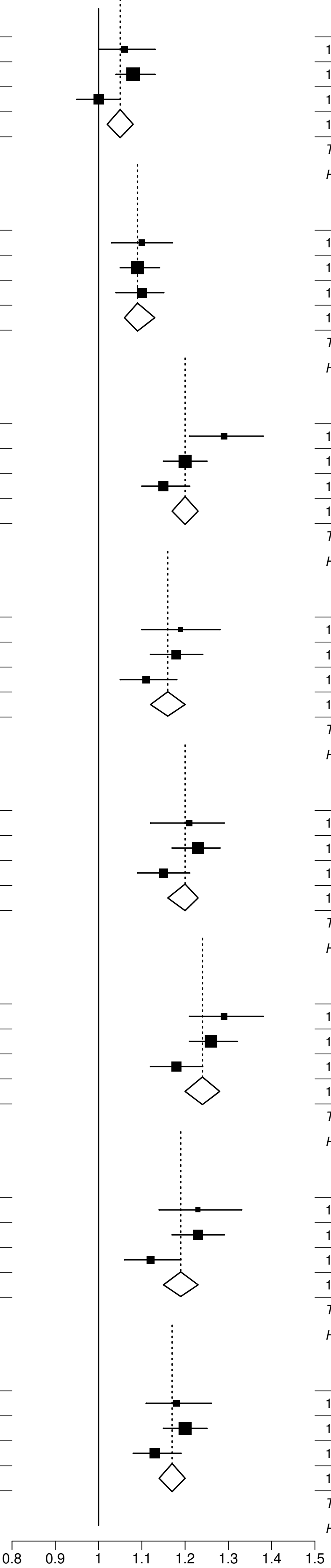


**Webfigure 5: Odds of premature CHD per 1SD increase in each PRS, by systolic blood pressure**
Analyses as for Figure 2, stratified by systolic blood pressure level <120, 120-139 and ≥140.

| PRS / diastolic blood pressure, mmHg | CHD cases | CHD controls | OR per SD (95% CI) | AUC (95%CI) |
|---|---|---|---|---|
| **Tada et al/PGS000011/ European/44 SNPs** | | | | |
| <80 | 1,246 | 40,134 | 1.07 (1.01-1.13) | 0.77 (0.76-0.79) |
| 80-89 | 2,028 | 55,371 | 1.05 (1.01-1.10) | 0.75 (0.74-0.76) |
| ≥90 | 1,889 | 32,539 | 1.03 (0.99-1.08) | 0.69 (0.68-0.70) |
| Overall | 5,163 | 128,044 | 1.05 (1.02-1.08) | 0.75 (0.74-0.75) |
| | | | *Trend: X²(df=1)=1.03 (p=0.31)* | |
| | | | *Heterogeneity:Q=1.03 (p=0.6)* | |
| **Oni-Orisan et al/no PGS ID/ European/141 SNPs** | | | | |
| <80 | 1,246 | 40,134 | 1.10 (1.04-1.17) | 0.77 (0.76-0.79) |
| 80-89 | 2,028 | 55,371 | 1.09 (1.04-1.14) | 0.75 (0.74-0.76) |
| ≥90 | 1,889 | 32,539 | 1.09 (1.04-1.14) | 0.69 (0.68-0.70) |
| Overall | 5,163 | 128,044 | 1.09 (1.06-1.13) | 0.75 (0.74-0.75) |
| | | | *Trend: X²(df=1)=0.05 (p=0.82)* | |
| | | | *Heterogeneity:Q=0.07 (p=0.96)* | |
| **Koyama et al/PGS000337/ Japanese and European/64,185 SNPs** | | | | |
| <80 | 1,246 | 40,134 | 1.28 (1.21-1.36) | 0.78 (0.77-0.79) |
| 80-89 | 2,028 | 55,371 | 1.19 (1.13-1.24) | 0.75 (0.74-0.76) |
| ≥90 | 1,889 | 32,539 | 1.16 (1.10-1.21) | 0.69 (0.68-0.70) |
| Overall | 5,163 | 128,044 | 1.20 (1.17-1.23) | 0.75 (0.74-0.75) |
| | | | *Trend: X²(df=1)=6.15 (p=0.013)* | |
| | | | *Heterogeneity:Q=6.76 (p=0.034)* | |
| **Tcheandjieu et al/PGS003446/ Multi-ancestry/485,464 SNPs** | | | | |
| <80 | 1,246 | 40,134 | 1.19 (1.11-1.27) | 0.77 (0.76-0.79) |
| 80-89 | 2,028 | 55,371 | 1.15 (1.09-1.21) | 0.75 (0.74-0.76) |
| ≥90 | 1,889 | 32,539 | 1.14 (1.08-1.21) | 0.69 (0.68-0.70) |
| Overall | 5,163 | 128,044 | 1.16 (1.12-1.20) | 0.75 (0.74-0.75) |
| | | | *Trend: X²(df=1)=0.8 (p=0.37)* | |
| | | | *Heterogeneity:Q=0.92 (p=0.63)* | |
| **Tamlander et al/ PGS001780/ European/1,087,958 SNPs** | | | | |
| <80 | 1,246 | 40,134 | 1.24 (1.17-1.32) | 0.78 (0.76-0.79) |
| 80-89 | 2,028 | 55,371 | 1.19 (1.13-1.25) | 0.75 (0.74-0.76) |
| ≥90 | 1,889 | 32,539 | 1.17 (1.11-1.23) | 0.69 (0.68-0.70) |
| Overall | 5,163 | 128,044 | 1.20 (1.16-1.23) | 0.75 (0.74-0.75) |
| | | | *Trend: X²(df=1)=1.91 (p=0.17)* | |
| | | | *Heterogeneity:Q=2.05 (p=0.36)* | |
| **Patel et al/PGS003725/ Multi-ancestry/1,273,824 SNPs** | | | | |
| <80 | 1,246 | 40,134 | 1.30 (1.22-1.37) | 0.78 (0.77-0.79) |
| 80-89 | 2,028 | 55,371 | 1.24 (1.18-1.29) | 0.75 (0.74-0.76) |
| ≥90 | 1,889 | 32,539 | 1.20 (1.14-1.26) | 0.69 (0.68-0.70) |
| Overall | 5,163 | 128,044 | 1.24 (1.20-1.28) | 0.75 (0.74-0.76) |
| | | | *Trend: X²(df=1)=4.19 (p=0.041)* | |
| | | | *Heterogeneity:Q=4.24 (p=0.12)* | |
| **Inouye et al/PGS000018/ European/1,720,068 SNPs** | | | | |
| <80 | 1,246 | 40,134 | 1.24 (1.16-1.33) | 0.77 (0.76-0.79) |
| 80-89 | 2,028 | 55,371 | 1.22 (1.15-1.28) | 0.75 (0.74-0.76) |
| ≥90 | 1,889 | 32,539 | 1.13 (1.07-1.20) | 0.69 (0.68-0.70) |
| Overall | 5,163 | 128,044 | 1.19 (1.15-1.23) | 0.75 (0.74-0.75) |
| | | | *Trend: X²(df=1)=4.66 (p=0.031)* | |
| | | | *Heterogeneity:Q=5.37 (p=0.068)* | |
| **Khera et al/PGS000013/ European/6,472,620 SNPs** | | | | |
| <80 | 1,246 | 40,134 | 1.18 (1.12-1.25) | 0.78 (0.76-0.79) |
| 80-89 | 2,028 | 55,371 | 1.17 (1.12-1.22) | 0.75 (0.74-0.76) |
| ≥90 | 1,889 | 32,539 | 1.16 (1.11-1.22) | 0.69 (0.68-0.70) |
| Overall | 5,163 | 128,044 | 1.17 (1.14-1.20) | 0.75 (0.74-0.75) |
| | | | *Trend: X²(df=1)=0.21 (p=0.65)* | |
| | | | *Heterogeneity:Q=0.21 (p=0.9)* | |

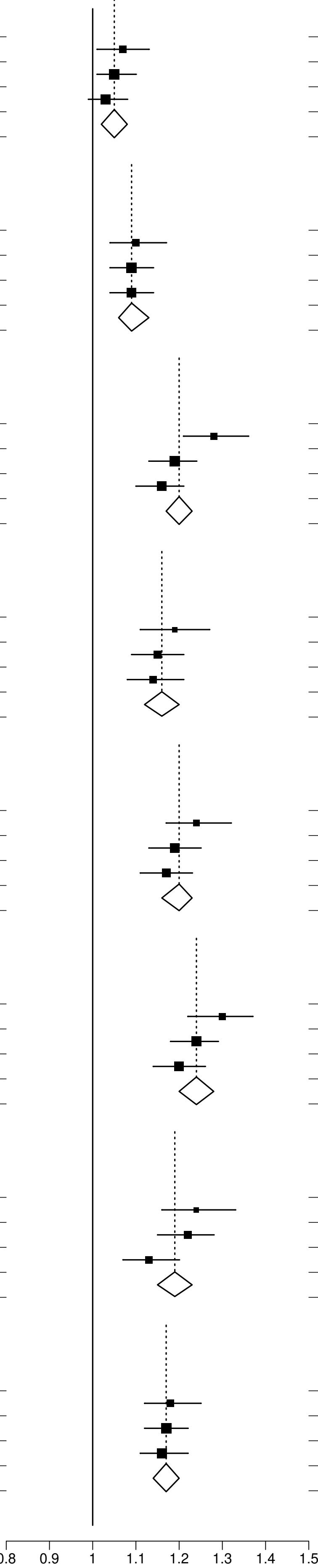


**Webfigure 6: Odds of premature CHD per 1SD increase in each PRS, by diastolic blood pressure**
Analyses as for Figure 2, stratified by diastolic blood pressure level <80, 80-89 and ≥90.

| PRS / smoking status group | CHD cases | CHD controls | OR per SD (95% CI) | AUC (95%CI) |
|---|---|---|---|---|
| **Tada et al/PGS000011/ European/44 SNPs** | | | | |
| Never | 2,166 | 62,535 | 1.07 (1.02-1.12) | 0.74 (0.73-0.75) |
| Former smoker | 1,467 | 24,709 | 1.04 (0.98-1.09) | 0.72 (0.71-0.74) |
| Current smoker | 1,530 | 40,800 | 1.03 (0.98-1.08) | 0.76 (0.74-0.77) |
| Overall | 5,163 | 128,044 | 1.05 (1.02-1.08) | 0.75 (0.74-0.75) |
| | | | *Trend: X²(df=1)=1.3 (p=0.25)* | |
| | | | *Heterogeneity:Q=1.38 (p=0.5)* | |
| **Oni-Orisan et al/no PGS ID/ European/141 SNPs** | | | | |
| Never | 2,166 | 62,535 | 1.06 (1.02-1.11) | 0.74 (0.73-0.75) |
| Former smoker | 1,467 | 24,709 | 1.15 (1.09-1.21) | 0.73 (0.71-0.74) |
| Current smoker | 1,530 | 40,800 | 1.09 (1.03-1.15) | 0.76 (0.75-0.77) |
| Overall | 5,163 | 128,044 | 1.09 (1.06-1.13) | 0.75 (0.74-0.75) |
| | | | *Trend: X²(df=1)=0.98 (p=0.32)* | |
| | | | *Heterogeneity:Q=5.29 (p=0.071)* | |
| **Koyama et al/PGS000337/ Japanese and European/64,185 SNPs** | | | | |
| Never | 2,166 | 62,535 | 1.14 (1.09-1.19) | 0.74 (0.73-0.75) |
| Former smoker | 1,467 | 24,709 | 1.27 (1.21-1.35) | 0.73 (0.72-0.74) |
| Current smoker | 1,530 | 40,800 | 1.22 (1.16-1.29) | 0.76 (0.75-0.77) |
| Overall | 5,163 | 128,044 | 1.20 (1.17-1.23) | 0.75 (0.74-0.75) |
| | | | *Trend: X²(df=1)=4.61 (p=0.032)* | |
| | | | *Heterogeneity:Q=9.68 (p=0.0079)* | |
| **Tcheandjieu et al/PGS003446/ Multi-ancestry/485,464 SNPs** | | | | |
| Never | 2,166 | 62,535 | 1.05 (0.99-1.10) | 0.74 (0.73-0.75) |
| Former smoker | 1,467 | 24,709 | 1.29 (1.21-1.38) | 0.73 (0.72-0.74) |
| Current smoker | 1,530 | 40,800 | 1.22 (1.14-1.30) | 0.76 (0.75-0.77) |
| Overall | 5,163 | 128,044 | 1.16 (1.12-1.20) | 0.75 (0.74-0.75) |
| | | | *Trend: X²(df=1)=15.19 (p<0.001)* | |
| | | | *Heterogeneity:Q=26.1 (p<0.001)* | |
| **Tamlander et al/ PGS001780/ European/1,087,958 SNPs** | | | | |
| Never | 2,166 | 62,535 | 1.13 (1.08-1.19) | 0.74 (0.73-0.75) |
| Former smoker | 1,467 | 24,709 | 1.25 (1.18-1.32) | 0.73 (0.72-0.74) |
| Current smoker | 1,530 | 40,800 | 1.25 (1.18-1.32) | 0.76 (0.75-0.77) |
| Overall | 5,163 | 128,044 | 1.20 (1.16-1.23) | 0.75 (0.74-0.75) |
| | | | *Trend: X²(df=1)=8.14 (p=0.0043)* | |
| | | | *Heterogeneity:Q=10.22 (p=0.006)* | |
| **Patel et al/PGS003725/ Multi-ancestry/1,273,824 SNPs** | | | | |
| Never | 2,166 | 62,535 | 1.17 (1.12-1.23) | 0.74 (0.73-0.75) |
| Former smoker | 1,467 | 24,709 | 1.31 (1.24-1.38) | 0.73 (0.72-0.74) |
| Current smoker | 1,530 | 40,800 | 1.28 (1.21-1.35) | 0.76 (0.75-0.77) |
| Overall | 5,163 | 128,044 | 1.24 (1.20-1.28) | 0.75 (0.74-0.76) |
| | | | *Trend: X²(df=1)=7.48 (p=0.0062)* | |
| | | | *Heterogeneity:Q=11.66 (p=0.0029)* | |
| **Inouye et al/PGS000018/ European/1,720,068 SNPs** | | | | |
| Never | 2,166 | 62,535 | 1.13 (1.07-1.19) | 0.74 (0.73-0.75) |
| Former smoker | 1,467 | 24,709 | 1.28 (1.20-1.36) | 0.73 (0.72-0.74) |
| Current smoker | 1,530 | 40,800 | 1.21 (1.14-1.29) | 0.76 (0.75-0.77) |
| Overall | 5,163 | 128,044 | 1.19 (1.15-1.23) | 0.75 (0.74-0.75) |
| | | | *Trend: X²(df=1)=3.44 (p=0.064)* | |
| | | | *Heterogeneity:Q=8.82 (p=0.012)* | |
| **Khera et al/PGS000013/ European/6,472,620 SNPs** | | | | |
| Never | 2,166 | 62,535 | 1.13 (1.08-1.18) | 0.74 (0.73-0.75) |
| Former smoker | 1,467 | 24,709 | 1.19 (1.13-1.26) | 0.73 (0.71-0.74) |
| Current smoker | 1,530 | 40,800 | 1.21 (1.15-1.27) | 0.76 (0.75-0.77) |
| Overall | 5,163 | 128,044 | 1.17 (1.14-1.20) | 0.75 (0.74-0.75) |
| | | | *Trend: X²(df=1)=4.14 (p=0.042)* | |
| | | | *Heterogeneity:Q=4.44 (p=0.11)* | |

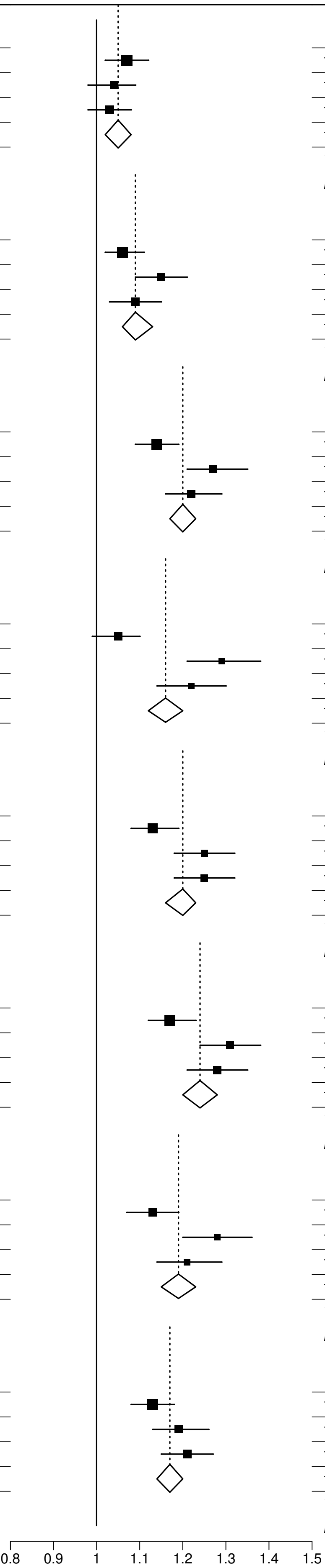


**Webfigure 7: Odds of premature CHD per 1SD increase in each PRS, by smoking status**
Analyses as for Figure 2, stratified by smoking status.

| PRS / diabetes at baseline | CHD cases | CHD controls | OR per SD (95% CI) | AUC (95%CI) |
|---|---|---|---|---|
| **Tada et al/PGS000011/ European/44 SNPs** | | | | |
| No Diabetes | 3,064 | 105,347 | 1.06 (1.02-1.10) | 0.73 (0.72-0.74) |
| Diabetes | 2,099 | 22,697 | 1.03 (0.98-1.07) | 0.64 (0.63-0.65) |
| Overall | 5,163 | 128,044 | 1.05 (1.02-1.08) | 0.75 (0.74-0.75) |
| | | | *Heterogeneity:Q=0.96 (p=0.33)* | |
| **Oni-Orisan et al/no PGS ID/ European/141 SNPs** | | | | |
| No Diabetes | 3,064 | 105,347 | 1.08 (1.04-1.12) | 0.73 (0.72-0.74) |
| Diabetes | 2,099 | 22,697 | 1.11 (1.06-1.16) | 0.64 (0.63-0.65) |
| Overall | 5,163 | 128,044 | 1.09 (1.06-1.13) | 0.75 (0.74-0.75) |
| | | | *Heterogeneity:Q=0.84 (p=0.36)* | |
| **Koyama et al/PGS000337/ Japanese and European/64,185 SNPs** | | | | |
| No Diabetes | 3,064 | 105,347 | 1.18 (1.14-1.23) | 0.74 (0.73-0.74) |
| Diabetes | 2,099 | 22,697 | 1.22 (1.16-1.28) | 0.65 (0.63-0.66) |
| Overall | 5,163 | 128,044 | 1.20 (1.17-1.23) | 0.75 (0.74-0.75) |
| | | | *Heterogeneity:Q=1.23 (p=0.27)* | |
| **Tcheandjieu et al/PGS003446/ Multi-ancestry/485,464 SNPs** | | | | |
| No Diabetes | 3,064 | 105,347 | 1.14 (1.09-1.19) | 0.73 (0.73-0.74) |
| Diabetes | 2,099 | 22,697 | 1.18 (1.12-1.25) | 0.64 (0.63-0.65) |
| Overall | 5,163 | 128,044 | 1.16 (1.12-1.20) | 0.75 (0.74-0.75) |
| | | | *Heterogeneity:Q=0.92 (p=0.34)* | |
| **Tamlander et al/ PGS001780/ European/1,087,958 SNPs** | | | | |
| No Diabetes | 3,064 | 105,347 | 1.18 (1.14-1.23) | 0.73 (0.73-0.74) |
| Diabetes | 2,099 | 22,697 | 1.21 (1.15-1.27) | 0.65 (0.63-0.66) |
| Overall | 5,163 | 128,044 | 1.20 (1.16-1.23) | 0.75 (0.74-0.75) |
| | | | *Heterogeneity:Q=0.62 (p=0.43)* | |
| **Patel et al/PGS003725/ Multi-ancestry/1,273,824 SNPs** | | | | |
| No Diabetes | 3,064 | 105,347 | 1.22 (1.17-1.26) | 0.74 (0.73-0.74) |
| Diabetes | 2,099 | 22,697 | 1.26 (1.21-1.32) | 0.65 (0.64-0.66) |
| Overall | 5,163 | 128,044 | 1.24 (1.20-1.28) | 0.75 (0.74-0.76) |
| | | | *Heterogeneity:Q=1.13 (p=0.29)* | |
| **Inouye et al/PGS000018/ European/1,720,068 SNPs** | | | | |
| No Diabetes | 3,064 | 105,347 | 1.19 (1.14-1.24) | 0.73 (0.73-0.74) |
| Diabetes | 2,099 | 22,697 | 1.19 (1.13-1.26) | 0.64 (0.63-0.65) |
| Overall | 5,163 | 128,044 | 1.19 (1.15-1.23) | 0.75 (0.74-0.75) |
| | | | *Heterogeneity:Q=0 (p=1)* | |
| **Khera et al/PGS000013/ European/6,472,620 SNPs** | | | | |
| No Diabetes | 3,064 | 105,347 | 1.17 (1.13-1.21) | 0.73 (0.73-0.74) |
| Diabetes | 2,099 | 22,697 | 1.17 (1.12-1.22) | 0.64 (0.63-0.66) |
| Overall | 5,163 | 128,044 | 1.17 (1.14-1.20) | 0.75 (0.74-0.75) |
| | | | *Heterogeneity:Q=0 (p=1)* | |

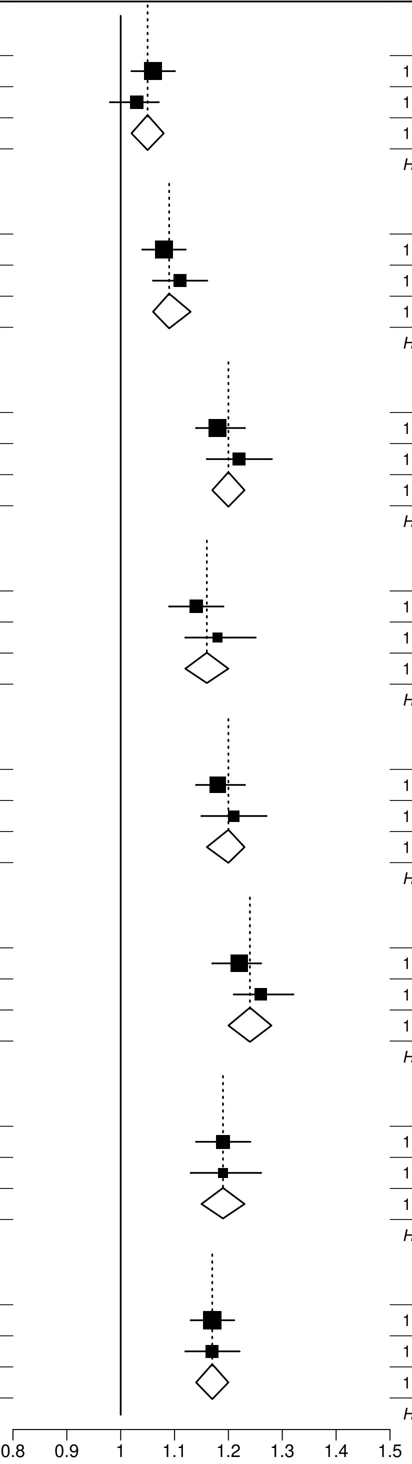


**Webfigure 8: Odds of premature CHD per 1SD increase in each PRS, by baseline diabetes status**
Analyses as for Figure 2, stratified by baseline diabetes status.

| PRS / indigenous ancestry proportion | CHD cases | CHD controls | OR per SD (95% CI) | AUC (95%CI) |
|---|---|---|---|---|
| **Tada et al/PGS000011/ European/44 SNPs** | | | | |
| <60% | 2,131 | 46,104 | 1.03 (0.98-1.07) | 0.75 (0.74-0.76) |
| 60% to <80% | 1,786 | 46,115 | 1.08 (1.03-1.13) | 0.75 (0.74-0.76) |
| ≥80% | 1,159 | 33,742 | 1.03 (0.97-1.10) | 0.72 (0.71-0.74) |
| Overall | 5,163 | 128,044 | 1.05 (1.02-1.08) | 0.75 (0.74-0.75) |
| | | | *Trend: $X^2$(df=1)=0.08 (p=0.78)* | |
| | | | *Heterogeneity:Q=2.45 (p=0.29)* | |
| **Oni-Orisan et al/no PGS ID/ European/141 SNPs** | | | | |
| <60% | 2,131 | 46,104 | 1.10 (1.06-1.15) | 0.75 (0.74-0.76) |
| 60% to <80% | 1,786 | 46,115 | 1.10 (1.05-1.15) | 0.75 (0.74-0.76) |
| ≥80% | 1,159 | 33,742 | 1.07 (1.00-1.14) | 0.72 (0.71-0.74) |
| Overall | 5,163 | 128,044 | 1.09 (1.06-1.13) | 0.75 (0.74-0.75) |
| | | | *Trend: $X^2$(df=1)=0.38 (p=0.54)* | |
| | | | *Heterogeneity:Q=0.58 (p=0.75)* | |
| **Koyama et al/PGS000337/ Japanese and European/64,185 SNPs** | | | | |
| <60% | 2,131 | 46,104 | 1.19 (1.14-1.24) | 0.75 (0.74-0.76) |
| 60% to <80% | 1,786 | 46,115 | 1.23 (1.17-1.30) | 0.76 (0.75-0.77) |
| ≥80% | 1,159 | 33,742 | 1.17 (1.10-1.25) | 0.73 (0.71-0.74) |
| Overall | 5,163 | 128,044 | 1.20 (1.17-1.23) | 0.75 (0.74-0.75) |
| | | | *Trend: $X^2$(df=1)=0.03 (p=0.87)* | |
| | | | *Heterogeneity:Q=1.74 (p=0.42)* | |
| **Tcheandjieu et al/PGS003446/ Multi-ancestry/485,464 SNPs** | | | | |
| <60% | 2,131 | 46,104 | 1.18 (1.12-1.24) | 0.75 (0.74-0.76) |
| 60% to <80% | 1,786 | 46,115 | 1.16 (1.09-1.23) | 0.75 (0.74-0.76) |
| ≥80% | 1,159 | 33,742 | 1.13 (1.05-1.23) | 0.72 (0.71-0.74) |
| Overall | 5,163 | 128,044 | 1.16 (1.12-1.20) | 0.75 (0.74-0.75) |
| | | | *Trend: $X^2$(df=1)=0.81 (p=0.37)* | |
| | | | *Heterogeneity:Q=0.82 (p=0.66)* | |
| **Tamlander et al/ PGS001780/ European/1,087,958 SNPs** | | | | |
| <60% | 2,131 | 46,104 | 1.19 (1.14-1.25) | 0.75 (0.74-0.76) |
| 60% to <80% | 1,786 | 46,115 | 1.24 (1.18-1.30) | 0.76 (0.75-0.77) |
| ≥80% | 1,159 | 33,742 | 1.15 (1.07-1.23) | 0.72 (0.71-0.74) |
| Overall | 5,163 | 128,044 | 1.20 (1.16-1.23) | 0.75 (0.74-0.75) |
| | | | *Trend: $X^2$(df=1)=0.19 (p=0.66)* | |
| | | | *Heterogeneity:Q=3.16 (p=0.21)* | |
| **Patel et al/PGS003725/ Multi-ancestry/1,273,824 SNPs** | | | | |
| <60% | 2,131 | 46,104 | 1.22 (1.17-1.27) | 0.76 (0.75-0.77) |
| 60% to <80% | 1,786 | 46,115 | 1.30 (1.24-1.37) | 0.76 (0.75-0.77) |
| ≥80% | 1,159 | 33,742 | 1.19 (1.12-1.27) | 0.73 (0.71-0.74) |
| Overall | 5,163 | 128,044 | 1.24 (1.20-1.28) | 0.75 (0.74-0.76) |
| | | | *Trend: $X^2$(df=1)=0.01 (p=0.93)* | |
| | | | *Heterogeneity:Q=5.61 (p=0.061)* | |
| **Inouye et al/PGS000018/ European/1,720,068 SNPs** | | | | |
| <60% | 2,131 | 46,104 | 1.16 (1.11-1.22) | 0.75 (0.74-0.76) |
| 60% to <80% | 1,786 | 46,115 | 1.24 (1.17-1.31) | 0.75 (0.74-0.77) |
| ≥80% | 1,159 | 33,742 | 1.18 (1.09-1.28) | 0.73 (0.71-0.74) |
| Overall | 5,163 | 128,044 | 1.19 (1.15-1.23) | 0.75 (0.74-0.75) |
| | | | *Trend: $X^2$(df=1)=0.6 (p=0.44)* | |
| | | | *Heterogeneity:Q=2.9 (p=0.23)* | |
| **Khera et al/PGS000013/ European/6,472,620 SNPs** | | | | |
| <60% | 2,131 | 46,104 | 1.15 (1.10-1.20) | 0.75 (0.74-0.76) |
| 60% to <80% | 1,786 | 46,115 | 1.24 (1.18-1.30) | 0.76 (0.75-0.77) |
| ≥80% | 1,159 | 33,742 | 1.12 (1.05-1.20) | 0.72 (0.71-0.74) |
| Overall | 5,163 | 128,044 | 1.17 (1.14-1.20) | 0.75 (0.74-0.75) |
| | | | *Trend: $X^2$(df=1)=0 (p=0.97)* | |
| | | | *Heterogeneity:Q=7.86 (p=0.02)* | |

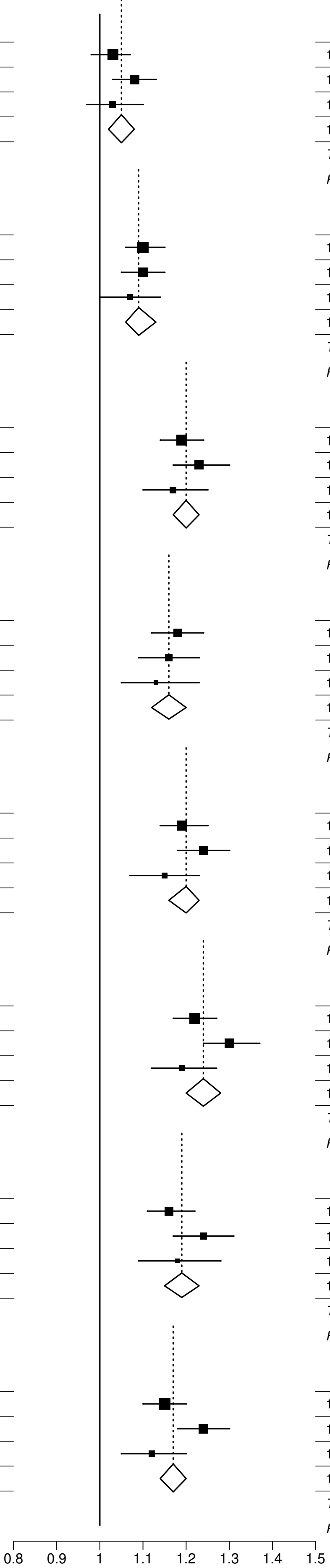


**Webfigure 9: Odds of premature CHD per 1SD increase in each PRS, by level of Indigenous American ancestry**
Analyses as for Figure 2, stratified by indigenous ancestry proportion <60%, 60% to <80% and ≥80%.

| PRS name | Cases | Controls | OR per SD (95% CI) | AUC (95%CI) |
|---|---|---|---|---|
| **Tada et al/PGS000011/ European/44 SNPs** | | | | |
| Partially adjusted | 494 | 132,713 | 1.07 (0.98−1.17) | 0.74 (0.72−0.76) |
| Fully adjusted | 494 | 132,713 | 1.07 (0.98−1.17) | 0.75 (0.73−0.77) |
| **Oni−Orisan et al/no PGS ID/ European/141 SNPs** | | | | |
| Partially adjusted | 494 | 132,713 | 1.05 (0.96−1.15) | 0.74 (0.72−0.76) |
| Fully adjusted | 494 | 132,713 | 1.05 (0.96−1.15) | 0.75 (0.73−0.77) |
| **Koyama et al/PGS000337/ Japanese and European/64,185 SNPs** | | | | |
| Partially adjusted | 494 | 132,713 | 1.21 (1.11−1.33) | 0.74 (0.72−0.76) |
| Fully adjusted | 494 | 132,713 | 1.21 (1.10−1.32) | 0.75 (0.73−0.77) |
| **Tcheandjieu et al/PGS003446/ Multi−ancestry/485,464 SNPs** | | | | |
| Partially adjusted | 494 | 132,713 | 1.11 (1.00−1.24) | 0.74 (0.72−0.76) |
| Fully adjusted | 494 | 132,713 | 1.11 (1.00−1.23) | 0.75 (0.73−0.77) |
| **Tamlander et al/ PGS001780/ European/1,087,958 SNPs** | | | | |
| Partially adjusted | 494 | 132,713 | 1.11 (1.01−1.22) | 0.74 (0.72−0.76) |
| Fully adjusted | 494 | 132,713 | 1.10 (1.00−1.21) | 0.75 (0.73−0.77) |
| **Patel et al/PGS003725/ Multi−ancestry/1,273,824 SNPs** | | | | |
| Partially adjusted | 494 | 132,713 | 1.15 (1.05−1.25) | 0.74 (0.72−0.76) |
| Fully adjusted | 494 | 132,713 | 1.14 (1.04−1.24) | 0.75 (0.73−0.77) |
| **Inouye et al/PGS000018/ European/1,720,068 SNPs** | | | | |
| Partially adjusted | 494 | 132,713 | 1.15 (1.04−1.28) | 0.74 (0.72−0.76) |
| Fully adjusted | 494 | 132,713 | 1.14 (1.03−1.27) | 0.75 (0.73−0.77) |
| **Khera et al/PGS000013/ European/6,472,620 SNPs** | | | | |
| Partially adjusted | 494 | 132,713 | 1.16 (1.06−1.27) | 0.74 (0.72−0.76) |
| Fully adjusted | 494 | 132,713 | 1.16 (1.06−1.26) | 0.75 (0.73−0.77) |

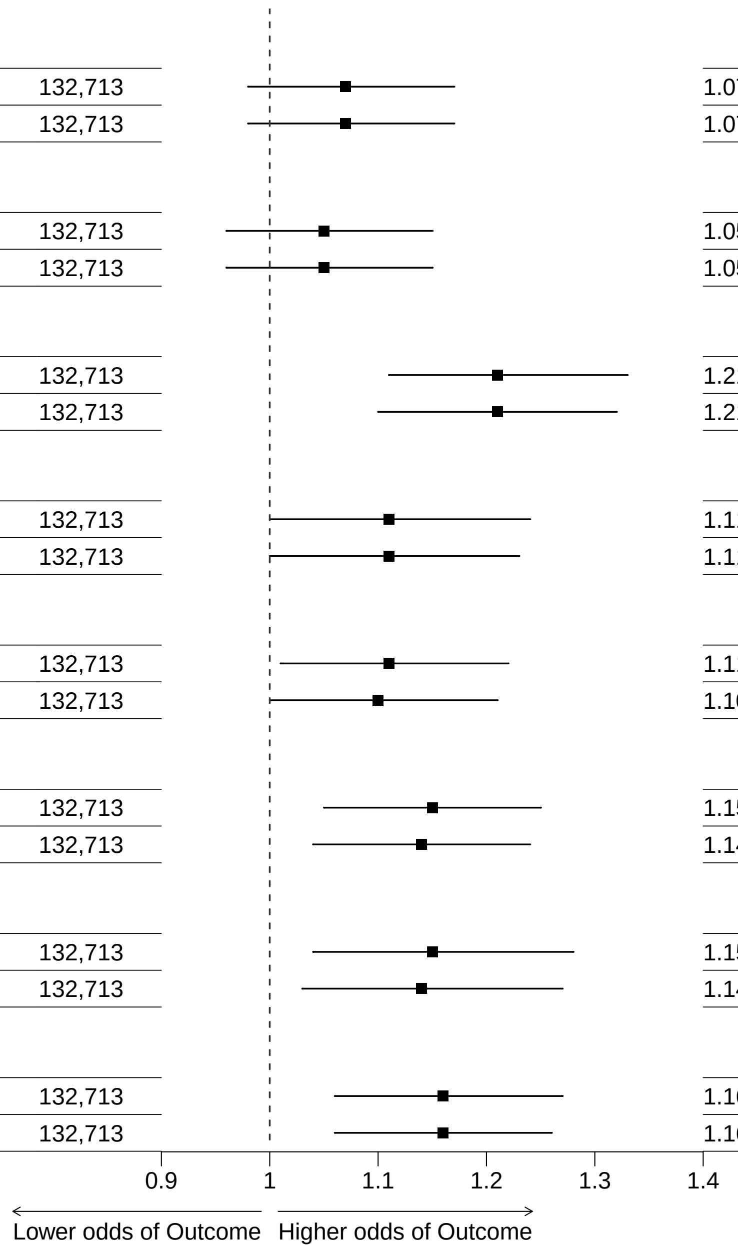


**Webfigure 10: Odds of baseline self-reported angina per 1SD increase in each PRS, for participants aged 35-79 years at recruitment**
Analyses as for Figure 2

| PRS name | Cases | Controls | OR per SD (95% CI) | AUC (95%CI) |
|---|---|---|---|---|
| **Tada et al/PGS000011/ European/44 SNPs** | | | | |
| Partially adjusted | 1,476 | 131,731 | 1.05 (1.00−1.11) | 0.76 (0.75−0.77) |
| Fully adjusted | 1,476 | 131,731 | 1.05 (0.99−1.10) | 0.77 (0.76−0.78) |
| **Oni−Orisan et al/no PGS ID/ European/141 SNPs** | | | | |
| Partially adjusted | 1,476 | 131,731 | 1.13 (1.08−1.19) | 0.76 (0.75−0.77) |
| Fully adjusted | 1,476 | 131,731 | 1.13 (1.07−1.19) | 0.77 (0.76−0.78) |
| **Koyama et al/PGS000337/ Japanese and European/64,185 SNPs** | | | | |
| Partially adjusted | 1,476 | 131,731 | 1.30 (1.24−1.37) | 0.76 (0.75−0.78) |
| Fully adjusted | 1,476 | 131,731 | 1.29 (1.22−1.36) | 0.78 (0.76−0.79) |
| **Tcheandjieu et al/PGS003446/ Multi−ancestry/485,464 SNPs** | | | | |
| Partially adjusted | 1,476 | 131,731 | 1.30 (1.22−1.38) | 0.76 (0.75−0.77) |
| Fully adjusted | 1,476 | 131,731 | 1.28 (1.20−1.37) | 0.77 (0.76−0.79) |
| **Tamlander et al/ PGS001780/ European/1,087,958 SNPs** | | | | |
| Partially adjusted | 1,476 | 131,731 | 1.30 (1.23−1.37) | 0.76 (0.75−0.77) |
| Fully adjusted | 1,476 | 131,731 | 1.29 (1.22−1.36) | 0.77 (0.76−0.79) |
| **Patel et al/PGS003725/ Multi−ancestry/1,273,824 SNPs** | | | | |
| Partially adjusted | 1,476 | 131,731 | 1.36 (1.30−1.44) | 0.77 (0.75−0.78) |
| Fully adjusted | 1,476 | 131,731 | 1.34 (1.27−1.41) | 0.78 (0.76−0.79) |
| **Inouye et al/PGS000018/ European/1,720,068 SNPs** | | | | |
| Partially adjusted | 1,476 | 131,731 | 1.31 (1.23−1.39) | 0.76 (0.75−0.77) |
| Fully adjusted | 1,476 | 131,731 | 1.30 (1.22−1.38) | 0.77 (0.76−0.79) |
| **Khera et al/PGS000013/ European/6,472,620 SNPs** | | | | |
| Partially adjusted | 1,476 | 131,731 | 1.26 (1.19−1.32) | 0.76 (0.75−0.77) |
| Fully adjusted | 1,476 | 131,731 | 1.25 (1.19−1.31) | 0.77 (0.76−0.79) |

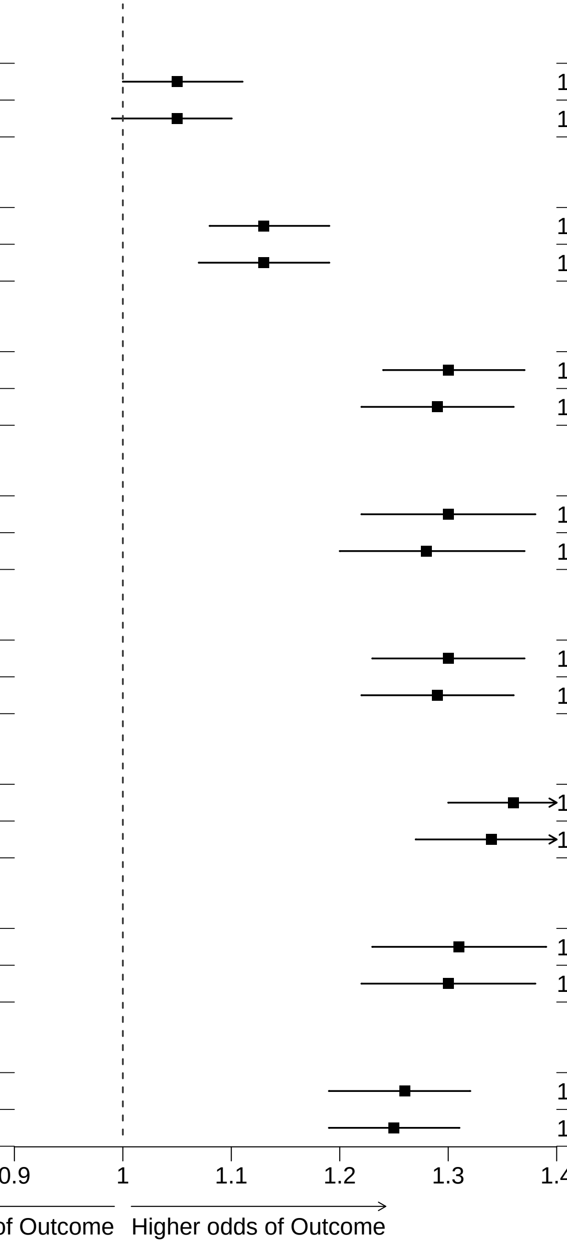


**Webfigure 11: Odds of baseline self-reported myocardial infarction per 1SD increase in each PRS, for participants aged 35-79 years at recruitment**
Analyses as for Figure 2

| PRS name | Cases | Controls | OR per SD (95% CI) | AUC (95%CI) |
|---|---|---|---|---|
| **Tada et al/PGS000011/ European/44 SNPs** | | | | |
| Partially adjusted | 1,901 | 131,306 | 1.05 (1.01−1.10) | 0.75 (0.74−0.76) |
| Fully adjusted | 1,901 | 131,306 | 1.05 (1.00−1.10) | 0.76 (0.75−0.77) |
| **Oni−Orisan et al/no PGS ID/ European/141 SNPs** | | | | |
| Partially adjusted | 1,901 | 131,306 | 1.11 (1.06−1.16) | 0.75 (0.74−0.76) |
| Fully adjusted | 1,901 | 131,306 | 1.11 (1.06−1.16) | 0.76 (0.75−0.77) |
| **Koyama et al/PGS000337/ Japanese and European/64,185 SNPs** | | | | |
| Partially adjusted | 1,901 | 131,306 | 1.28 (1.22−1.34) | 0.75 (0.74−0.76) |
| Fully adjusted | 1,901 | 131,306 | 1.27 (1.21−1.32) | 0.76 (0.75−0.78) |
| **Tcheandjieu et al/PGS003446/ Multi−ancestry/485,464 SNPs** | | | | |
| Partially adjusted | 1,901 | 131,306 | 1.25 (1.18−1.32) | 0.75 (0.74−0.76) |
| Fully adjusted | 1,901 | 131,306 | 1.23 (1.16−1.30) | 0.76 (0.75−0.77) |
| **Tamlander et al/ PGS001780/ European/1,087,958 SNPs** | | | | |
| Partially adjusted | 1,901 | 131,306 | 1.24 (1.18−1.31) | 0.75 (0.74−0.76) |
| Fully adjusted | 1,901 | 131,306 | 1.23 (1.17−1.29) | 0.76 (0.75−0.77) |
| **Patel et al/PGS003725/ Multi−ancestry/1,273,824 SNPs** | | | | |
| Partially adjusted | 1,901 | 131,306 | 1.30 (1.24−1.36) | 0.75 (0.74−0.76) |
| Fully adjusted | 1,901 | 131,306 | 1.27 (1.22−1.34) | 0.76 (0.75−0.77) |
| **Inouye et al/PGS000018/ European/1,720,068 SNPs** | | | | |
| Partially adjusted | 1,901 | 131,306 | 1.26 (1.20−1.33) | 0.75 (0.74−0.76) |
| Fully adjusted | 1,901 | 131,306 | 1.25 (1.18−1.32) | 0.76 (0.75−0.77) |
| **Khera et al/PGS000013/ European/6,472,620 SNPs** | | | | |
| Partially adjusted | 1,901 | 131,306 | 1.23 (1.17−1.28) | 0.75 (0.74−0.76) |
| Fully adjusted | 1,901 | 131,306 | 1.22 (1.17−1.28) | 0.76 (0.75−0.77) |

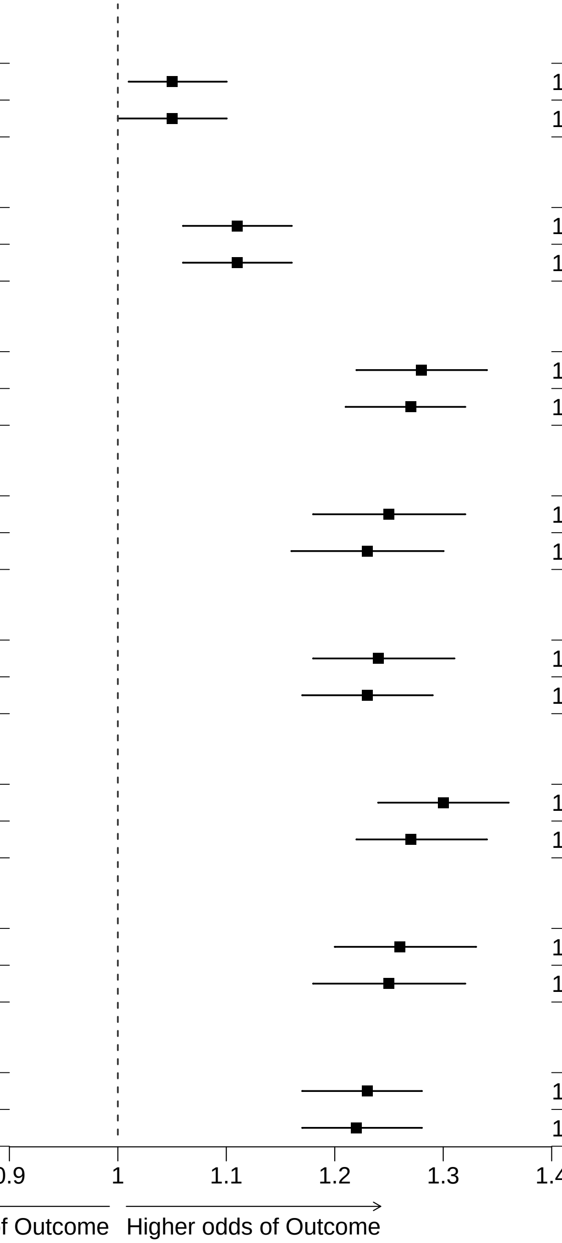


**Webfigure 12: Odds of baseline self-reported angina or myocardial infarction per 1SD increase in each PRS, for participants aged 35-79 years at recruitment**
Analyses as for Figure 2

| PRS name | Cases | Controls | OR per SD (95% CI) | AUC (95%CI) |
|---|---|---|---|---|
| **Tada et al/PGS000011/ European/44 SNPs** | | | | |
| Partially adjusted | 4,649 | 128,558 | 1.05 (1.02−1.08) | 0.72 (0.71−0.72) |
| Fully adjusted | 4,649 | 128,558 | 1.04 (1.01−1.07) | 0.75 (0.74−0.76) |
| **Oni−Orisan et al/no PGS ID/ European/141 SNPs** | | | | |
| Partially adjusted | 4,649 | 128,558 | 1.10 (1.06−1.13) | 0.72 (0.71−0.72) |
| Fully adjusted | 4,649 | 128,558 | 1.09 (1.06−1.12) | 0.75 (0.74−0.76) |
| **Koyama et al/PGS000337/ Japanese and European/64,185 SNPs** | | | | |
| Partially adjusted | 4,649 | 128,558 | 1.25 (1.21−1.28) | 0.72 (0.72−0.73) |
| Fully adjusted | 4,649 | 128,558 | 1.21 (1.18−1.25) | 0.75 (0.75−0.76) |
| **Tcheandjieu et al/PGS003446/ Multi−ancestry/485,464 SNPs** | | | | |
| Partially adjusted | 4,649 | 128,558 | 1.21 (1.17−1.26) | 0.72 (0.71−0.73) |
| Fully adjusted | 4,649 | 128,558 | 1.18 (1.13−1.22) | 0.75 (0.74−0.76) |
| **Tamlander et al/ PGS001780/ European/1,087,958 SNPs** | | | | |
| Partially adjusted | 4,649 | 128,558 | 1.23 (1.19−1.27) | 0.72 (0.71−0.73) |
| Fully adjusted | 4,649 | 128,558 | 1.20 (1.17−1.24) | 0.75 (0.75−0.76) |
| **Patel et al/PGS003725/ Multi−ancestry/1,273,824 SNPs** | | | | |
| Partially adjusted | 4,649 | 128,558 | 1.30 (1.26−1.34) | 0.73 (0.72−0.73) |
| Fully adjusted | 4,649 | 128,558 | 1.25 (1.21−1.28) | 0.75 (0.75−0.76) |
| **Inouye et al/PGS000018/ European/1,720,068 SNPs** | | | | |
| Partially adjusted | 4,649 | 128,558 | 1.24 (1.19−1.28) | 0.72 (0.71−0.73) |
| Fully adjusted | 4,649 | 128,558 | 1.21 (1.17−1.25) | 0.75 (0.75−0.76) |
| **Khera et al/PGS000013/ European/6,472,620 SNPs** | | | | |
| Partially adjusted | 4,649 | 128,558 | 1.20 (1.16−1.23) | 0.72 (0.71−0.73) |
| Fully adjusted | 4,649 | 128,558 | 1.18 (1.15−1.22) | 0.75 (0.75−0.76) |

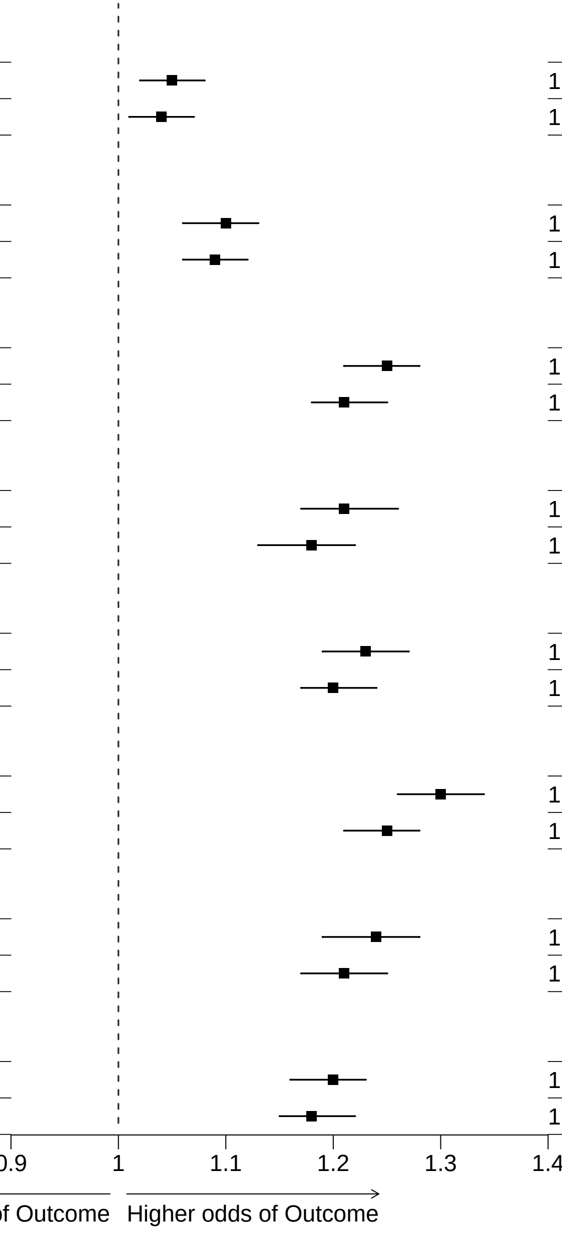


**Webfigure 13: Odds of premature CHD (now defined as baseline self-reported angina or myocardial infarction, or death before age 80 with CHD listed as the primary cause of death) per 1SD increase in each PRS, for participants aged 35-79 years at recruitment**

Analyses as for Figure 2

| PRS name | Cases | Controls | OR per SD (95% CI) | AUC (95%CI) |
|---|---|---|---|---|
| **Tada et al/PGS000011/ European/44 SNPs** | | | | |
| Partially adjusted | 3,479 | 129,728 | 1.06 (1.02−1.09) | 0.70 (0.70−0.71) |
| Fully adjusted | 3,479 | 129,728 | 1.05 (1.01−1.08) | 0.75 (0.74−0.76) |
| **Oni−Orisan et al/no PGS ID/ European/141 SNPs** | | | | |
| Partially adjusted | 3,479 | 129,728 | 1.10 (1.06−1.14) | 0.71 (0.70−0.71) |
| Fully adjusted | 3,479 | 129,728 | 1.09 (1.06−1.13) | 0.75 (0.74−0.76) |
| **Koyama et al/PGS000337/ Japanese and European/64,185 SNPs** | | | | |
| Partially adjusted | 3,479 | 129,728 | 1.21 (1.17−1.25) | 0.71 (0.70−0.72) |
| Fully adjusted | 3,479 | 129,728 | 1.17 (1.13−1.21) | 0.75 (0.74−0.76) |
| **Tcheandjieu et al/PGS003446/ Multi−ancestry/485,464 SNPs** | | | | |
| Partially adjusted | 3,479 | 129,728 | 1.17 (1.13−1.22) | 0.71 (0.70−0.71) |
| Fully adjusted | 3,479 | 129,728 | 1.13 (1.09−1.18) | 0.75 (0.74−0.76) |
| **Tamlander et al/ PGS001780/ European/1,087,958 SNPs** | | | | |
| Partially adjusted | 3,479 | 129,728 | 1.22 (1.17−1.26) | 0.71 (0.70−0.72) |
| Fully adjusted | 3,479 | 129,728 | 1.18 (1.14−1.23) | 0.75 (0.74−0.76) |
| **Patel et al/PGS003725/ Multi−ancestry/1,273,824 SNPs** | | | | |
| Partially adjusted | 3,479 | 129,728 | 1.30 (1.25−1.34) | 0.71 (0.71−0.72) |
| Fully adjusted | 3,479 | 129,728 | 1.23 (1.19−1.28) | 0.75 (0.74−0.76) |
| **Inouye et al/PGS000018/ European/1,720,068 SNPs** | | | | |
| Partially adjusted | 3,479 | 129,728 | 1.21 (1.16−1.26) | 0.71 (0.70−0.72) |
| Fully adjusted | 3,479 | 129,728 | 1.17 (1.13−1.22) | 0.75 (0.74−0.76) |
| **Khera et al/PGS000013/ European/6,472,620 SNPs** | | | | |
| Partially adjusted | 3,479 | 129,728 | 1.17 (1.13−1.21) | 0.71 (0.70−0.72) |
| Fully adjusted | 3,479 | 129,728 | 1.15 (1.11−1.19) | 0.75 (0.74−0.76) |

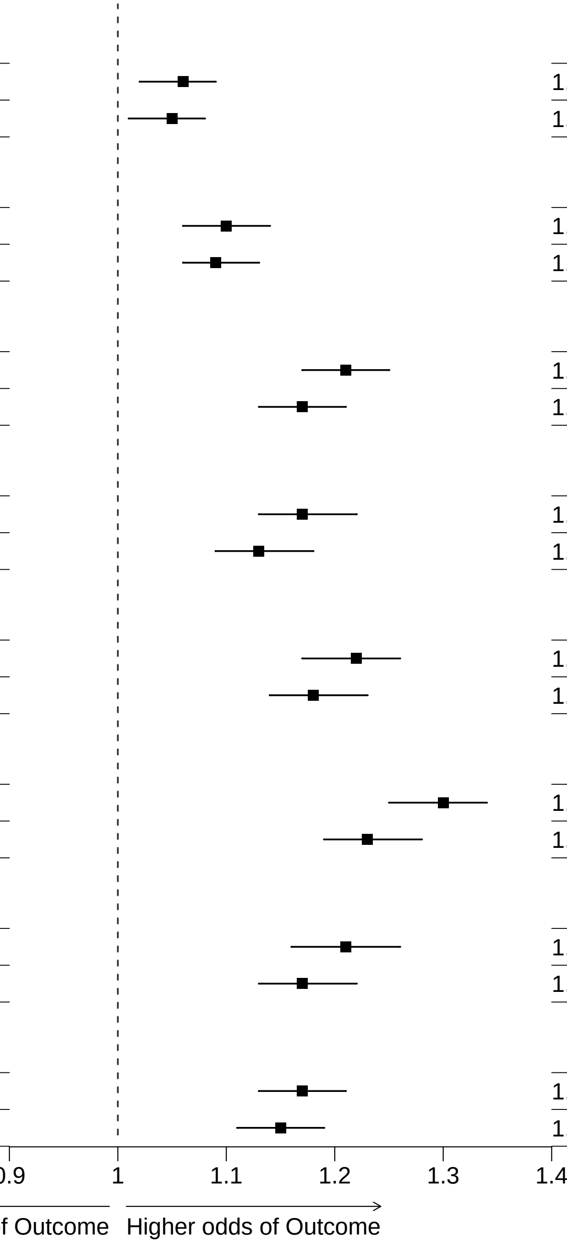


**Webfigure 14: Odds of death before age 80 with CHD listed anywhere on the death certificate, per 1SD increase in each PRS**
Analyses as for Figure 2

| PRS name | Cases | Controls | OR per SD (95% CI) | AUC (95%CI) |
|---|---|---|---|---|
| **Tada et al/PGS000011/ European/44 SNPs** | | | | |
| Partially adjusted | 2,927 | 130,280 | 1.05 (1.01−1.09) | 0.70 (0.70−0.71) |
| Fully adjusted | 2,927 | 130,280 | 1.04 (1.00−1.08) | 0.76 (0.75−0.76) |
| **Oni−Orisan et al/no PGS ID/ European/141 SNPs** | | | | |
| Partially adjusted | 2,927 | 130,280 | 1.09 (1.05−1.13) | 0.70 (0.70−0.71) |
| Fully adjusted | 2,927 | 130,280 | 1.08 (1.04−1.12) | 0.76 (0.75−0.76) |
| **Koyama et al/PGS000337/ Japanese and European/64,185 SNPs** | | | | |
| Partially adjusted | 2,927 | 130,280 | 1.23 (1.19−1.28) | 0.71 (0.70−0.72) |
| Fully adjusted | 2,927 | 130,280 | 1.19 (1.14−1.23) | 0.76 (0.75−0.77) |
| **Tcheandjieu et al/PGS003446/ Multi−ancestry/485,464 SNPs** | | | | |
| Partially adjusted | 2,927 | 130,280 | 1.20 (1.14−1.25) | 0.71 (0.70−0.71) |
| Fully adjusted | 2,927 | 130,280 | 1.15 (1.10−1.20) | 0.76 (0.75−0.76) |
| **Tamlander et al/ PGS001780/ European/1,087,958 SNPs** | | | | |
| Partially adjusted | 2,927 | 130,280 | 1.22 (1.18−1.27) | 0.71 (0.70−0.72) |
| Fully adjusted | 2,927 | 130,280 | 1.19 (1.14−1.24) | 0.76 (0.75−0.77) |
| **Patel et al/PGS003725/ Multi−ancestry/1,273,824 SNPs** | | | | |
| Partially adjusted | 2,927 | 130,280 | 1.31 (1.26−1.36) | 0.71 (0.71−0.72) |
| Fully adjusted | 2,927 | 130,280 | 1.23 (1.19−1.28) | 0.76 (0.75−0.77) |
| **Inouye et al/PGS000018/ European/1,720,068 SNPs** | | | | |
| Partially adjusted | 2,927 | 130,280 | 1.23 (1.18−1.29) | 0.71 (0.70−0.72) |
| Fully adjusted | 2,927 | 130,280 | 1.19 (1.14−1.24) | 0.76 (0.75−0.76) |
| **Khera et al/PGS000013/ European/6,472,620 SNPs** | | | | |
| Partially adjusted | 2,927 | 130,280 | 1.18 (1.14−1.23) | 0.71 (0.70−0.72) |
| Fully adjusted | 2,927 | 130,280 | 1.16 (1.12−1.21) | 0.76 (0.75−0.77) |

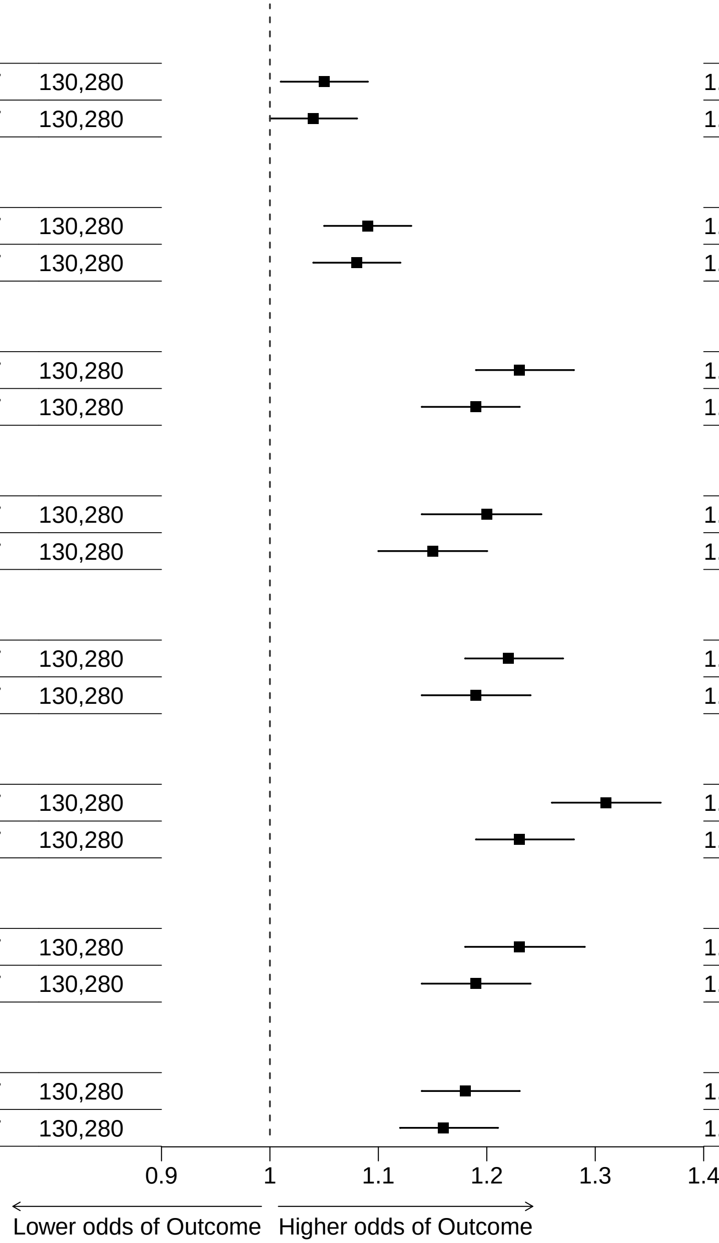


**Webfigure 15: Odds of death before age 80 with CHD listed as the primary cause on the death certificate, per 1SD increase in each PRS**
Analyses as for Figure 2

| Analysis outcome | Cases | Controls | OR per SD (95% CI) | AUC (95%CI) |
|---|---|---|---|---|
| **Tada et al/PGS000011/ European/44 SNPs** | | | | |
| Primary definition of premature CHD | 5,163 | 128,044 | 1.05 (1.03−1.08) | 0.72 (0.71−0.72) |
| Baseline angina | 494 | 132,713 | 1.07 (0.98−1.17) | 0.74 (0.72−0.76) |
| Baseline myocardial infarction | 1,476 | 131,731 | 1.05 (1.00−1.11) | 0.76 (0.75−0.77) |
| Baseline CHD | 1,901 | 131,306 | 1.05 (1.01−1.10) | 0.75 (0.74−0.76) |
| Baseline CHD or CHD death | 4,649 | 128,558 | 1.05 (1.02−1.08) | 0.72 (0.71−0.72) |
| CHD death (any where on the death certificate) | 3,479 | 129,728 | 1.06 (1.02−1.09) | 0.70 (0.70−0.71) |
| CHD death (primary cause on death certificate) | 2,927 | 130,280 | 1.05 (1.01−1.09) | 0.70 (0.70−0.71) |
| Primary definition (among unrelated participants) | 2,947 | 72,928 | 1.06 (1.02−1.10) | 0.72 (0.71−0.72) |
| **Oni−Orisan et al/no PGS ID/ European/141 SNPs** | | | | |
| Primary definition of premature CHD | 5,163 | 128,044 | 1.10 (1.07−1.13) | 0.72 (0.71−0.72) |
| Baseline angina | 494 | 132,713 | 1.05 (0.96−1.15) | 0.74 (0.72−0.76) |
| Baseline myocardial infarction | 1,476 | 131,731 | 1.13 (1.08−1.19) | 0.76 (0.75−0.77) |
| Baseline CHD | 1,901 | 131,306 | 1.11 (1.06−1.16) | 0.75 (0.74−0.76) |
| Baseline CHD or CHD death | 4,649 | 128,558 | 1.10 (1.06−1.13) | 0.72 (0.71−0.72) |
| CHD death (any where on the death certificate) | 3,479 | 129,728 | 1.10 (1.06−1.14) | 0.71 (0.70−0.71) |
| CHD death (primary cause on death certificate) | 2,927 | 130,280 | 1.09 (1.05−1.13) | 0.70 (0.70−0.71) |
| Primary definition (among unrelated participants) | 2,947 | 72,928 | 1.09 (1.05−1.13) | 0.72 (0.71−0.72) |
| **Koyama et al/PGS000337/ Japanese and European/64,185 SNPs** | | | | |
| Primary definition of premature CHD | 5,163 | 128,044 | 1.23 (1.19−1.26) | 0.72 (0.71−0.73) |
| Baseline angina | 494 | 132,713 | 1.21 (1.11−1.33) | 0.74 (0.72−0.76) |
| Baseline myocardial infarction | 1,476 | 131,731 | 1.30 (1.24−1.37) | 0.76 (0.75−0.78) |
| Baseline CHD | 1,901 | 131,306 | 1.28 (1.22−1.34) | 0.75 (0.74−0.76) |
| Baseline CHD or CHD death | 4,649 | 128,558 | 1.25 (1.21−1.28) | 0.72 (0.72−0.73) |
| CHD death (any where on the death certificate) | 3,479 | 129,728 | 1.21 (1.17−1.25) | 0.71 (0.70−0.72) |
| CHD death (primary cause on death certificate) | 2,927 | 130,280 | 1.23 (1.19−1.28) | 0.71 (0.70−0.72) |
| Primary definition (among unrelated participants) | 2,947 | 72,928 | 1.23 (1.19−1.28) | 0.72 (0.71−0.73) |
| **Tcheandjieu et al/PGS003446/ Multi−ancestry/485,464 SNPs** | | | | |
| Primary definition of premature CHD | 5,163 | 128,044 | 1.19 (1.15−1.23) | 0.72 (0.71−0.73) |
| Baseline angina | 494 | 132,713 | 1.11 (1.00−1.24) | 0.74 (0.72−0.76) |
| Baseline myocardial infarction | 1,476 | 131,731 | 1.30 (1.22−1.38) | 0.76 (0.75−0.77) |
| Baseline CHD | 1,901 | 131,306 | 1.25 (1.18−1.32) | 0.75 (0.74−0.76) |
| Baseline CHD or CHD death | 4,649 | 128,558 | 1.21 (1.17−1.26) | 0.72 (0.71−0.73) |
| CHD death (any where on the death certificate) | 3,479 | 129,728 | 1.17 (1.13−1.22) | 0.71 (0.70−0.71) |
| CHD death (primary cause on death certificate) | 2,927 | 130,280 | 1.20 (1.14−1.25) | 0.71 (0.70−0.71) |
| Primary definition (among unrelated participants) | 2,947 | 72,928 | 1.20 (1.14−1.25) | 0.72 (0.71−0.73) |
| **Tamlander et al/ PGS001780/ European/1,087,958 SNPs** | | | | |
| Primary definition of premature CHD | 5,163 | 128,044 | 1.22 (1.19−1.26) | 0.72 (0.71−0.73) |
| Baseline angina | 494 | 132,713 | 1.11 (1.01−1.22) | 0.74 (0.72−0.76) |
| Baseline myocardial infarction | 1,476 | 131,731 | 1.30 (1.23−1.37) | 0.76 (0.75−0.77) |
| Baseline CHD | 1,901 | 131,306 | 1.24 (1.18−1.31) | 0.75 (0.74−0.76) |
| Baseline CHD or CHD death | 4,649 | 128,558 | 1.23 (1.19−1.27) | 0.72 (0.71−0.73) |
| CHD death (any where on the death certificate) | 3,479 | 129,728 | 1.22 (1.17−1.26) | 0.71 (0.70−0.72) |
| CHD death (primary cause on death certificate) | 2,927 | 130,280 | 1.22 (1.18−1.27) | 0.71 (0.70−0.72) |
| Primary definition (among unrelated participants) | 2,947 | 72,928 | 1.22 (1.17−1.27) | 0.72 (0.71−0.73) |
| **Patel et al/PGS003725/ Multi−ancestry/1,273,824 SNPs** | | | | |
| Primary definition of premature CHD | 5,163 | 128,044 | 1.29 (1.25−1.33) | 0.72 (0.72−0.73) |
| Baseline angina | 494 | 132,713 | 1.15 (1.05−1.25) | 0.74 (0.72−0.76) |
| Baseline myocardial infarction | 1,476 | 131,731 | 1.36 (1.30−1.44) | 0.77 (0.75−0.78) |
| Baseline CHD | 1,901 | 131,306 | 1.30 (1.24−1.36) | 0.75 (0.74−0.76) |
| Baseline CHD or CHD death | 4,649 | 128,558 | 1.30 (1.26−1.34) | 0.73 (0.72−0.73) |
| CHD death (any where on the death certificate) | 3,479 | 129,728 | 1.30 (1.25−1.34) | 0.71 (0.71−0.72) |
| CHD death (primary cause on death certificate) | 2,927 | 130,280 | 1.31 (1.26−1.36) | 0.71 (0.71−0.72) |
| Primary definition (among unrelated participants) | 2,947 | 72,928 | 1.29 (1.25−1.34) | 0.72 (0.71−0.73) |
| **Inouye et al/PGS000018/ European/1,720,068 SNPs** | | | | |
| Primary definition of premature CHD | 5,163 | 128,044 | 1.22 (1.18−1.26) | 0.72 (0.71−0.73) |
| Baseline angina | 494 | 132,713 | 1.15 (1.04−1.28) | 0.74 (0.72−0.76) |
| Baseline myocardial infarction | 1,476 | 131,731 | 1.31 (1.23−1.39) | 0.76 (0.75−0.77) |
| Baseline CHD | 1,901 | 131,306 | 1.26 (1.20−1.33) | 0.75 (0.74−0.76) |
| Baseline CHD or CHD death | 4,649 | 128,558 | 1.24 (1.19−1.28) | 0.72 (0.71−0.73) |
| CHD death (any where on the death certificate) | 3,479 | 129,728 | 1.21 (1.16−1.26) | 0.71 (0.70−0.72) |
| CHD death (primary cause on death certificate) | 2,927 | 130,280 | 1.23 (1.18−1.29) | 0.71 (0.70−0.72) |
| Primary definition (among unrelated participants) | 2,947 | 72,928 | 1.23 (1.17−1.28) | 0.72 (0.71−0.73) |
| **Khera et al/PGS000013/ European/6,472,620 SNPs** | | | | |
| Primary definition of premature CHD | 5,163 | 128,044 | 1.19 (1.15−1.22) | 0.72 (0.71−0.73) |
| Baseline angina | 494 | 132,713 | 1.16 (1.06−1.27) | 0.74 (0.72−0.76) |
| Baseline myocardial infarction | 1,476 | 131,731 | 1.26 (1.19−1.32) | 0.76 (0.75−0.77) |
| Baseline CHD | 1,901 | 131,306 | 1.23 (1.17−1.28) | 0.75 (0.74−0.76) |
| Baseline CHD or CHD death | 4,649 | 128,558 | 1.20 (1.16−1.23) | 0.72 (0.71−0.73) |
| CHD death (any where on the death certificate) | 3,479 | 129,728 | 1.17 (1.13−1.21) | 0.71 (0.70−0.72) |
| CHD death (primary cause on death certificate) | 2,927 | 130,280 | 1.18 (1.14−1.23) | 0.71 (0.70−0.72) |
| Primary definition (among unrelated participants) | 2,947 | 72,928 | 1.18 (1.14−1.22) | 0.72 (0.71−0.73) |

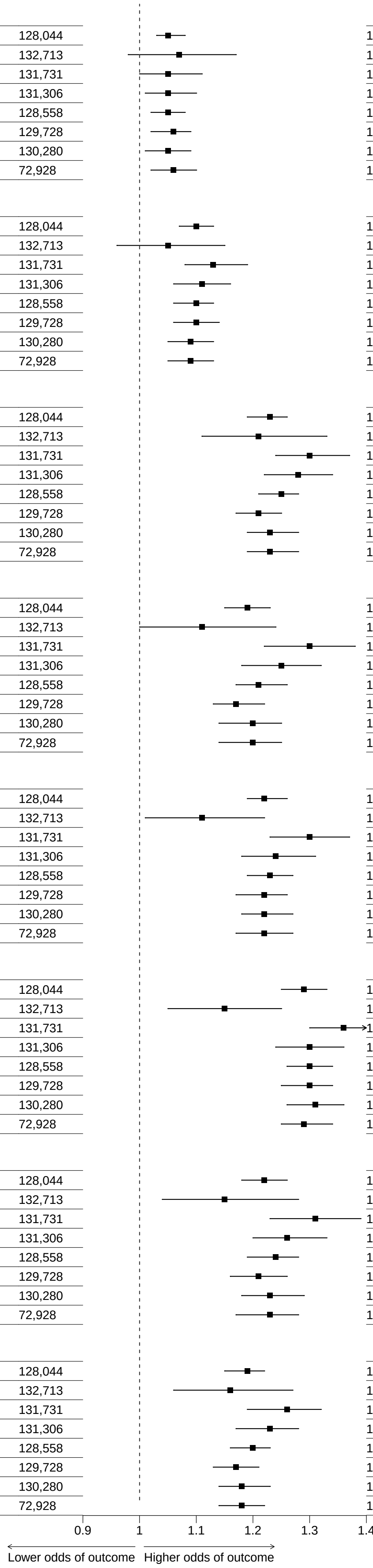


**Webfigure 16: Sensitivity analyses (varying the CHD outcome) with partial adjustments**
Odd ratios (OR) were estimated with regression models adjusted for sex, age at baseline and 7 genetic principal components

| Analysis outcome | Cases | Controls | OR per SD (95% CI) | AUC (95%CI) |
|---|---|---|---|---|
| **Tada et al/PGS000011/ European/44 SNPs** | | | | |
| Primary definition of premature CHD | 5,163 | 128,044 | 1.05 (1.02−1.08) | 0.75 (0.74−0.75) |
| Baseline angina | 494 | 132,713 | 1.07 (0.98−1.17) | 0.75 (0.73−0.77) |
| Baseline myocardial infarction | 1,476 | 131,731 | 1.05 (0.99−1.10) | 0.77 (0.76−0.78) |
| Baseline CHD | 1,901 | 131,306 | 1.05 (1.00−1.10) | 0.76 (0.75−0.77) |
| Baseline CHD or CHD death | 4,649 | 128,558 | 1.04 (1.01−1.07) | 0.75 (0.74−0.76) |
| CHD death (any where on the death certificate) | 3,479 | 129,728 | 1.05 (1.01−1.08) | 0.75 (0.74−0.76) |
| CHD death (primary cause on death certificate) | 2,927 | 130,280 | 1.04 (1.00−1.08) | 0.76 (0.75−0.76) |
| Primary definition (among unrelated participants) | 2,947 | 72,928 | 1.05 (1.01−1.09) | 0.75 (0.74−0.76) |
| **Oni−Orisan et al/no PGS ID/ European/141 SNPs** | | | | |
| Primary definition of premature CHD | 5,163 | 128,044 | 1.09 (1.06−1.13) | 0.75 (0.74−0.75) |
| Baseline angina | 494 | 132,713 | 1.05 (0.96−1.15) | 0.75 (0.73−0.77) |
| Baseline myocardial infarction | 1,476 | 131,731 | 1.13 (1.07−1.19) | 0.77 (0.76−0.78) |
| Baseline CHD | 1,901 | 131,306 | 1.11 (1.06−1.16) | 0.76 (0.75−0.77) |
| Baseline CHD or CHD death | 4,649 | 128,558 | 1.09 (1.06−1.12) | 0.75 (0.74−0.76) |
| CHD death (any where on the death certificate) | 3,479 | 129,728 | 1.09 (1.06−1.13) | 0.75 (0.74−0.76) |
| CHD death (primary cause on death certificate) | 2,927 | 130,280 | 1.08 (1.04−1.12) | 0.76 (0.75−0.76) |
| Primary definition (among unrelated participants) | 2,947 | 72,928 | 1.08 (1.04−1.12) | 0.75 (0.74−0.76) |
| **Koyama et al/PGS000337/ Japanese and European/64,185 SNPs** | | | | |
| Primary definition of premature CHD | 5,163 | 128,044 | 1.20 (1.17−1.23) | 0.75 (0.74−0.75) |
| Baseline angina | 494 | 132,713 | 1.21 (1.10−1.32) | 0.75 (0.73−0.77) |
| Baseline myocardial infarction | 1,476 | 131,731 | 1.29 (1.22−1.36) | 0.78 (0.76−0.79) |
| Baseline CHD | 1,901 | 131,306 | 1.27 (1.21−1.32) | 0.76 (0.75−0.78) |
| Baseline CHD or CHD death | 4,649 | 128,558 | 1.21 (1.18−1.25) | 0.75 (0.75−0.76) |
| CHD death (any where on the death certificate) | 3,479 | 129,728 | 1.17 (1.13−1.21) | 0.75 (0.74−0.76) |
| CHD death (primary cause on death certificate) | 2,927 | 130,280 | 1.19 (1.14−1.23) | 0.76 (0.75−0.77) |
| Primary definition (among unrelated participants) | 2,947 | 72,928 | 1.21 (1.16−1.25) | 0.75 (0.74−0.76) |
| **Tcheandjieu et al/PGS003446/ Multi−ancestry/485,464 SNPs** | | | | |
| Primary definition of premature CHD | 5,163 | 128,044 | 1.16 (1.12−1.20) | 0.75 (0.74−0.75) |
| Baseline angina | 494 | 132,713 | 1.11 (1.00−1.23) | 0.75 (0.73−0.77) |
| Baseline myocardial infarction | 1,476 | 131,731 | 1.28 (1.20−1.37) | 0.77 (0.76−0.79) |
| Baseline CHD | 1,901 | 131,306 | 1.23 (1.16−1.30) | 0.76 (0.75−0.77) |
| Baseline CHD or CHD death | 4,649 | 128,558 | 1.18 (1.13−1.22) | 0.75 (0.74−0.76) |
| CHD death (any where on the death certificate) | 3,479 | 129,728 | 1.13 (1.09−1.18) | 0.75 (0.74−0.76) |
| CHD death (primary cause on death certificate) | 2,927 | 130,280 | 1.15 (1.10−1.20) | 0.76 (0.75−0.76) |
| Primary definition (among unrelated participants) | 2,947 | 72,928 | 1.16 (1.11−1.22) | 0.75 (0.74−0.76) |
| **Tamlander et al/ PGS001780/ European/1,087,958 SNPs** | | | | |
| Primary definition of premature CHD | 5,163 | 128,044 | 1.20 (1.16−1.23) | 0.75 (0.74−0.75) |
| Baseline angina | 494 | 132,713 | 1.10 (1.00−1.21) | 0.75 (0.73−0.77) |
| Baseline myocardial infarction | 1,476 | 131,731 | 1.29 (1.22−1.36) | 0.77 (0.76−0.79) |
| Baseline CHD | 1,901 | 131,306 | 1.23 (1.17−1.29) | 0.76 (0.75−0.77) |
| Baseline CHD or CHD death | 4,649 | 128,558 | 1.20 (1.17−1.24) | 0.75 (0.75−0.76) |
| CHD death (any where on the death certificate) | 3,479 | 129,728 | 1.18 (1.14−1.23) | 0.75 (0.74−0.76) |
| CHD death (primary cause on death certificate) | 2,927 | 130,280 | 1.19 (1.14−1.24) | 0.76 (0.75−0.77) |
| Primary definition (among unrelated participants) | 2,947 | 72,928 | 1.19 (1.15−1.24) | 0.75 (0.74−0.76) |
| **Patel et al/PGS003725/ Multi−ancestry/1,273,824 SNPs** | | | | |
| Primary definition of premature CHD | 5,163 | 128,044 | 1.24 (1.20−1.28) | 0.75 (0.74−0.76) |
| Baseline angina | 494 | 132,713 | 1.14 (1.04−1.24) | 0.75 (0.73−0.77) |
| Baseline myocardial infarction | 1,476 | 131,731 | 1.34 (1.27−1.41) | 0.78 (0.76−0.79) |
| Baseline CHD | 1,901 | 131,306 | 1.27 (1.22−1.34) | 0.76 (0.75−0.77) |
| Baseline CHD or CHD death | 4,649 | 128,558 | 1.25 (1.21−1.28) | 0.75 (0.75−0.76) |
| CHD death (any where on the death certificate) | 3,479 | 129,728 | 1.23 (1.19−1.28) | 0.75 (0.74−0.76) |
| CHD death (primary cause on death certificate) | 2,927 | 130,280 | 1.23 (1.19−1.28) | 0.76 (0.75−0.77) |
| Primary definition (among unrelated participants) | 2,947 | 72,928 | 1.24 (1.20−1.29) | 0.75 (0.74−0.76) |
| **Inouye et al/PGS000018/ European/1,720,068 SNPs** | | | | |
| Primary definition of premature CHD | 5,163 | 128,044 | 1.19 (1.15−1.23) | 0.75 (0.74−0.75) |
| Baseline angina | 494 | 132,713 | 1.14 (1.03−1.27) | 0.75 (0.73−0.77) |
| Baseline myocardial infarction | 1,476 | 131,731 | 1.30 (1.22−1.38) | 0.77 (0.76−0.79) |
| Baseline CHD | 1,901 | 131,306 | 1.25 (1.18−1.32) | 0.76 (0.75−0.77) |
| Baseline CHD or CHD death | 4,649 | 128,558 | 1.21 (1.17−1.25) | 0.75 (0.75−0.76) |
| CHD death (any where on the death certificate) | 3,479 | 129,728 | 1.17 (1.13−1.22) | 0.75 (0.74−0.76) |
| CHD death (primary cause on death certificate) | 2,927 | 130,280 | 1.19 (1.14−1.24) | 0.76 (0.75−0.76) |
| Primary definition (among unrelated participants) | 2,947 | 72,928 | 1.20 (1.14−1.25) | 0.75 (0.74−0.76) |
| **Khera et al/PGS000013/ European/6,472,620 SNPs** | | | | |
| Primary definition of premature CHD | 5,163 | 128,044 | 1.17 (1.14−1.20) | 0.75 (0.74−0.75) |
| Baseline angina | 494 | 132,713 | 1.16 (1.06−1.26) | 0.75 (0.73−0.77) |
| Baseline myocardial infarction | 1,476 | 131,731 | 1.25 (1.19−1.31) | 0.77 (0.76−0.79) |
| Baseline CHD | 1,901 | 131,306 | 1.22 (1.17−1.28) | 0.76 (0.75−0.77) |
| Baseline CHD or CHD death | 4,649 | 128,558 | 1.18 (1.15−1.22) | 0.75 (0.75−0.76) |
| CHD death (any where on the death certificate) | 3,479 | 129,728 | 1.15 (1.11−1.19) | 0.75 (0.74−0.76) |
| CHD death (primary cause on death certificate) | 2,927 | 130,280 | 1.16 (1.12−1.21) | 0.76 (0.75−0.77) |
| Primary definition (among unrelated participants) | 2,947 | 72,928 | 1.17 (1.13−1.21) | 0.75 (0.74−0.76) |

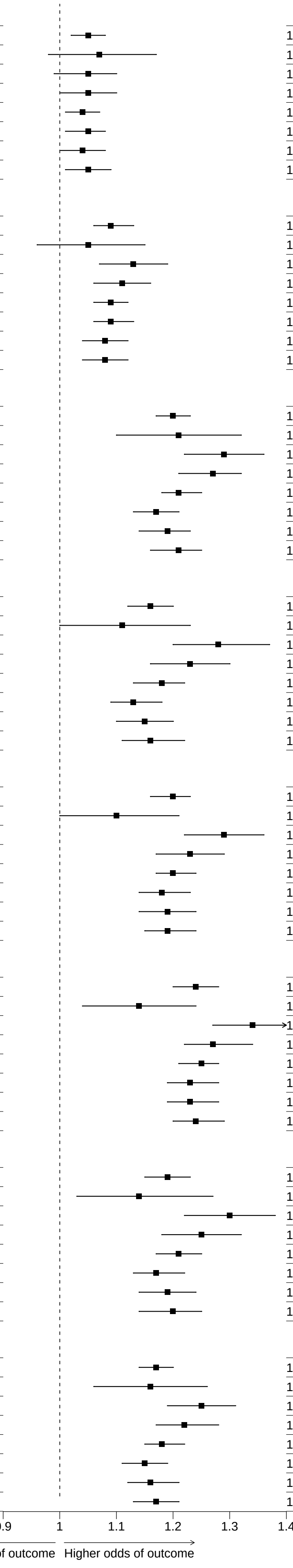


**Webfigure 17: Sensitivity analyses (varying the CHD outcome) with full adjustments**

Odd ratios (OR) were estimated with regression models adjusted for sex, age at baseline, 7 genetic principal components, waist-to-hip ratio, systolic and diastolic blood pressures, education attainment level, smoking status, and diabetes at baseline.

| PRS name | CHD cases | CHD controls | OR per SD (95% CI) | AUC (95%CI) |
|---|---|---|---|---|
| **Tada et al/PGS000011/ European/44 SNPs** | | | | |
| Partially adjusted | 2,947 | 72,928 | 1.06 (1.02−1.10) | 0.72 (0.71−0.72) |
| Fully adjusted | 2,947 | 72,928 | 1.05 (1.01−1.09) | 0.75 (0.74−0.76) |
| **Oni−Orisan et al/no PGS ID/ European/141 SNPs** | | | | |
| Partially adjusted | 2,947 | 72,928 | 1.09 (1.05−1.13) | 0.72 (0.71−0.72) |
| Fully adjusted | 2,947 | 72,928 | 1.08 (1.04−1.12) | 0.75 (0.74−0.76) |
| **Koyama et al/PGS000337/ Japanese and European/64,185 SNPs** | | | | |
| Partially adjusted | 2,947 | 72,928 | 1.23 (1.19−1.28) | 0.72 (0.71−0.73) |
| Fully adjusted | 2,947 | 72,928 | 1.21 (1.16−1.25) | 0.75 (0.74−0.76) |
| **Tcheandjieu et al/PGS003446/ Multi−ancestry/485,464 SNPs** | | | | |
| Partially adjusted | 2,947 | 72,928 | 1.20 (1.14−1.25) | 0.72 (0.71−0.73) |
| Fully adjusted | 2,947 | 72,928 | 1.16 (1.11−1.22) | 0.75 (0.74−0.76) |
| **Tamlander et al/ PGS001780/ European/1,087,958 SNPs** | | | | |
| Partially adjusted | 2,947 | 72,928 | 1.22 (1.17−1.27) | 0.72 (0.71−0.73) |
| Fully adjusted | 2,947 | 72,928 | 1.19 (1.15−1.24) | 0.75 (0.74−0.76) |
| **Patel et al/PGS003725/ Multi−ancestry/1,273,824 SNPs** | | | | |
| Partially adjusted | 2,947 | 72,928 | 1.29 (1.25−1.34) | 0.72 (0.71−0.73) |
| Fully adjusted | 2,947 | 72,928 | 1.24 (1.20−1.29) | 0.75 (0.74−0.76) |
| **Inouye et al/PGS000018/ European/1,720,068 SNPs** | | | | |
| Partially adjusted | 2,947 | 72,928 | 1.23 (1.17−1.28) | 0.72 (0.71−0.73) |
| Fully adjusted | 2,947 | 72,928 | 1.20 (1.14−1.25) | 0.75 (0.74−0.76) |
| **Khera et al/PGS000013/ European/6,472,620 SNPs** | | | | |
| Partially adjusted | 2,947 | 72,928 | 1.18 (1.14−1.22) | 0.72 (0.71−0.73) |
| Fully adjusted | 2,947 | 72,928 | 1.17 (1.13−1.21) | 0.75 (0.74−0.76) |

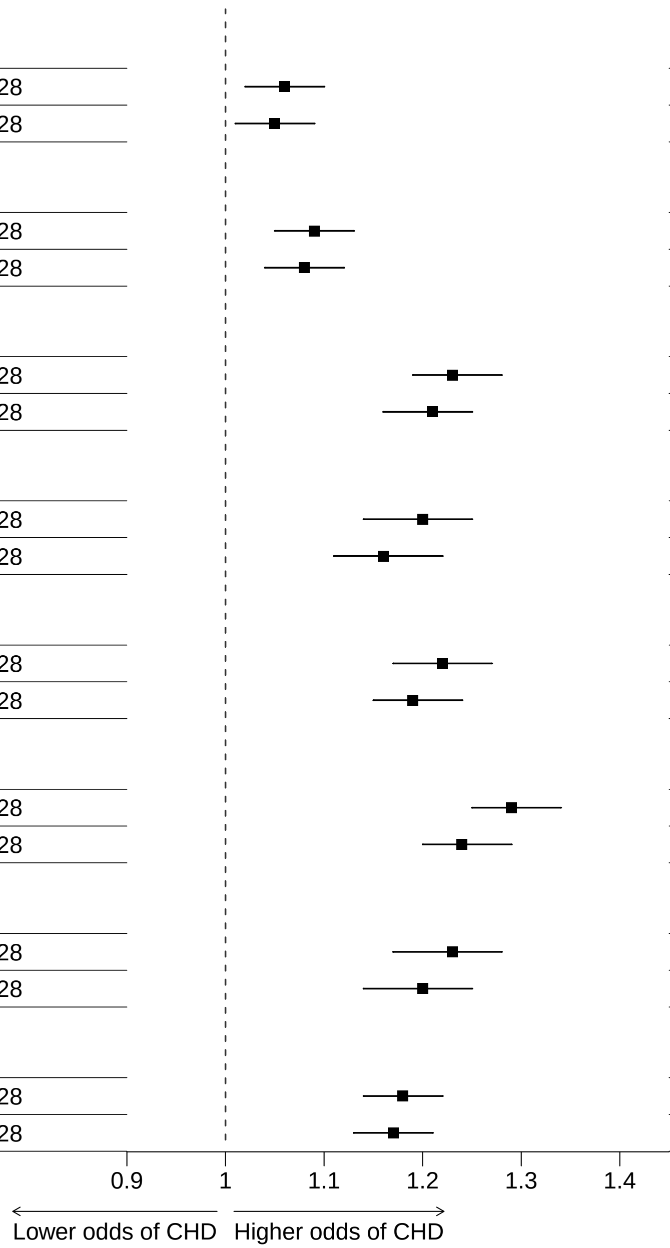


**Webfigure 18: Sensitivity analyses (primary CHD definition) among participants unrelated to the 3rd degree**
Analyses as for Figure 2

| PRS name | CHD cases | CHD controls | OR per SD (95% CI) | AUC (95%CI) |
|---|---|---|---|---|
| **Tada et al/PGS000011/ European/44 SNPs** | | | | |
| Partially adjusted | 7,155 | 130,236 | 1.06 (1.03−1.09) | 0.76 (0.76−0.77) |
| Fully adjusted | 7,155 | 130,236 | 1.06 (1.03−1.08) | 0.78 (0.77−0.78) |
| **Oni−Orisan et al/no PGS ID/ European/141 SNPs** | | | | |
| Partially adjusted | 7,155 | 130,236 | 1.09 (1.06−1.12) | 0.76 (0.76−0.77) |
| Fully adjusted | 7,155 | 130,236 | 1.08 (1.06−1.11) | 0.78 (0.77−0.78) |
| **Koyama et al/PGS000337/ Japanese and European/64,185 SNPs** | | | | |
| Partially adjusted | 7,155 | 130,236 | 1.19 (1.16−1.22) | 0.77 (0.76−0.77) |
| Fully adjusted | 7,155 | 130,236 | 1.17 (1.14−1.20) | 0.78 (0.77−0.78) |
| **Tcheandjieu et al/PGS003446/ Multi−ancestry/485,464 SNPs** | | | | |
| Partially adjusted | 7,155 | 130,236 | 1.15 (1.12−1.19) | 0.76 (0.76−0.77) |
| Fully adjusted | 7,155 | 130,236 | 1.13 (1.09−1.16) | 0.78 (0.77−0.78) |
| **Tamlander et al/ PGS001780/ European/1,087,958 SNPs** | | | | |
| Partially adjusted | 7,155 | 130,236 | 1.19 (1.15−1.22) | 0.76 (0.76−0.77) |
| Fully adjusted | 7,155 | 130,236 | 1.16 (1.13−1.19) | 0.78 (0.77−0.78) |
| **Patel et al/PGS003725/ Multi−ancestry/1,273,824 SNPs** | | | | |
| Partially adjusted | 7,155 | 130,236 | 1.24 (1.21−1.27) | 0.77 (0.76−0.77) |
| Fully adjusted | 7,155 | 130,236 | 1.20 (1.17−1.23) | 0.78 (0.78−0.79) |
| **Inouye et al/PGS000018/ European/1,720,068 SNPs** | | | | |
| Partially adjusted | 7,155 | 130,236 | 1.19 (1.15−1.22) | 0.76 (0.76−0.77) |
| Fully adjusted | 7,155 | 130,236 | 1.16 (1.13−1.20) | 0.78 (0.77−0.78) |
| **Khera et al/PGS000013/ European/6,472,620 SNPs** | | | | |
| Partially adjusted | 7,155 | 130,236 | 1.15 (1.12−1.18) | 0.76 (0.76−0.77) |
| Fully adjusted | 7,155 | 130,236 | 1.14 (1.11−1.17) | 0.78 (0.77−0.78) |

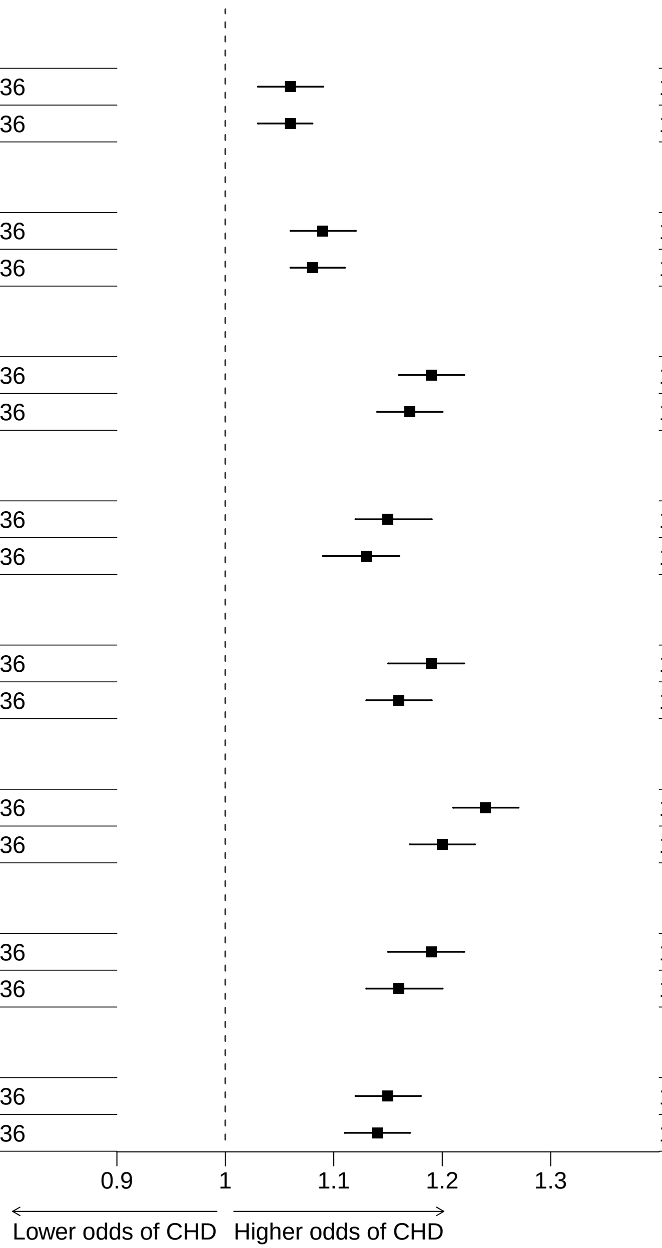


**Webfigure 19: Odds of CHD before age 90 years per 1SD increase in each PRS, among participants age 35-89 at recruitment**
Analyses as for Figure 2

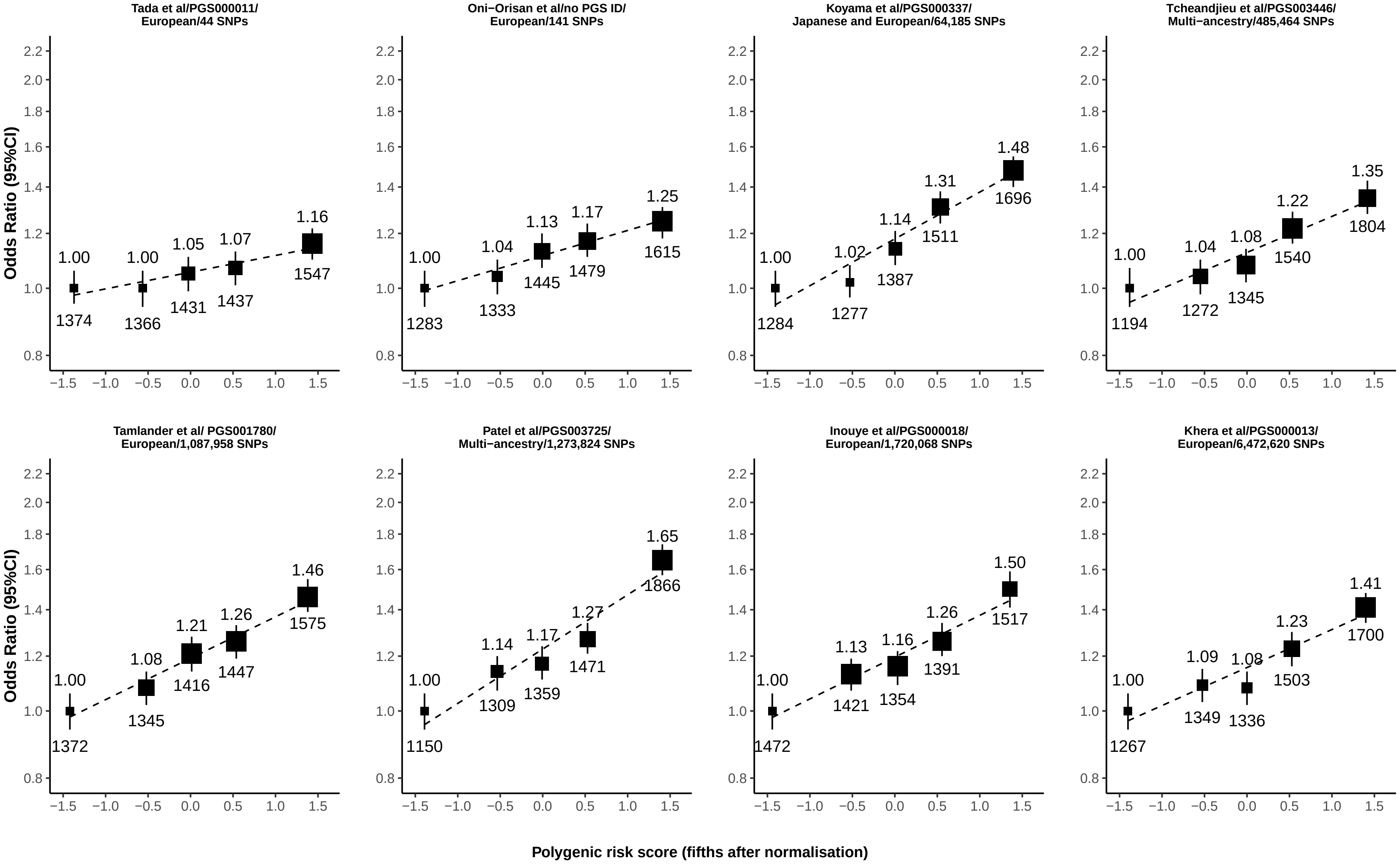


**Webfigure 20: Odds of CHD before age 90 years by fifth of each PRS, among participants age 35-89 years at recruitment**
Analyses as for Figure 1
